# Identification of occupations susceptible to high exposure and risk associated with multiple toxicants in an observational study: National Health and Nutrition Examination Survey 1999-2014

**DOI:** 10.1101/2021.11.23.21266764

**Authors:** Vy Kim Nguyen, Justin Colacino, Chirag J Patel, Maureen Sartor, Olivier Jolliet

## Abstract

**Background:** According to the World Health Organization, occupational exposures to hazardous chemicals are estimated to cause over 370,000 premature annual deaths. The risks due to multiple workplace chemical exposures, and those occupations most susceptible to the resulting health effects, remain poorly characterized.

**Objectives:** The aim of this study is to identify occupations with elevated toxicant biomarker concentrations and increased health risk associated with toxicant exposures in a working US population from diverse categories of occupation. More specifically, we aim to 1) define differences in chemical exposures based on occupation description, 2) identify occupational groups with similar chemical exposure profiles, and 3) identify occupational groups with chemical biomarker levels exceeding acceptable health-based biomarker levels.

**Methods:** For this observational study of 51,008 participants, we used data from the 1999-2014 National Health and Nutrition Examination Survey. We characterized differences in chemical exposures by occupational group for 129 chemicals by applying a series of generalized linear models with the outcome as biomarker concentrations and the main predictor as the occupational groups, adjusting for age, sex, race/ethnicity, poverty income ratio, study period, and biomarker of tobacco use. We identified groups of occupations with similar chemical exposure profiles via hierarchical clustering. For each occupational group, we calculated percentages of participants with chemical biomarker levels exceeding acceptable health-based guidelines.

**Results:** Blue collar workers from “Construction”, “Professional, Scientific, Technical Services”, “Real Estate, Rental, Leasing”, “Manufacturing”, and “Wholesale Trade” have higher biomarker levels of toxic chemicals such as several heavy metals, acrylamide, glycideamide, and several volatile organic compounds compared to their white-collar counterparts. For these toxicants, 1-58% of blue-collar workers from these industries have toxicant concentrations exceeding acceptable levels.

**Discussion:** Blue collar workers have toxicant levels higher relative to their white-collar counterparts, often exceeding acceptable levels associated with noncancer effects. Our findings identify multiple occupations to prioritize for targeted interventions and health policies to monitor and reduce high toxicant exposures.

## 1. Introduction

Data from the World Health Organization suggest that exposures to hazardous chemicals in an occupational setting are responsible for over 370,000 premature deaths annually (Lim et al., 2012; Stanaway et al., 2018). Such findings lend urgency to characterize occupational exposures to identify workers from which industries or jobs are susceptible to adverse effects from toxicant exposures. Many studies tend to focus on one chemical or one chemical family to evaluate occupational exposures (Birks et al., 2016; Brenner et al., 2015). In doing so, these studies may miss the complete picture of being exposed to a slew of toxicants if the focus is only directed at one chemical or one chemical family. Furthermore, exposures to multiple chemicals can further increase the risk of a disease. For a few studies that have investigated across multiple chemicals, they have narrowed their focus on a limited set of industries and job titles (Kijko et al., 2016; Mater et al., 2016). Thus, there is a need for a comprehensive, untargeted approach to study occupational exposures for a wide range of chemicals across a variety of occupations.

Furthermore, many studies on occupational exposures use estimates of exposures based on job titles or air measurements at the workplace (Birks et al., 2016; Kijko et al., 2016; Mater et al., 2016). These indirect measures are limited in their ability to accurately quantify the distribution of chemical exposures within the human body. In contrast, human biomonitoring provides a more direct estimate of exposure while also integrating exposures which derive from multiple sources and pathways. In addition, another advantage of biomonitoring data is that it provides an internal dose that can be related to a toxicological response (Hays et al., 2007). In particular, biomonitoring equivalents define a concentration cutoff of a chemical or its metabolites in a biological medium such as blood, urine, or serum based on acceptable exposure values such as reference dose, tolerable daily intakes, or minimal risk levels (Aylward et al., 2013). Several studies have used biomonitoring equivalents as a screening method to evaluate risk from exposures to environmental toxicants in the general population (Aylward et al., 2013; Faure et al., 2020). However, few studies have used biomonitoring equivalents in an occupational context to determine prevalence of workers with concentrations above acceptable levels by industry and job description (Aylward et al., 2010; Hays et al., 2008). Such insight will help identify which toxicants and occupations should be prioritized for further human biomonitoring, health risk evaluation, and targeted interventions. Our goal is to broadly identify occupations susceptible to high exposure and risk associated with combinations of multiple toxicants. To accomplish this goal, we used data from the National Health and Nutrition Examination Survey (NHANES), which measures a broad range of 517 chemical biomarkers as part of their mission to assess the health and nutritional status of the US noninstitutionalized population. Occupational information, particularly the 21 industrial and 19 occupation codes, are also available. Our objectives are to 1) define differences in chemical exposures based on occupation description, 2) identify occupational groups with similar chemical exposure profiles, and 3) identify occupational groups with chemical biomarker levels exceeding acceptable health-based biomarker levels.

## 2. Methods

### 2.1 Study Population

Since 1999, the Centers for Disease Control (CDC) has conducted the Continuous NHANES to collect cross-sectional data on demographic, socioeconomic, dietary, and health-related information in the US population. For this analysis, we combined data from the chemical biomarker, demographic, and occupational datasets between years 1999-2014 for an initial sample of 82,091 participants. We categorized participants as different groups of unemployment status using the questionnaires on the type of work done last week and main reason for not working last week. We categorized the workers into their corresponding industry by using the publicly available industry code on the participants’ current job. We categorized participants into white- or blue-collar workers by using the publicly available occupational codes on the participants’ current job and the US Department of Labor definition of blue-collar (U.S. Department of Labor, 2019). Blue-collar workers are defined as workers who perform repetitive tasks with their hands, physical skill, and energy. The industry and occupational codes can be found at https://www.n.cdc.gov/nchs/nhanes/search/datapage.aspx?Component=Questionnaire. We tabulated the job occupation description and the collar category in Table S1. We then excluded participants under 16 years old (N = 30,987) as this is the minimum age at which NHANES recorded occupation status. We excluded participants who recorded their industry as “Blank but applicable” (N = 9). We also excluded participants (N = 87) from the following occupational groups if the sample size was less than 50 participants: blue-collar workers from “Armed Forces” and “Finance, Insurance” and white-collar workers from “Armed Forces”, “Private Household”, and “Mining”. These exclusion and inclusion criteria are further detailed in **Figure 1**. The resulting sample size of our studied population was 51,008 participants. Table S2 provides the sample size of each industry-collar combinations and unemployment.

**Figure 1.**
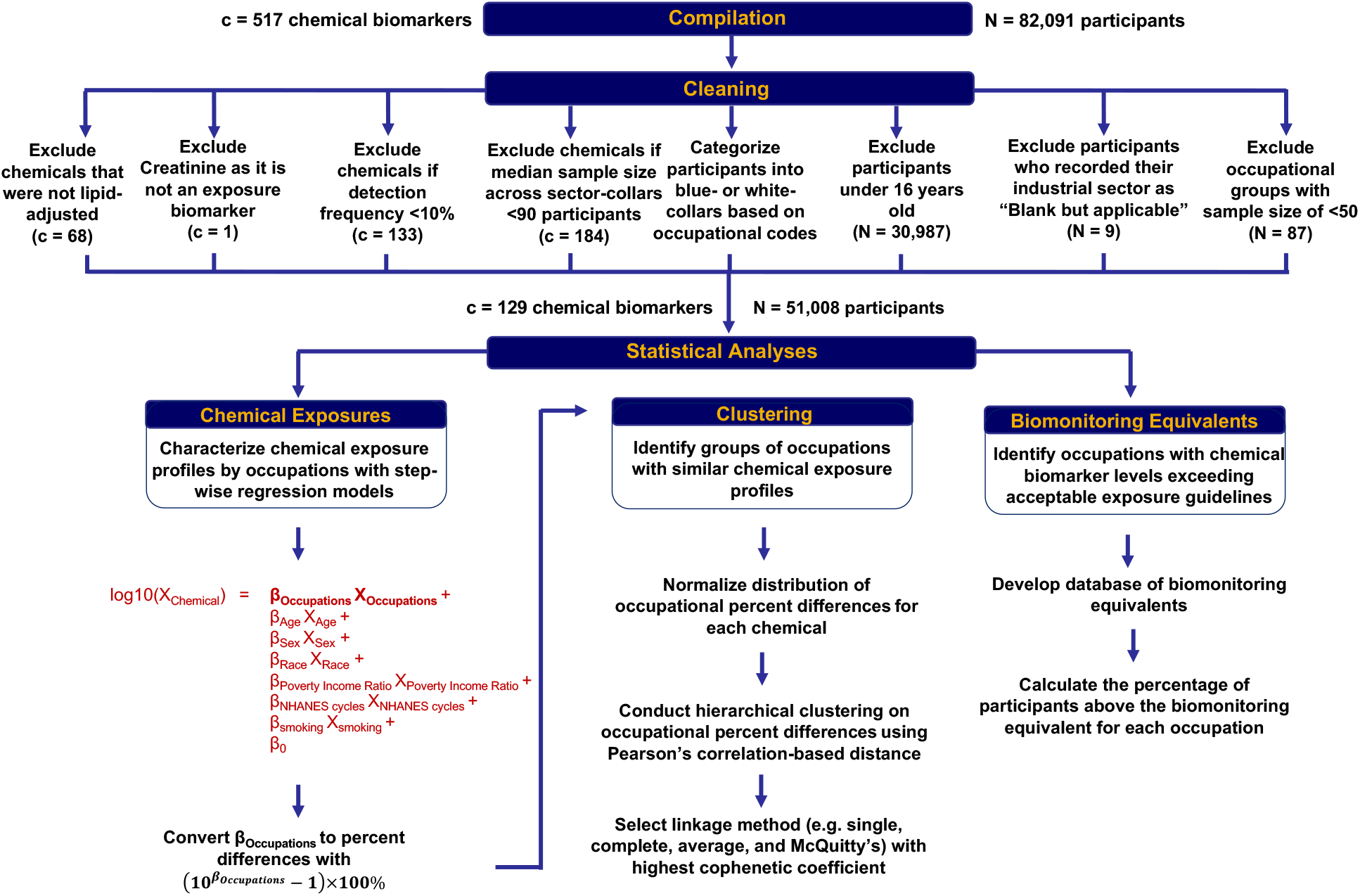
Schematic description on curation of chemical biomarker and inclusion criteria of participants and of the analytical methods used to characterize occupational variations in chemical exposures. Reference group for the analysis on the industry-collar combinations is white collars from public administration.

The National Center for Health Statistics research ethics review board provided ethical approval of the study. All participants provided written informed consent.

### 2.2 Chemical Biomarkers of Occupational Exposures

We defined chemical biomarker, *c*, as an indicator of environmental exposure that can be measured in blood, serum, or urine. We replaced all measurements below the limit of detection (LOD) with the LOD divided by the square root of 2, as recommended by the CDC (CDC, 2009) to produce reasonably unbiased means and standard deviations (Hornung & Reed, 1990). For di-2-ethylhexyl phthalate (DEHP) and arsenic separately, we calculated the sum of metabolites to compare with respect to the biomonitoring equivalents. We used mono-(2-ethyl-5-oxohexyl) phthalate, mono-(2-ethyl-5-hydroxyhexyl) phthalate, mono-(2-ethylhexyl) phthalate, and mono-2-ethyl-5-carboxypentyl phthalate to calculate the summation of DEHP metabolites. We calculated the summation of arsenic metabolites with monomethylarsonic acid and dimethylarsonic acid. Therefore, we have a total of c = 517 chemical biomarkers.

Then, we further excluded chemicals that a) have a median sample size of less than 90 participants across the occupational groups, b) have non-lipid adjusted measurements when lipid adjusted measurements are available and c) have a detection frequency less than 10%. We delineated in detail which chemical biomarkers were excluded from our analyses in Text S1. We tabulated the inclusion criteria for each chemical in Table S4. The final dataset for analysis consisted of 129 chemical biomarkers from 12 classes (Figure 1 and Table S5). We tabulated the sample size of each chemical in Table S6. We tabulated the distribution statistics for each chemical in Table S7. We displayed the detection frequency by each combination of chemical biomarker and occupational group in Figure S1. Laboratory methods used to measure the chemical biomarkers are provided at https://www.n.cdc.gov/nchs/nhanes/search/datapage.aspx?Component=Laboratory.

### 2.3 Statistical Analysis

We performed all analyses using R version 3·6·0. Our analytic code is publicly available on GitHub (https://github.com/vynguyen92/nhanes_occupational_exposures). We applied the survey weights to all our statistical models to 1) account for NHANES sampling designs and 2) enable the generalizability of our findings to the non-institutionalized, civilian US population.

We used multivariate regression models to evaluate differences in the chemical biomarker levels across the occupational groups, which includes the industry-collar combinations and unemployed groups. We conducted a series of stepwise linear regression models with the log10 transformed chemical measurements as the outcome variable and the main predictor as the occupation groups with the reference group as white collars from Public Administration. The selection of the reference group was based on the *a priori* hypothesis that white collar workers from Public Administration would be exposed to toxicants at relatively low levels for most toxicants. We adjusted for age (continuous), gender (categorical), race/ethnicity (categorical), poverty income ratio (continuous), NHANES cycle (continuous), and tobacco use/exposure status using serum cotinine levels (continuous). Race is self-reported by the participants. The poverty income ratio is defined by dividing the total family income by the poverty income line. A poverty income ratio lower than 1 implies that the participant’s total family income is below the poverty income line. For ease of interpretation, the regression coefficients for the occupational groups were converted to percent differences [10^coefficient^ - 1] × 100. To identify significant comparisons while maintaining a lower false positive rate, we used the False Discovery Rate (FDR) method on the p-values of the regression coefficients pertaining to the occupational groups (Benjamini & Hochberg, 1995).

The non-random sparsity of the chemical biomarker dataset in the NHANES worker population creates challenges in applying machine learning techniques to group individual workers together based on similarity in chemical exposure profiles. Applying most machine learning techniques requires a complete dataset (Soley-Bori, 2013). However, as no worker has data available for all studied toxicants (Table S8, Figure S2), we cannot characterize the chemical profile for each individual worker. Such challenges limited studies to characterizing combinations of toxicant exposures within a specific chemical family, but workers are exposed to multiple toxicants across a variety of chemical families at their workplace. Instead of characterizing the toxicant profile of a given worker, we can characterize the profile for a group of individual workers to address this sparsity issue by identifying clusters of occupations with similar chemical exposure profiles. Thus, we performed hierarchical agglomerative clustering analyses on the dataset of percent differences for the occupational groups. Text S2 describes the methodology used to identify clusters of occupations with similar chemical exposure profiles.

To identify susceptible occupations with chemical biomarker levels exceeding acceptable exposure levels, we compared workers’ biomarker levels to biomonitoring equivalents, i.e. the internal levels corresponding to these acceptable exposures. We, first, performed a literature review to develop a database of biomonitoring equivalents (Table S10-S11). There are three types of effects for the biomonitoring equivalents: noncancer, inhalation cancer, and ingestion cancer. Noncancer effects can include mutagenicity, developmental toxicity, neurotoxicity, and reproductive toxicity. Cancer effects are specific for different route of exposures. Then we used this database to calculate the percentage of participants above the biomonitoring equivalent for each occupation and each chemical. We used hierarchical clustering to group the occupations who have similar profiles in chemical biomarker levels exceeding acceptable levels.

Text S3 provided full details on the methodology to quantify the contribution of occupation in explaining chemical biomarker levels. Text S4 provided full details on the sensitivity analyses to characterize the influence of smoking on differences in chemical biomarker levels by occupation.

## 3. Results

All figures are available on our interactive app at https://chiragjp.shinyapps.io/nhanes_occupational_exposures/.

### 3.1 Study Population

**Table 1 and 2** presents population characteristics for the 51,008 NHANES participants from 1999-2014. **Figure 2** shows the percentage of categories for age group, sex, race, poverty income ratio, smoking status, and study period for each occupational group. Participants working in blue-collar jobs tend to be on average younger compared to those working in white-collar jobs (**Figure 2A**). Blue-collar jobs are primarily occupied by males, while females tend to work in private household, health care, and education (**Figure 2B**). White-collar jobs are predominantly comprised of Non-Hispanic White participants, whereas blue-collar workers tend to be more diverse (**Figure 2C**). There is a socioeconomic gradient with white-collar workers having higher poverty income ratio (i.e. lower socioeconomic status) compared to blue-collar workers (**Figure 2D**). There is a substantial proportion of active smokers in blue collar jobs as well as those “Looking for work”, “Disabled”, or “On layoff”, whereas the proportions of active smokers are much lower in white-collar jobs (**Figure 2E**). **There is** a relative uniform distribution of participants by study period for each occupation (**Figure 2F**). These figures are available on our interactive app at https://chiragjp.shinyapps.io/nhanes_occupational_exposures/.

**Table 1.**
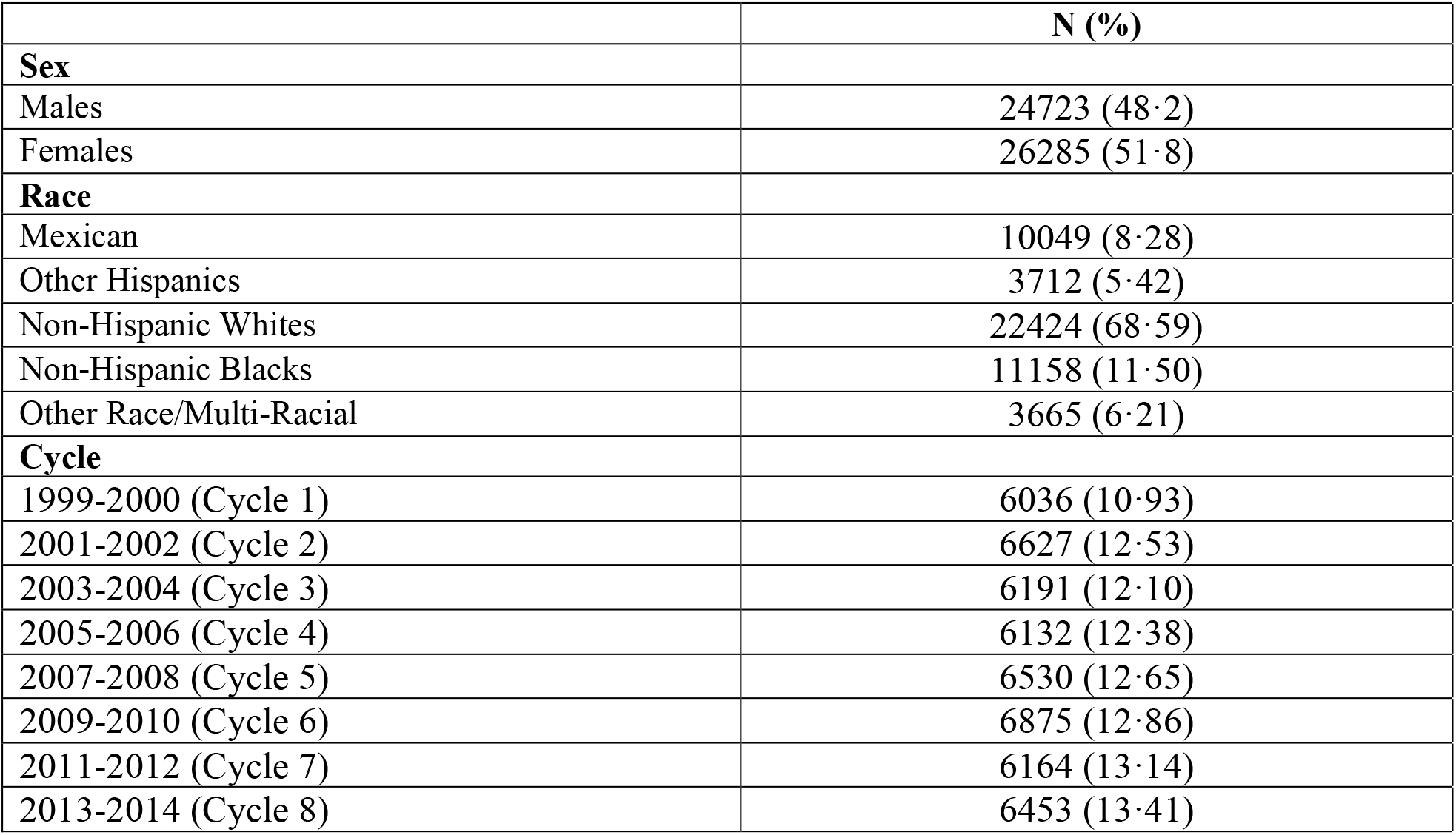
Population statistics of the categorical variables for 51,008 NHANES participants who are eligible to have an occupation title. NHANES sampling design is accounted in calculating percentages (%), while counts (N) pertains to the number of NHANES participants.

**Table 2.** Distribution statistics of the categorical variables for 51,008 NHANES participants who are eligible to have an occupation title. NHANES sampling design is accounted in the calculations of the distribution statistics, while counts (N) pertains to the number of NHANES participants.

|  | N (%) | Minimum | 5<sup>th</sup> | 10th | Median | Mean (SE) | 90<sup>th</sup> | 99<sup>th</sup> | Maximum |
| --- | --- | --- | --- | --- | --- | --- | --- | --- | --- |
| Age (years) | 51008 (100) | 16 | 17 | 21 | 44 | 44.7 (0.20) | 71 | 83 | 85 |
| Poverty Income Ratio (-) | 46441 (91.0) | 0 | 0.49 | 0.77 | 2.86 | 2.93 (0.031) | 5 | 5 | 5 |
| Serum cotinine (ng/mL) | 45376 (88.9) | 0.011 | 0.011 | 0.011 | 0.061 | 58.55 (1.56) | 250 | 514 | 1820 |

**Figure 2.**
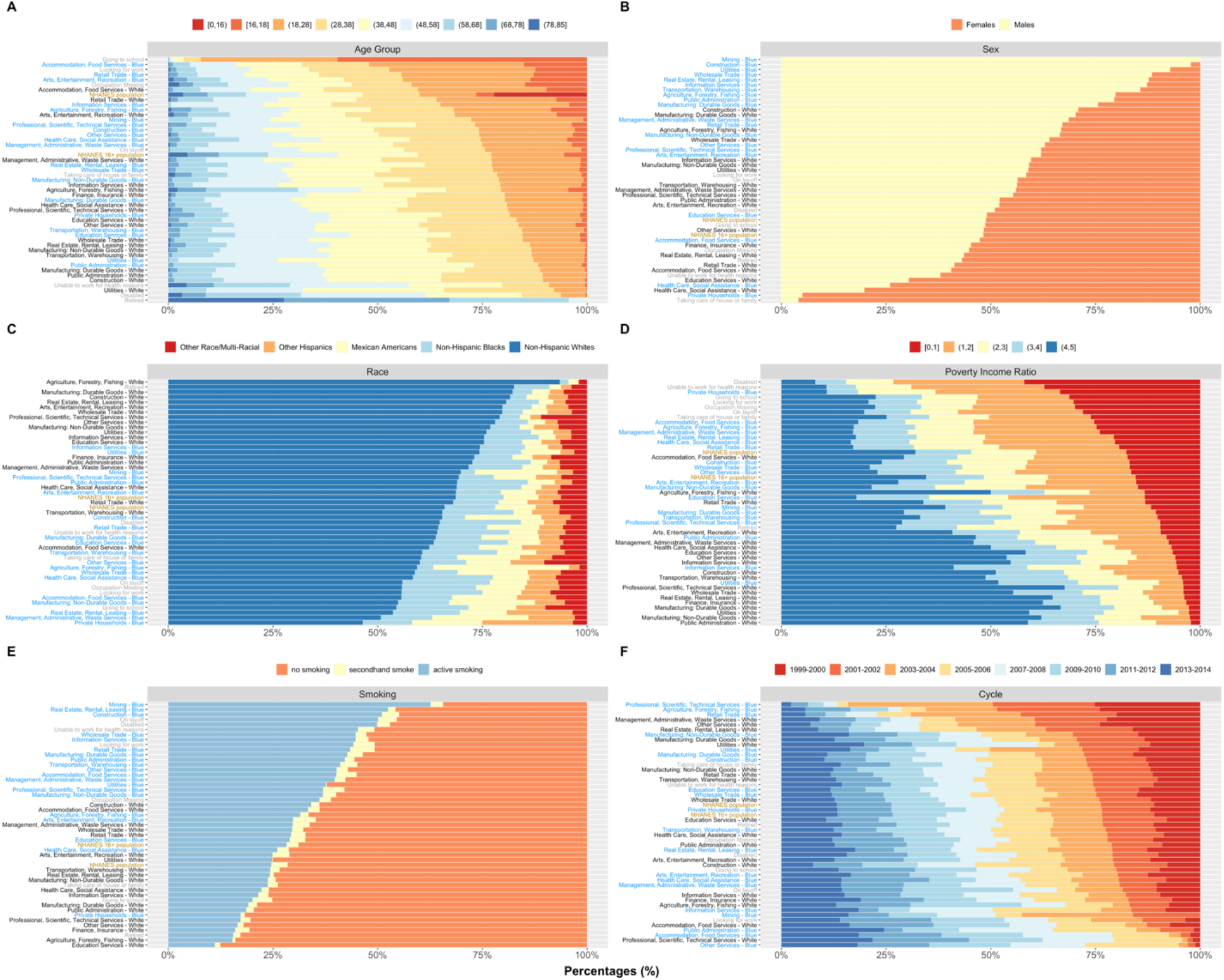
Panel of bar plots showing the percentage of participants by A) age group, B) sex, C) race/ethnicity, D) poverty income ratio (PIR), E) smoking status, and F) NHANES cycle for each industry-collar combination and unemployment status. The occupational groups are ordered in ascending order based on percentage of A) participants who are 28 years and younger, B) males, C) Non-Hispanic Whites, D) PIR = [0,1] (i.e. participants who are below the poverty income line), E) participants who do not smoke, and F) participants in 1999-2002. The “NHANES Population” consists of all participants in 1999-2014. The “NHANES 16+ Population” consists of participants in 1999-2014 and are 16 years old or older. Smoking status is defined using serum cotinine levels: no smoking ≤ 1 ng/mL, secondhand smoke 1-3 ng/mL, and active smoking > 3 ng/mL. These individual figures are available on our interactive app at https://chiragjp.shinyapps.io/nhanes_occupational_exposures/.

### 3.2 Differences in Chemical Biomarker Levels by Occupational Groups

**Figure 3** displays the differences in chemical biomarker levels across the occupational groups using regression and distribution statistics for the following chemicals: lead, m-/p-xylene, cotinine, 2,4-D, glycidamide, and sum of DEHP metabolites. We selected these chemicals for the at least one of the following reasons: 1) availability of having a biomonitoring equivalent and 2) existence of statistically significant differences in biomarker levels across the occupational groups. Biomonitoring equivalents are available for the selected chemicals except for cotinine. In Figure 3A, blood lead is, on average, significantly higher in most blue-collar workers and unemployed groups such as “Looking for work”, “On layoff”, “Disabled”, and “Retired” compared to the other white-collar workers. We observed a similar pattern in blood cadmium (Figure S4). In contrast, metabolites of mercury display the opposite exposure patterns to those of lead and cadmium. Total blood mercury levels of most blue-collar workers are substantially and significantly lower compared to those of white-collar workers (Figure S5). Similarly, m-/p-xylene (Figure 3B) and toluene (Figure S6) are higher in blue-collars and unemployed participants in “On layoff”, “Disabled”, and “Unable to work for health reasons” compared to white-collar workers. Similar results are observed for several Polyaromatic Hydrocarbons (PAHs) such as 1-pyrene, 2-fluorene, 3-fluorene, and 1-naphthol (Figure S7), but the signals are not as strong as those for toluene and m-/p-xylene. It is noteworthy that within the same industry such as “Professional, Scientific, Technical Services”, levels of m-/p-xylene are substantial different between white versus blue collars. Figure 3C shows a smoking gradient with blue-collar workers and unemployed participants having substantially higher levels of cotinine compared to white collars and the NHANES populations. NNAL, which is primarily found in tobacco products, shows a similar trend (Figure S8). The signals for the smoking related compounds are among the strongest and most substantial. Figure 4D shows how concentrations of a herbicide, 2,4-D, are significantly and substantially higher in blue-collar and white-collar workers from “Agriculture, Forestry, Fishing” along with blue-collar workers from “Information Services”. Though, none of the participants have 2,4-D levels exceeding the guideline biomonitoring equivalent levels. DEET acid, a metabolite of DEET and a common ingredient used in insect repellant, shows a similar pattern (Figure S9). In **Figure 3E**, glycidamide levels, on average, are significantly higher in blue collars from “Wholesale Trade”, “Other Services”, “Professional, Scientific, Technical Services”, “Retail Trade”, “Construction”, and “Arts, Entertainment, Recreation”. In addition, glycidamide levels are also significantly higher in white collars from “Accommodation, Food Services”, “Agriculture, Forestry, Fishing”, and “Retail Trade”. Acrylamide show similar pattern (Figure S10). These two chemicals are found in food prepared at high temperature via frying, baking, or roasting and are used in textile, paper processing, and cosmetics. In **Figure 3F**, levels of the sum of urinary DEHP metabolites are significantly higher in “Professional, Scientific, Technical Services” and “Accommodation, Food Services” compared to the reference group. Phthalates in general are used as plasticizers. On the contrary, mono-ethyl phthalate, an indicator of personal care product usage, are, on average higher, in the reference group (Figure S11).

**Figure 3.**
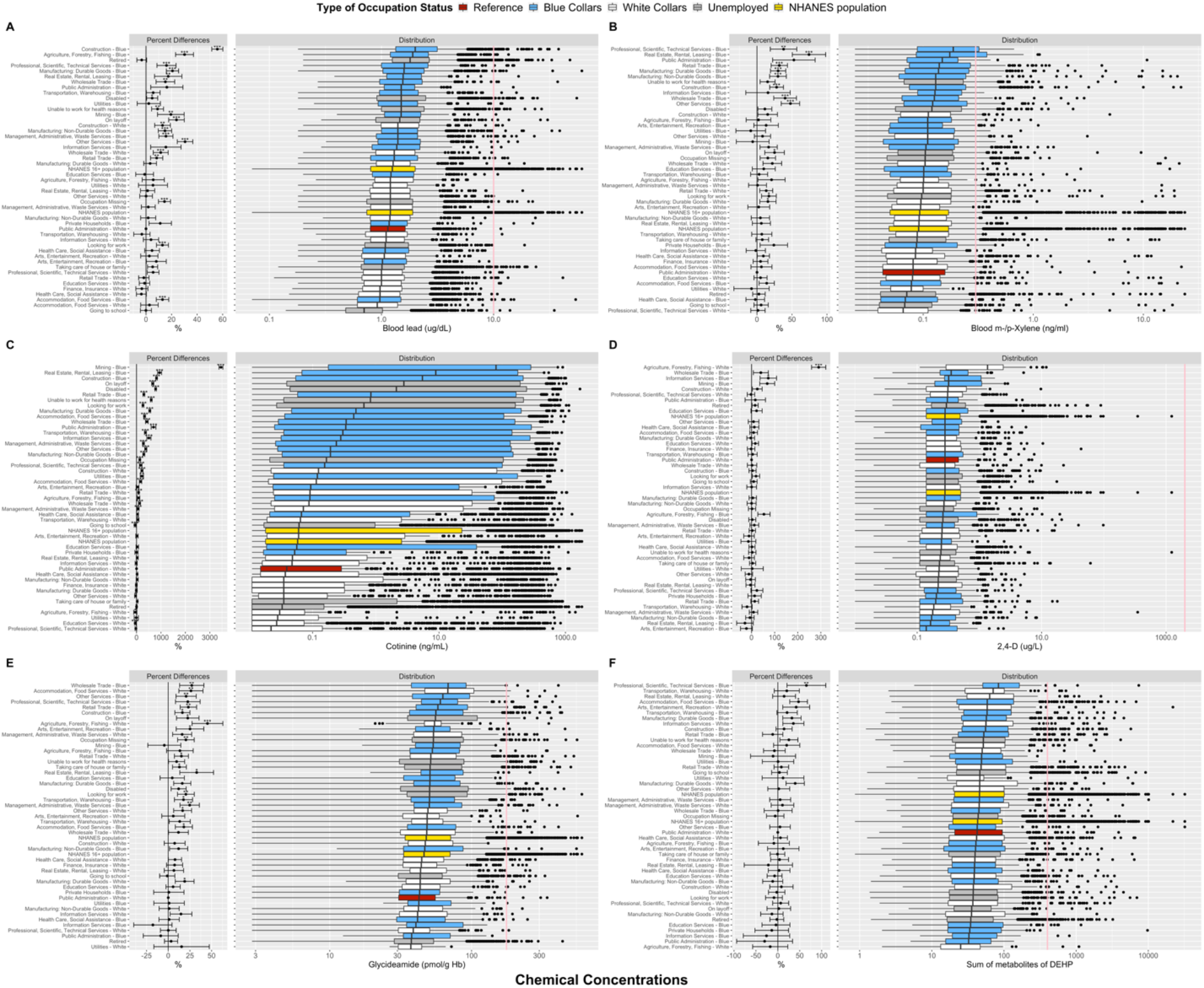
Panel of boxplots of chemical distribution for A) blood Lead, B) m-/p-Xylene, C) Cotinine, D) 2,4-D, E) Glycidamide, and F) sum of DEHP metabolites. Purple triangle represents the geometric mean of chemical levels for a given occupational group. The pink line represents the biomonitoring equivalent of the chemical for noncancer effects. The “NHANES Population” consists of participants in 1999-2014. The “NHANES 16+ Population” consists of participants in 1999-2014 and are 16 years old or older. Percent differences are derived from fully adjusted models, which were adjusted for age, sex, race/ethnicity, poverty income ratio, study period, and serum cotinine (biomarker of smoking). Reference group for the occupational groups is comprised of white collars from Public Administration. Number of asterisks indicate statistical significance of the percent differences: * (p-value ∈ (0·01, 0.05]), ** (p-value ∈ (0·001, 0.01]), and *** (p-value ≤ 0·001). The p-values corrected for multiple comparison with the Benjamini and Hochberg FDR procedure of 5%. These individual figures are available on our interactive app at https://chiragjp.shinyapps.io/nhanes_occupational_exposures/.

**Figure 4.**
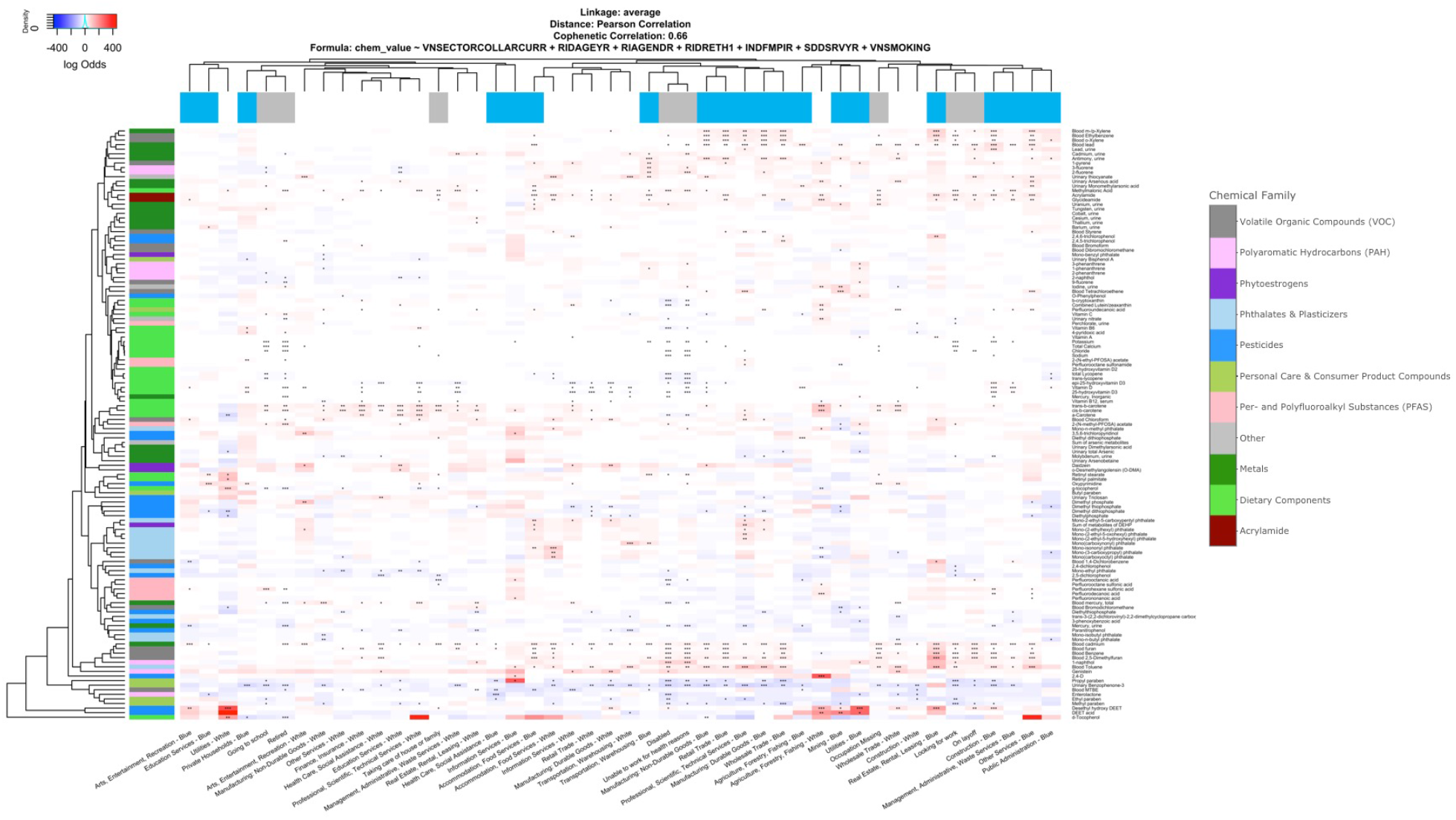
Heatmap of percent differences in chemical biomarker concentrations by occupational group, relative to white collars from Public Administration. Chemical biomarkers in white color indicates that the concentrations are the same between the given industry-collar combination and the reference group. The color bar for the columns represents the collar categorization and unemployment. The color bar for the rows represents the chemical classes. Blue presents the blue-collar workers. White represents the white-collar workers. Gray presents the unemployed participants. The dendrogram of the occupational groups is defined based on using the average linkage function with Pearson’s correlation-based distance. Results are adjusted for age, sex, race/ethnicity, poverty income ratio, study period, and serum cotinine (biomarker of smoking). Number of asterisks indicate statistical significance of the percent differences: * (p-value ∈ (0·01, 0·05]), ** (p-value ∈ (0·001, 0·01]), and *** (p-value ≤ 0·001). This figure is available on our interactive app at https://chiragjp.shinyapps.io/nhanes_occupational_exposures/.

### 3.3 Chemical Exposure Profiles

**Figure 4** shows differences in chemical exposure profiles by occupational groups for the 129 studied chemicals. The chemical exposure profiles of blue-collar workers are more similar to those of other blue-collar workers than to their white-collar counterparts. For example, blue-collar workers from “Construction”, “Other Services”, “Professional, Scientific, Technical Services”, “Real Estate, Rental, Leasing”, “Manufacturing”, and “Wholesale Trade” have some of the highest biomarker levels of heavy metals, such as cadmium and lead, PAHs, and volatile organic chemicals (VOCs), including m-/p-xylene and toluene, but have lower levels of arsenic and mercury metabolites and BP-3, which is a UV blocking chemical used in sunscreen. There are some blue collars who have higher levels of dietary components such as orange or red plant pigments found in fruits and vegetables such as trans–b–carotene, cis–b–carotene, and a–carotene and a form of vitamin E such as d-Tocopherol. Interestingly, participants who are “Looking for work”, “On layoff”, “Disabled”, “Unable to work for health reasons”, and “Occupation Missing” have similar chemical exposure profiles to the aforementioned blue-collar workers. Participants who report being “Disabled” or “Unable to work for health reasons” have significantly lower levels of several dietary components. Within the food services cluster, which includes blue and white collars from “Transportation, Warehousing” and “Accommodation, Food Services”, the phthalates signal is particularly stronger in the blue-collar workers from “Accommodation, Food Services”. The far left of the heatmap consists of mostly white-collar workers. These occupational groups have the most similar chemical exposure profiles to that of white-collar workers from “Public Administration” as the percent differences across most studied chemicals are near 0, i.e. the blue and red boxes are faded. We excluded smoking-related compounds such as NNAL and Cotinine from the figure, or else we would not be able to observe any signals from the other chemical biomarkers (Figure S12).

### 3.4 Occupational Groups with Chemical Biomarker Levels Exceeding Acceptable Guidelines

**Figure 5** shows the percentage of a given occupational group with chemical biomarker levels exceeding the biomonitoring equivalents for noncancer effects. Our hierarchical clustering analysis on the occupational groups show that the left half of Figure 5 includes predominantly blue collars and unemployed groups, while the right half includes predominantly white collars. This suggests that blue collars and unemployed groups have similar chemical exposure profiles to each other, and such profiles are different from the chemical profiles of the white collars. Several blue collar jobs along with unemployed groups such as those who are “On layoff”, “Unable to work for health reasons”, and “Disabled” have some of the highest percentages of participants with biomarker levels exceeding acceptable health levels for VOCs such as m-/p-xylene, benzene, pesticides such as 3–phenoxybenzoic acid, heavy metals such as cadmium, lead, and arsenic, metabolites of DEHP, and acrylamide and its metabolite glycideamide. These findings suggest that these toxicants for the aforementioned occupations may be further monitored to understand why biomarker levels are exceeding acceptable guidelines. Figure S13-S14 shows the percentage of participants with excessive toxicant levels for the cancer effects.

**Figure 5.**
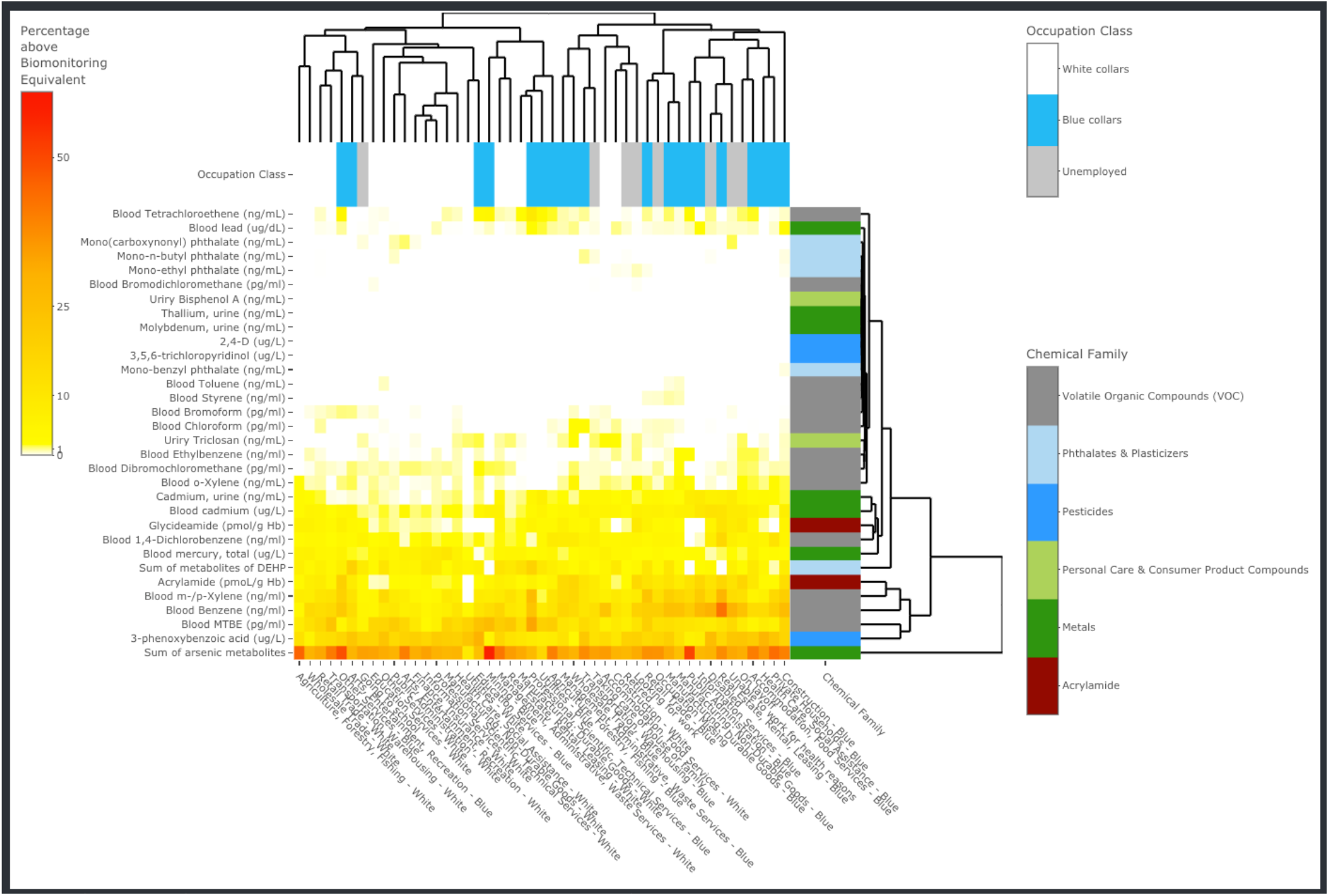
Heatmap of percentages of workers with biomarker levels exceeding biomonitoring equivalents for noncancer effects. Chemical biomarkers in white color indicates that no worker in a given occupational group has biomarker levels exceeding acceptable guidelines. The color bar for the columns represents the collar categorization and unemployment. Blue presents the blue-collar workers. White represents the white-collar workers. Gray presents the unemployed participants. This figure is available on our interactive app at https://chiragjp.shinyapps.io/nhanes_occupational_exposures/.

## 4. Discussion

In this study, we systematically characterize differences in chemical biomarker levels across a diverse suite of chemical contaminants and occupations. This is the first application of hierarchical clustering on differences by chemical exposures to identify groups of workers with similar chemical exposure profiles. This is also the first study to determine the percentage of a given occupation who are exceeding acceptable levels for a broad set of toxicants. Furthermore, this is the first application of hierarchical clustering to systematically identify which chemicals for which occupations have biomarker levels exceeding acceptable guidelines. Our findings are informative for identifying which workers are susceptible to higher exposures from which toxicants.

Contact with products and equipment may explained higher biomarker levels of heavy metals such as lead and cadmium, PAHs, and VOCs such as toluene and benzene found in blue-collar workers. Higher lead levels found in blue collars may be due to the presence of lead in old and commercial paint, car parts, batteries, glass, and consumer products made of plastics (CDC, 2018). In addition, this same group of workers may be exposed to cadmium via industrial uses of cadmium in making batteries, plating, pigments, and plastics (Miao & Ji, 2019). Sources of occupational PAH exposures to this group may be due to engaging in tasks that involve combustion emission (Raymond, 1998). Similarly, higher VOCs levels may also be due to working with products containing VOCs (Minnesota Department of Health, 2020). Overall, higher biomarker levels of heavy metals, PAHs, and VOCs in predominantly blue-collar workers may be due to contact with products containing these chemicals. This suggests that the forementioned blue collars should be further examined to understand sources of exposures for such toxicants and reasons for exceeding acceptable levels.

On the other hand, behaviors associated with higher socioeconomic status may explain why most white-collar workers have higher levels of metabolites of arsenic and mercury along with a biomarker of sunscreen use, BP-3. While arsenic (CDC, 2019a) and mercury (CDC, 2019b) are used in many industries, it is less likely that higher biomarker levels of arsenic metabolites in white collars are due to occupational exposures. Although, health care workers may be exposed to mercury via medical or dental equipment (Trzcinka-Ochocka et al., 2007). Higher mercury and arsenic biomarker levels among these white-collar workers may indicate higher fish consumption (Shimshack et al., 2007), which associated with higher socioeconomic status instead of an indicator of occupational exposures (Tyrrell et al., 2013). It is also doubtful that white-collar workers are manufacturing products containing exposed to BP-3 (Benzophenone, 2013; Program, 2006). Instead, as BP-3 is used to prevent UV light from damaging scents and colors in personal care products (CDC, 2017), it more likely that higher levels of this chemical may suggest that cosmetics usage has a major role in strategic self-presentation. Overall, the chemical exposure profiles of white-collar workers likely indicate behaviors associated with socioeconomic status. Many studies using NHANES have been limited to studying chemical co-exposures in one chemical family due to the non-random sparsity of the chemical biomarker data (Sadetzki et al., 2016; Shim et al., 2017). To address this sparsity challenge, we conducted clustering analysis on exposure differences among the occupational groups, i.e. clustering analysis on statistics of the biomarker data instead of on the raw data. Our framework enabled the identification of co-exposure across a wide range of chemicals not only limited to one chemical family. This framework can be applied in other settings to help cluster observations based on similar profiles especially in a non-randomly sparse dataset. This can be done without having to form a complete dataset or impute the missing values.

Chemical exposures have been implicated as etiological agents in adverse noncancer effects on the nervous, reproductive, immune, and cardiovascular systems. Such toxic chemicals include heavy metals such as lead and cadmium, VOCs, PAHs, phthalates, and smoking-related compounds. Several epidemiological and toxicological studies have implicated the neurotoxicity of heavy metals, especially lead (Garza et al., 2006) and cadmium (Méndez-Armenta & Ríos, 2007). Many studies have shown the reproductive toxicity of phthalates (Martino-Andrade & Chahoud, 2010) and PAHs (Yin et al., 2017). PAHs (Schober et al., 2007) and VOCs such as toluene and benzene (Jiménez-Garza et al., 2017) are also known to elicit an inflammatory response individually and in combination (Kuang et al., 2020). Heavy metals have been recently linked to cardiovascular disease (Duan et al., 2020). Tobacco exposures have been causal factors in several noncancer effects such as respiratory problems (Anderson & Ferris Jr, 1962), heart disease (Lakier, 1992), infections (Arcavi & Benowitz, 2004), and fertility problems (Stillman et al., 1986). Overall, our findings of toxicant levels in blue collar workers exceeding acceptable levels coupled with existing literature on the adverse effects of such toxicants continues to further implicate blue collars from the aforementioned industries to be a high-risk population.

Our findings lend urgency to understand how one’s occupation can be a route for smoking initiation in young people and consequentially exposure to other toxicants. The connection between being a blue collar, active smoker, and younger may suggest that being an active smoker is part of the culture of a blue-collar worker (Ham et al., 2011). Furthermore, blue-collar workers are additionally exposed to VOCs, PAHs, and heavy metals, since these chemicals have been detected in tobacco products. Interestingly, blue-collar workers have higher levels of nicotine metabolites and dietary components such as beta- and alpha-carotenes. This finding is especially alarming, since cancer-preventative trials have shown that vitamin A analogues, alone or in combination with vitamin E, are risk factors for lung cancer and mortality in active smokers (Omenn et al., 1996; Paolini et al., 1999). Our findings can inspire future studies to develop interventions to understand cultural and behavioral factors leading to smoking initialization and implement evidence-based regulations on tobacco control to prevent the younger population from initiating smoking (Kunst, 2021).

Environmental injustice is defined as the disproportionate exposures of toxicants and their consequential effects on health to disadvantaged groups such as individuals from racial minorities and/or low socioeconomic status (Maantay, 2002). Blue-collar workers are disproportionally exposed to some of the most toxic chemicals with several found at concentration exceeding acceptable guidelines for cancer and noncancer effects (Kolonel, 1976; Swartz, 2001). Furthermore, many blue-collar workers may come from the lowest socioeconomic status (Kivimäki et al., 2006). Thus, our findings call for an increase in exposure surveillance and industrial controls, effect regulations on chemical exposures, inform remediation strategies, and help implement interventions programs to improve the health of workers susceptible to toxicant exposures. Moreover, health providers such as occupational physicians and industrial health personnel can use our findings to inform their patients on preventatives measures to avoid occupations and toxicant exposures that may increase their personal and familial disease risk.

The present study has several other limitations. First, as NHANES is a cross-sectional survey, we cannot make claims on causal factors of chemical exposures. Second, a limitation of using chemical biomarker data is that a delay between the time of exposure and time of data collection may prevent the detection of higher occupational exposures. This limitation is especially salient for VOCs, which have short half-life ranging from 2 to 128 hours (OECD, 2004, 2014), which implies that substantially higher biomarker levels could be observed at workplace. Third, while we identified differences in chemical biomarker levels by occupation, we cannot claim that such differences are due to occupational exposures, since we do not know the source of all exposures for each study participant. Nevertheless, our findings can inspire future studies to prioritize chemicals and susceptible occupations in specific industries to measure at the workplace. Fourth, while NHANES has biomonitoring data on 517 chemicals, we only obtained biomonitoring equivalents for 106 chemicals (20.5%). Fifth, there is potential uncertainty in how the biomonitoring equivalents are converted from exposures values to biomarker concentrations. Such uncertainty can either decrease or increase the value of the biomonitoring equivalent and in turn change the interpretation of which chemicals for which occupations should be further monitored.

## 5. Conclusions

Evaluating differences in chemical exposures by occupation is essential to identify occupations susceptible to high exposures to toxicants as well as understand how occupational exposures play a role in adverse health outcomes. We applied an unbiased approach to screen across 129 chemical biomarkers to characterize the chemical exposure profiles across white- and blue-collar workers from 20 different industries and 7 unemployed groups. We developed a framework using hierarchical clustering on differences of chemical biomarker levels to identify clusters of occupations with similar chemical exposures. Our framework enabled 1) comprehensive characterization of chemical exposures across a wide variety of occupations and toxicants and 2) identification of occupations susceptible to high toxicant exposure and exceeding acceptable health-based levels. These findings can guide efforts to design targeted interventions to reduce and prevent exposures in susceptible occupations and help mitigate negative effects from toxicant exposures.

## Data Availability

All code and data produced for this manuscript are available online at https://github.com/vynguyen92/nhanes_occupational_exposures. All data produced are available as an interactive app at https://chiragjp.shinyapps.io/nhanes_occupational_exposures/.

https://chiragjp.shinyapps.io/nhanes_occupational_exposures/

## CRediT authorship contribution statement

VKN: conceptualisation, data curation, formal analysis, software, visualization, writing – original draft, writing – review & editing, verification of underlying data

JC: conceptualisation, funding acquisition, resources, writing – original draft, writing – review & editing

CJP: funding acquisition, resources, writing – original draft, writing – review & editing

MS: writing – original draft, writing – review & editing

OJ: conceptualisation, resources, writing – original draft, writing – review & editing

## Data Sharing

Data for the study was collected by the Centers of Disease Control and Prevention and is publicly available at https://www.n.cdc.gov/nchs/nhanes/Default.aspx. Data includes individual participant data with corresponding demographics, occupational codes, and measurements for chemical biomarkers. A data dictionary defining each field in the set is publicly available at https://www.n.cdc.gov/nchs/nhanes/Search/variablelist.aspx?Component=Demographics, https://www.n.cdc.gov/nchs/nhanes/search/datapage.aspx?Component=Questionnaire, and https://www.n.cdc.gov/nchs/nhanes/Search/variablelist.aspx?Component=Laboratory. Our curated data along with related documents such as excel documents and R code for the statistical analysis will be available at https://github.com/vynguyen92/nhanes_occupational_exposures with publication. Data will be available to the requester via email after the approval of the request.

