## Supplemental material for "Identification of occupations susceptible to high exposure and risk associated with multiple toxicants in an observational study: National Health and Nutrition Examination Survey 1999-2014"

**Formatted:** Position: Horizontal: Right, Relative to: Margin, Vertical: 0", Relative to: Paragraph, Wrap Around

**Formatted:** Position: Horizontal: Right, Relative to: Margin, Vertical: 0", Relative to: Paragraph, Wrap Around

**Text S1. Exclusion criteria on chemical biomarkers**

We preferred lipid adjusted measurements for biomarkers indicated by 7- or 8-letter NHANES codename ending in "L" or "LA," respectively, for which NHANES provided both lipid-adjusted and non-lipid adjusted measurements. Therefore, we excluded non-lipid adjusted chemical biomarkers for which there was a corresponding lipid-adjusted measure (c = 68). We excluded creatinine as it is not an exposure biomarker (c = 1). We calculated detection frequencies for each combination of chemical biomarkers and occupations. For a given chemical, we calculated the average detection frequency across the occupational groups and excluded chemicals with detection frequency below 10% overall (c = 133). Then for each remaining chemical, we calculated the sample size for each sector-collar combination, determined the median sample size across all sector-collars, and excluded chemical biomarkers with a median sample size of less than 90 participants (c = 184). We chose 90 participants as the cut-off based on a power calculation to have a 70% chance of detecting a significant difference with a p-value of less than 5% and with an effect size of 0.37 (Table S3).

**Formatted:** Position: Horizontal: Right, Relative to: Margin, Vertical: 0", Relative to: Paragraph, Wrap Around

**Text S2. Hierarchal clustering procedure to identify groups of occupations with similar chemical profile**

First, we normalized the percent differences across a given chemical biomarker with the mean equal to 0 and the standard deviation equal to 1 to make the chemicals comparable to each other. Normalizing also prevents the chemical biomarker(s) with the greatest variation from driving the clustering [21]. Second, we used Pearson's correlation-based distance as our distance measure [22,23]. This distance measure calculates the similarity between the chemicals of every pair of occupational groups by using Pearson's correlation. We preferred Pearson's correlation-based distance to Euclidean distance as we are interested in identifying clusters of occupations with similar overall profiles [21] in chemical exposures. Using Euclidean distance would cluster the occupation groups based on magnitude of the percent differences, e.g. occupations with the highest chemical biomarker levels would cluster together. Third, we needed to decide which linkage method to use in order to best determine how the occupational groups should cluster together based on similarity in chemical exposures [21]. We tested four different linkage functions: single, complete, average, and McQuitty's linkage method. We provided description of each linkage method in **Table S8**. Fourth, we selected the linkage method that generated the dendrogram that best preserves the dissimilarity among the occupation groups. We evaluated this performance with the cophenetic correlation coefficient, which is a correlation coefficient between the original distance matrix and the distance matrix generated by the clustering configuration [24]. A cophenetic correlation coefficient equal to 1 implies that the clustering configuration perfectly preserves the dissimilarity between the objects, e.g. occupation groups. The average linkage method has the highest cophenetic coefficients (**Figure S3**), so we report the clustering results from the average linkage method. Finally, we conducted hierarchical agglomerative clustering with 1500 multiscale bootstrap replicates to calculate the approximately unbiased (AU) p-values [25]. An AU p-value of 1 implies that all bootstrap replicates support the cluster, implying that the cluster is strongly supported by the data [25]. We visualized the clusters of occupation groups with dendrograms. We also used Pearson's correlation-based distance to cluster the chemical biomarkers based on having similar profiles on occupational groups.

**Formatted:** Position: Horizontal: Right, Relative to: Margin, Vertical: 0", Relative to: Paragraph, Wrap Around

**Text S3. Sensitivity Analysis on Contribution of Occupation in Explaining Chemical Biomarker Levels**

We examined how occupation contributes to explaining chemical biomarker levels. To quantify how occupation explains variation in chemical biomarker levels, we first determined the Pearson's R2 for univariate linear models with the outcome variable as the chemical biomarker levels, treated as a continuous variable, and the main predictor as the occupational groups. In addition, we calculated how much of the variation of chemical biomarker levels is additionally explained by occupation after controlling for all other covariates.

Figure S15 in appendix p 61 shows the variation of chemical biomarker levels explained by occupation alone and after adjusting for all covariates. The chemical biomarker levels are treated as a continuous variable. The "1"s show the how much of the variation of chemical biomarker levels is explained by occupation alone, i.e., the R2 of the univariate model with the outcome variable as the chemical biomarker levels and the main predictor as the occupational groups. The R2s of the univariate models range from 0.58% to 14.3%. Blood lead (14.3%), NNAL (11.1%), cotinine (9.6%), 3-fluorene (8.5%), and trans-b-carotene (8.4%) show the highest variations of chemical biomarker levels explained by occupation alone. The "2"s show the contribution of occupation on explaining chemical biomarker levels changes after adjusting for all other covariates, i.e., the difference in R2 between the fully adjusted models and the models not adjusting for occupation. The R2s, showing the additional contribution of occupation, ranges from 0.28% to 4.8%. Chemicals showing the highest, additional contribution of occupation include cotinine (4.8%), d-Tocopherol (3.3%), Desethyl hydroxy DEET (3.0%), Combined Lutein/zeaxanthin (2.4%), and a-carotene (2.4%). There are substantial differences between the contribution of occupation alone versus contribution of occupation independent of other confounders in explaining chemical biomarker levels. Such differences suggest the complex correlation structure among the covariates and occupation in explaining the biomarker levels.

**Formatted:** Position: Horizontal: Right, Relative to: Margin, Vertical: 0", Relative to: Paragraph, Wrap Around

**Text S4. Sensitivity Analysis on the Influence of Smoking**

We performed several sensitivity analyses to examine the influence of tobacco exposures on characterizing occupational differences in chemical biomarker levels. We have interest in tobacco exposures as we observed the signals for cotinine were the most substantial and significant. First, we observed how smoking contributes to explaining the variance of the chemical biomarker levels. We, therefore, calculated the difference in R2 between the fully adjusted models and the models not adjusting for tobacco exposures. The chemical biomarkers with the highest difference in R2 may suggest that exposures to these chemicals could be due, at least in part, to smoking. Second, to determine the degree to which tobacco exposures was driving the clustering of the occupations, we excluded the regression results of biomarkers of tobacco exposures such as cotinine and NNAL from the clustering analyses. Third, we performed analyses to understand the extent to which similarity in chemical exposure profiles among the blue-collar workers were due to these workers being predominantly active smokers. To do this, we characterized the chemical exposure profiles for those who were not actively smoking, by performing additional regression and clustering analyses after excluding active smokers. Active smokers were defined with serum cotinine levels above 3 ng/mL26. The sample size after excluding active smokers is 30,545 participants.

Figure S16 shows how smoking contributes to the explaining variation in chemical biomarker levels, i.e difference in R2 between the fully adjusted models and the models not adjusting for smoking. The differences in R2 ranges from 1.4e-7% to 46.9%. Smoking explains a substantial amount of the variation for VOCs, PAHs, smoking related compounds, and blood cadmium, suggesting that participants may be exposed to such toxicants via smoking.

Figure S17 is a heatmap showing relative differences in chemical biomarker profiles across occupational groups without including cotinine and NNAL as part of the clustering analysis to form the dendrogram on the occupational groups. The dendrogram in this figure is similar to the dendrogram when smoking related compounds were included in the clustering analysis of the occupational groups. This suggests that the smoking related compounds are not solely driving the clustering of the occupational groups.

Figure S18 is a panel of correlation plots comparing the relative difference in chemical biomarker levels when including active smokers versus excluding active smokers. Each point is for comparing the relative difference in chemical biomarkers between a given occupational group and the reference group of white collars from “Public Administration”. The chemicals and occupational groups that are affected by the exclusion of active smokers (i.e. the points that deviate from the 1-1 line) include VOCs, PAHs, and smoking related compounds in blue-collars and unemployed participants.

**Formatted:** Position: Horizontal: Right, Relative to: Margin, Vertical: 0", Relative to: Paragraph, Wrap Around

Tables

Table S1. Job occupation description with corresponding collar category. OCD240 was used in Cycle 1-3, while OCD241 was used in Cycle 4-8.

| NHANES codename | Original occupational code | Original occupation description | Harmonized occupational code | Harmonized occupation description | Collar category |
|---|---|---|---|---|---|
| OCD240 | 1 | Executive, administrators, and managers | 1 | Management Occupations | White |
| OCD240 | 2 | Management related occupations | 1 | Management Occupations | White |
| OCD240 | 5 | Teachers | 8 | Education, Training, Library Occupations | White |
| OCD240 | 6 | Writers, artists, entertainers, and athletes | 9 | Arts, Design, Entertainment, Sports, Media Occupations | White |
| OCD240 | 4 | Health diagnosing, assessing and treating occupations | 10 | Healthcare Practitioner, Technical Occupations | White |
| OCD240 | 22 | Health service occupations | 10 | Healthcare Practitioner, Technical Occupations | White |
| OCD240 | 18 | Protective service occupations | 12 | Protective Service Occupations | White |
| OCD240 | 20 | Cooks | 13 | Food Preparation, Serving Occupations | Blue |
| OCD240 | 21 | Miscellaneous food preparation and service occupations' | 13 | Food Preparation, Serving Occupations | Blue |
| OCD240 | 19 | Waiters and waitresses | 13 | Food Preparation, Serving Occupations | Blue |
| OCD240 | 23 | Cleaning and building service occupations | 14 | Building & Grounds Cleaning, Maintenance Occupations | Blue |
| OCD240 | 17 | Private household occupations | 14 | Building & Grounds Cleaning, Maintenance Occupations | Blue |
| OCD240 | 24 | Personal service occupations | 15 | Personal Care, Service Occupations | Blue |
| OCD240 | 10 | Sales representatives, finance, business, & commodities ex. Retail | 16 | Sales & Related Occupations | White |
| OCD240 | 11 | Sales workers, retail and personal services | 16 | Sales & Related Occupations | White |
| OCD240 | 9 | Supervisors and proprietors, sales occupations | 16 | Sales & Related Occupations | White |
| OCD240 | 13 | Information clerks | 17 | Office, Administrative Support Occupations | White |
| OCD240 | 15 | Material recording, scheduling, and distributing clerks | 17 | Office, Administrative Support Occupations | White |
| OCD240 | 16 | Miscellaneous administrative support occupations | 17 | Office, Administrative Support Occupations | White |
| OCD240 | 14 | Records processing occupations | 17 | Office, Administrative Support Occupations | White |
| OCD240 | 12 | Secretaries, stenographers, and typists | 17 | Office, Administrative Support Occupations | White |

**Formatted:** Position: Horizontal: Right, Relative to: Margin, Vertical: 0", Relative to: Paragraph, Wrap Around

| | | | | | |
|---|---|---|---|---|---|
| OCD240 | 26 | Farm and nursery workers | 18 | Farming, Fishing, Forestry Occupations | Blue |
| OCD240 | 25 | Farm operators, managers, and supervisors | 18 | Farming, Fishing, Forestry Occupations | Blue |
| OCD240 | 27 | Related agricultural, forestry, and fishing occupations | 18 | Farming, Fishing, Forestry Occupations | Blue |
| OCD240 | 37 | Construction laborers | 19 | Construction, Extraction Occupations | Blue |
| OCD240 | 30 | Construction trades | 19 | Construction, Extraction Occupations | Blue |
| OCD240 | 31 | Extractive and precision production occupations | 19 | Construction, Extraction Occupations | Blue |
| OCD240 | 38 | Laborers, except construction | 19 | Construction, Extraction Occupations | Blue |
| OCD240 | 29 | Other mechanics and repairers | 20 | Installation, Maintenance, Repair Occupations | Blue |
| OCD240 | 28 | Vehicle and mobile equipment mechanics and repairers | 20 | Installation, Maintenance, Repair Occupations | Blue |
| OCD240 | 34 | Fabricators, assemblers, inspectors, and samplers | 21 | Production Occupations | Blue |
| OCD240 | 33 | Machine operators, assorted materials | 21 | Production Occupations | Blue |
| OCD240 | 32 | Textile, apparel, and furnishings machine operators | 21 | Production Occupations | Blue |
| OCD240 | 39 | Freight, stock, and material movers, hand | 22 | Transportation, Material Moving Occupations | Blue |
| OCD240 | 35 | Motor vehicle operators | 22 | Transportation, Material Moving Occupations | Blue |
| OCD240 | 40 | Other helpers, equipment cleaners, hand packagers and laborers | 22 | Transportation, Material Moving Occupations | Blue |
| OCD240 | 36 | Other transportation and material moving occupations | 22 | Transportation, Material Moving Occupations | Blue |
| OCD240 | 41 | Military occupations | 23 | Armed Forces | Blue |
| OCD240 | 98 | Blank but applicable | 99 | Blank but applicable | |
| OCD240 | 8 | Technicians and related support occupations | 3,4,5,6,7 | STEM, Social, and Legal Services Occupations | White |
| OCD240 | 3 | Engineers, architects and scientists | 3,4,5,6,7 | STEM, Social, and Legal Services Occupations | White |
| OCD240 | 7 | Other professional specialty occupations | 3,4,5,6,7 | STEM, Social, and Legal Services Occupations | White |
| OCD241 | 1 | Management Occupations | 1 | Management Occupations | White |
| OCD241 | 2 | Business, Financial Operations Occupations | 2 | Business, Financial Operations Occupations | White |
| OCD241 | 8 | Education, Training, Library Occupations | 8 | Education, Training, Library Occupations | White |
| OCD241 | 9 | Arts, Design, Entertainment, Sports, Media Occupations | 9 | Arts, Design, Entertainment, Sports, Media Occupations | White |
| OCD241 | 10 | Healthcare Practitioner, Technical Occupations | 10 | Healthcare Practitioner, Technical Occupations | White |

**Formatted:** Position: Horizontal: Right, Relative to: Margin, Vertical: 0", Relative to: Paragraph, Wrap Around

| | | | | | |
|---|---|---|---|---|---|
| OCD241 | 11 | Healthcare Support Occupations | 11 | Healthcare Support Occupations | White |
| OCD241 | 12 | Protective Service Occupations | 12 | Protective Service Occupations | White |
| OCD241 | 13 | Food Preparation, Serving Occupations | 13 | Food Preparation, Serving Occupations | Blue |
| OCD241 | 14 | Building & Grounds Cleaning, Maintenance Occupations | 14 | Building & Grounds Cleaning, Maintenance Occupations | Blue |
| OCD241 | 15 | Personal Care, Service Occupations | 15 | Personal Care, Service Occupations | Blue |
| OCD241 | 16 | Sales & Related Occupations | 16 | Sales & Related Occupations | White |
| OCD241 | 17 | Office, Administrative Support Occupations | 17 | Office, Administrative Support Occupations | White |
| OCD241 | 18 | Farming, Fishing, Forestry Occupations | 18 | Farming, Fishing, Forestry Occupations | Blue |
| OCD241 | 19 | Construction, Extraction Occupations | 19 | Construction, Extraction Occupations | Blue |
| OCD241 | 20 | Installation, Maintenance, Repair Occupations | 20 | Installation, Maintenance, Repair Occupations | Blue |
| OCD241 | 21 | Production Occupations | 21 | Production Occupations | Blue |
| OCD241 | 22 | Transportation, Material Moving Occupations | 22 | Transportation, Material Moving Occupations | Blue |
| OCD241 | 23 | Armed Forces | 23 | Armed Forces | Blue |
| OCD241 | 98 | Text present but uncodable | 98 | Text present but uncodable | |
| OCD241 | 99 | Blank but applicable | 99 | Blank but applicable | |
| OCD241 | 4 | Architecture, Engineering Occupations | 3,4,5,6,7 | STEM, Social, and Legal Services Occupations | White |
| OCD241 | 6 | Community, Social Services Occupations | 3,4,5,6,7 | STEM, Social, and Legal Services Occupations | White |
| OCD241 | 3 | Computer, Mathematical Occupations | 3,4,5,6,7 | STEM, Social, and Legal Services Occupations | White |
| OCD241 | 7 | Legal Occupations | 3,4,5,6,7 | STEM, Social, and Legal Services Occupations | White |
| OCD241 | 5 | Life, Physical, Social Science Occupations | 3,4,5,6,7 | STEM, Social, and Legal Services Occupations | White |

Table S2. Number of participants by occupation group. These sector-occupation groups include sector-collar combinations and unemployment status (e.g. Looking for work, On layoff, Disabled, Going to school, Taking care of house or family, Retired). "Occupation Missing" consist of participants who are at least 16 years old but did not record their occupation status.

| Occupation Group | Number of Participants | Include? |
|---|---|---|
| Blank but applicable - White | 4 | no |
| Blank but applicable - Blue | 5 | no |
| Finance, Insurance - Blue | 5 | no |
| Armed Forces - White | 8 | no |
| Armed Forces - Blue | 16 | no |
| Private Households - White | 19 | no |
| Mining - White | 39 | no |
| Public Administration - Blue | 77 | yes |
| Utilities - White | 93 | yes |
| Information Services - Blue | 95 | yes |
| Mining - Blue | 98 | yes |
| Utilities - Blue | 128 | yes |
| Agriculture, Forestry, Fishing - White | 142 | yes |
| Real Estate, Rental, Leasing - Blue | 152 | yes |
| Arts, Entertainment, Recreation - Blue | 280 | yes |
| Private Households - Blue | 286 | yes |
| Professional, Scientific, Technical Services - Blue | 288 | yes |
| Wholesale Trade - Blue | 312 | yes |
| Arts, Entertainment, Recreation - White | 315 | yes |
| Construction - White | 322 | yes |
| Education Services - Blue | 345 | yes |
| Transportation, Warehousing - White | 370 | yes |
| Accommodation, Food Services - White | 381 | yes |
| Manufacturing: Non-Durable Goods - White | 398 | yes |
| Wholesale Trade - White | 401 | yes |
| Information Services - White | 443 | yes |
| Agriculture, Forestry, Fishing - Blue | 493 | yes |
| Real Estate, Rental, Leasing - White | 498 | yes |
| Other Services - Blue | 513 | yes |
| On layoff | 565 | yes |
| Management, Administrative, Waste Services - White | 625 | yes |
| Manufacturing: Durable Goods - White | 640 | yes |
| Transportation, Warehousing - Blue | 682 | yes |
| Health Care, Social Assistance - Blue | 697 | yes |
| Other Services - White | 720 | yes |

**Formatted:** Position: Horizontal: Right, Relative to: Margin, Vertical: 0", Relative to: Paragraph, Wrap Around

| | | |
|---|---|---|
| Management, Administrative, Waste Services - Blue | 793 | yes |
| Finance, Insurance - White | 832 | yes |
| Manufacturing: Non-Durable Goods - Blue | 884 | yes |
| Public Administration - White | 889 | yes |
| Professional, Scientific, Technical Services - White | 899 | yes |
| Manufacturing: Durable Goods - Blue | 1086 | yes |
| Accommodation, Food Services - Blue | 1293 | yes |
| Retail Trade - Blue | 1323 | yes |
| Looking for work | 1559 | yes |
| Education Services - White | 1669 | yes |
| Construction - Blue | 1700 | yes |
| Unable to work for health reasons | 1708 | yes |
| Occupation Missing | 1766 | yes |
| Disabled | 2400 | yes |
| Retail Trade - White | 2503 | yes |
| Health Care, Social Assistance - White | 2705 | yes |
| Going to school | 3680 | yes |
| Taking care of house or family | 3699 | yes |
| Retired | 9261 | yes |

**Formatted:** Position: Horizontal: Right, Relative to: Margin, Vertical: 0", Relative to: Paragraph, Wrap Around

Table S3. Sample size required to detect significant differences. x% power mean that a study has a x% chance of ending up with a p-value of less than 5% in a statistical test when the effect size (or regression coefficient) is a corresponding value if the sample size is a particular number. For example, a study has a 70% chance of detecting a significance difference with a regression coefficient of 0.37 when the sample size is 90 participants.

| Effect Size (or Regression Coefficients) | 90% Power | 80% Power | 70% Power |
|---|---|---|---|
| 0.01 | 209952 | 156800 | 123008 |
| 0.02 | 52488 | 39200 | 30752 |
| 0.03 | 23328 | 17422 | 13668 |
| 0.04 | 13122 | 9800 | 7688 |
| 0.05 | 8398 | 6272 | 4920 |
| 0.06 | 5832 | 4356 | 3417 |
| 0.07 | 4285 | 3200 | 2510 |
| 0.08 | 3281 | 2450 | 1922 |
| 0.09 | 2592 | 1936 | 1519 |
| 0.1 | 2100 | 1568 | 1230 |
| 0.11 | 1735 | 1296 | 1017 |
| 0.2 | 525 | 392 | 308 |
| 0.3 | 233 | 174 | 137 |
| 0.31 | 218 | 163 | 128 |
| 0.32 | 205 | 153 | 120 |
| 0.33 | 193 | 144 | 113 |
| 0.34 | 182 | 136 | 106 |
| 0.35 | 171 | 128 | 100 |
| 0.36 | 162 | 121 | 95 |
| 0.37 | 153 | 115 | 90 |
| 0.38 | 145 | 109 | 85 |
| 0.39 | 138 | 103 | 81 |
| 0.4 | 131 | 98 | 77 |
| 0.5 | 84 | 63 | 49 |
| 0.6 | 58 | 44 | 34 |
| 0.7 | 43 | 32 | 25 |
| 0.8 | 33 | 25 | 19 |
| 0.9 | 26 | 19 | 15 |
| 1 | 21 | 16 | 12 |

**Formatted:** Position: Horizontal: Right, Relative to: Margin, Vertical: 0", Relative to: Paragraph, Wrap Around

**Table S4. Inclusion criteria for 517 chemical biomarkers.**

| Chemical name | Detection frequency | Median sample size across occupational groups | Include? |
|---|---|---|---|
| Total Calcium | NA | 561 | yes |
| Chloride | NA | 561 | yes |
| Potassium | NA | 561 | yes |
| Sodium | NA | 561 | yes |
| Vitamin B12, serum | NA | 348 | yes |
| 4-pyridoxic acid | NA | 203 | yes |
| Vitamin B6 | NA | 202 | yes |
| Vitamin A | NA | 187 | yes |
| d-Tocopherol | NA | 186 | yes |
| g-tocopherol | NA | 178 | yes |
| Retinyl palmitate | NA | 172 | yes |
| Retinyl stearate | NA | 150 | yes |
| a-Carotene | NA | 118 | yes |
| trans-b-carotene | NA | 118 | yes |
| Combined Lutein/zeaxanthin | NA | 118 | yes |
| trans-lycopene | NA | 118 | yes |
| b-cryptoxanthin | NA | 117 | yes |
| total Lycopene | NA | 117 | yes |
| cis-b-carotene | NA | 114 | yes |
| Vitamin D | 100.00 | 454 | yes |
| 25-hydroxyvitamin D3 | 100.00 | 238 | yes |
| Iodine, urine | 100.00 | 207 | yes |
| Methylmalonic Acid | 100.00 | 119 | yes |
| 2-fluorene | 99.99 | 174 | yes |
| 2-naphthol | 99.99 | 143 | yes |
| Perchlorate, urine | 99.99 | 246 | yes |
| 9-fluorene | 99.99 | 120 | yes |
| Cesium, urine | 99.95 | 199 | yes |
| Molybdenum, urine | 99.94 | 197 | yes |
| 1-naphthol | 99.93 | 141 | yes |
| Mono-ethyl phthalate | 99.88 | 184 | yes |
| Urinary thiocyanate | 99.88 | 227 | yes |
| Vitamin C | 99.87 | 122 | yes |
| Mono-2-ethyl-5-carboxypentyl phthalate | 99.86 | 139 | yes |

**Formatted:** Position: Horizontal: Right, Relative to: Margin, Vertical: 0", Relative to: Paragraph, Wrap Around

| | | | |
|---|---|---|---|
| Sum of metabolites of DEHP | 99.85 | 139 | yes |
| Urinary nitrate | 99.75 | 227 | yes |
| 1-phenanthrene | 99.71 | 175 | yes |
| Perfluorooctane sulfonic acid | 99.69 | 154 | yes |
| Daidzein | 99.61 | 118 | yes |
| Acrylamide | 99.60 | 132 | yes |
| Perfluorooctanoic acid | 99.55 | 154 | yes |
| Enterolactone | 99.50 | 118 | yes |
| Blood lead | 99.47 | 522 | yes |
| Methyl paraben | 99.33 | 109 | yes |
| Thallium, urine | 99.32 | 198 | yes |
| Mono-(2-ethyl-5-hydroxyhexyl) phthalate | 99.26 | 162 | yes |
| Urinary total Arsenic | 98.95 | 146 | yes |
| 3-fluorene | 98.89 | 172 | yes |
| 3-phenanthrene | 98.88 | 146 | yes |
| Mono-(2-ethyl-5-oxohexyl) phthalate | 98.62 | 162 | yes |
| 2-phenanthrene | 98.57 | 145 | yes |
| Cobalt, urine | 98.53 | 199 | yes |
| Genistein | 98.43 | 123 | yes |
| Glycideamide | 98.38 | 128 | yes |
| Mono-n-butyl phthalate | 98.31 | 184 | yes |
| 2,5-dichlorophenol | 98.23 | 139 | yes |
| Mono(carboxyoctyl) phthalate | 98.15 | 109 | yes |
| Mono-benzyl phthalate | 98.12 | 184 | yes |
| Barium, urine | 98.04 | 196 | yes |
| Urinary Benzophenone-3 | 96.64 | 139 | yes |
| Lead, urine | 96.01 | 199 | yes |
| Mono-(3-carboxypropyl) phthalate | 95.28 | 162 | yes |
| 1-pyrene | 95.19 | 139 | yes |
| Mono(carboxynonyl) phthalate | 95.05 | 109 | yes |
| Propyl paraben | 94.66 | 109 | yes |
| Blood Toluene | 94.63 | 153 | yes |
| Mono-isobutyl phthalate | 94.52 | 162 | yes |
| Cadmium, urine | 93.48 | 197 | yes |
| Urinary Bisphenol A | 92.09 | 139 | yes |
| Mercury, urine | 91.14 | 192 | yes |
| Blood Chloroform | 90.95 | 159 | yes |
| 2,4-dichlorophenol | 88.95 | 139 | yes |
| Blood mercury, total | 88.82 | 416 | yes |
| 3,5,6-trichloropyridinol | 87.94 | 120 | yes |

**Formatted:** Position: Horizontal: Right, Relative to: Margin, Vertical: 0", Relative to: Paragraph, Wrap Around

| | | | |
|---|---|---|---|
| o-Desmethylangolensin (O-DMA) | 86.18 | 145 | yes |
| Blood m-/p-Xylene | 85.27 | 203 | yes |
| Paranitrophenol | 83.16 | 125 | yes |
| Perfluorononanoic acid | 82.84 | 154 | yes |
| Urinary Dimethylarsonic acid | 82.68 | 147 | yes |
| Uranium, urine | 82.56 | 162 | yes |
| Perfluorohexane sulfonic acid | 82.52 | 154 | yes |
| Tungsten, urine | 82.43 | 199 | yes |
| DEET acid | 81.59 | 93 | yes |
| Sum of arsenic metabolites | 80.05 | 147 | yes |
| Blood cadmium | 79.20 | 522 | yes |
| 3-phenoxybenzoic acid | 78.78 | 140 | yes |
| Antimony, urine | 77.11 | 198 | yes |
| epi-25-hydroxyvitamin D3 | 76.95 | 229 | yes |
| Urinary Triclosan | 76.26 | 139 | yes |
| Cotinine | 76.08 | 561 | yes |
| Blood Bromodichloromethane | 75.51 | 170 | yes |
| Dimethyl thiophosphate | 72.44 | 127 | yes |
| Mono-(2-ethylhexyl) phthalate | 71.53 | 184 | yes |
| 2-(N-methyl-PFOSA) acetate | 66.52 | 133 | yes |
| NNAL , urine | 61.47 | 247 | yes |
| Urinary Arsenobetaine | 61.40 | 147 | yes |
| 2,4-D | 57.26 | 164 | yes |
| Perfluorodecanoic acid | 56.05 | 154 | yes |
| Mono-n-methyl phthalate | 55.29 | 134 | yes |
| Diethylthiophosphate | 54.03 | 126 | yes |
| Diethylphosphate | 53.55 | 125 | yes |
| Dimethyl phosphate | 53.42 | 125 | yes |
| Blood 1,4-Dichlorobenzene | 52.71 | 140 | yes |
| Ethyl paraben | 49.34 | 109 | yes |
| Blood Ethylbenzene | 47.60 | 204 | yes |
| Blood Dibromochloromethane | 46.94 | 194 | yes |
| Blood o-Xylene | 46.17 | 200 | yes |
| Blood Styrene | 43.79 | 114 | yes |
| Urinary Monomethylarsonic acid | 40.10 | 147 | yes |
| Butyl paraben | 38.86 | 109 | yes |
| Blood Benzene | 38.47 | 204 | yes |
| Blood MTBE | 37.98 | 193 | yes |
| Dimethyl dithiophosphate | 35.34 | 127 | yes |
| Perfluoroundecanoic acid | 33.11 | 154 | yes |

**Formatted:** Position: Horizontal: Right, Relative to: Margin, Vertical: 0", Relative to: Paragraph, Wrap Around

| | | | |
|---|---|---|---|
| 2,4,5-trichlorophenol | 31.90 | 95 | yes |
| 2,4,6-trichlorophenol | 28.68 | 95 | yes |
| O-Phenylphenol | 25.91 | 95 | yes |
| Blood Bromoform | 24.21 | 176 | yes |
| Mono-isononyl phthalate | 22.24 | 184 | yes |
| Mercury, Inorganic | 20.82 | 416 | yes |
| Perfluorooctane sulfonamide | 20.81 | 133 | yes |
| Blood 2,5-Dimethylfuran | 20.40 | 181 | yes |
| 25-hydroxyvitamin D2 | 19.89 | 238 | yes |
| Urinary Arsenous acid | 19.80 | 147 | yes |
| trans-3-(2,2-dichlorovinyl)-2,2-dimethylcyclopropane carboxylic acid | 19.48 | 142 | yes |
| Oxypyrimidine | 18.36 | 128 | yes |
| 2-(N-ethyl-PFOSA) acetate | 15.87 | 133 | yes |
| Blood Tetrachloroethene | 15.83 | 206 | yes |
| Blood furan | 14.73 | 115 | yes |
| Diethyl dithiophosphate | 12.05 | 126 | yes |
| Desethyl hydroxy DEET | 11.34 | 92 | yes |
| Creatinine, urine | NA | 584 | no |
| a-Cryptoxanthin | NA | 62 | no |
| a-Tocopherol | NA | 62 | no |
| total b-Carotene | NA | 62 | no |
| cis-Lycopene | NA | 62 | no |
| cis-Lutein/Zeaxanthin | NA | 62 | no |
| Lutein | NA | 62 | no |
| Phytoene | NA | 62 | no |
| Phytofluene | NA | 62 | no |
| Zeaxanthin | NA | 62 | no |
| Serum Copper | NA | 39 | no |
| Serum Zinc | NA | 39 | no |
| Melamine | NA | 5.5 | no |
| Cyanuric acid | NA | 4 | no |
| Blood manganese | 100.00 | 86 | no |
| Selenium | 100.00 | 86 | no |
| Blood Nitromethane | 100.00 | 85 | no |
| Formaldehyde | 100.00 | 18 | no |
| PCB28 Lipid Adj | 100.00 | 17 | no |
| PCB52 Lipid Adj | 100.00 | 17 | no |
| PCB74 Lipid Adj | 100.00 | 17 | no |
| PCB118 Lipid Adj | 100.00 | 17 | no |

**Formatted:** Position: Horizontal: Right, Relative to: Margin, Vertical: 0", Relative to: Paragraph, Wrap Around

| | | | |
|---|---|---|---|
| PCB138 Lipid Adj | 100.00 | 17 | no |
| PCB138 & 158 Lipid Adj | 100.00 | 17 | no |
| PCB153 Lipid Adj | 100.00 | 17 | no |
| PCB28 | 100.00 | 17 | no |
| PCB52 | 100.00 | 17 | no |
| PCB74 | 100.00 | 17 | no |
| PCB118 | 100.00 | 17 | no |
| PCB138 | 100.00 | 17 | no |
| PCB138 & 158 | 100.00 | 17 | no |
| PCB153 | 100.00 | 17 | no |
| Strontium, urine | 99.97 | 43 | no |
| Harman | 99.95 | 20 | no |
| Hexachlorobenzene | 99.94 | 17 | no |
| Hexachlorobenzene Lipid Adj | 99.94 | 17 | no |
| PCB99 Lipid Adj | 99.94 | 16.5 | no |
| PCB99 | 99.94 | 16.5 | no |
| N-acetyl-s-(3-hydroxypropyl-1-methyl)-L-cysteine | 99.91 | 74 | no |
| Norharman | 99.90 | 20 | no |
| p,p'-DDE | 99.86 | 51 | no |
| p,p'-DDE Lipid Adj | 99.86 | 51 | no |
| PCB44 Lipid Adj | 99.81 | 17 | no |
| PCB44 | 99.81 | 17 | no |
| N-acetyl-S-(3,4-dihydroxybutyl)-L-cysteine | 99.79 | 75 | no |
| Equol | 99.57 | 86 | no |
| 2 & 3-Hydroxyphenanthrene | 99.55 | 20 | no |
| PCB180 Lipid Adj | 99.50 | 17 | no |
| PCB180 | 99.50 | 17 | no |
| 3-methylhippuric acid & 4-methylhippuric acid | 99.49 | 75 | no |
| Enterodiol | 99.49 | 71 | no |
| N-acetyl-S-(2-carbamoylethyl)-L-cysteine | 99.44 | 74 | no |
| PCB49 Lipid Adj | 99.43 | 17 | no |
| PCB49 | 99.43 | 17 | no |
| N-acetyl-S-(3-Hydroxypropyl)-L-cysteine | 99.39 | 75 | no |
| N-acetyl-S-(N-methylcarbamoyl)-L-cysteine | 99.30 | 75 | no |
| Isopentanaldehyde | 99.14 | 18 | no |
| Linear perfluorooctanoate | 99.13 | 20 | no |
| N-Acetyl-S-(benzyl)-L-cysteine | 99.09 | 75 | no |
| Linear perfluorooctane sulfonate | 99.03 | 20 | no |
| PCB66 Lipid Adj | 99.01 | 17 | no |
| PCB66 | 99.01 | 17 | no |

**Formatted:** Position: Horizontal: Right, Relative to: Margin, Vertical: 0", Relative to: Paragraph, Wrap Around

| | | | |
|---|---|---|---|
| PCB146 Lipid Adj | 98.88 | 17 | no |
| PCB146 | 98.88 | 17 | no |
| PCB187 Lipid Adj | 98.63 | 17 | no |
| PCB187 | 98.63 | 17 | no |
| PCB170 Lipid Adj | 98.57 | 17 | no |
| PCB170 | 98.57 | 17 | no |
| Mandelic acid | 98.56 | 74 | no |
| N-acetyl-S-(2-Carboxyethyl)-L-cysteine | 98.54 | 75 | no |
| Branched iso of PFOS | 98.27 | 20 | no |
| PCB110 Lipid Adj | 98.18 | 17 | no |
| PCB110 | 98.18 | 17 | no |
| 1,2,3,4,6,7,8-Heptachlororodibenzo-p-dioxin Lipid Adj | 98.11 | 35 | no |
| 1,2,3,4,6,7,8-Heptachlororodibenzo-p-dioxin | 98.11 | 35 | no |
| Total Hydroxycotinine, urine | 98.07 | 20 | no |
| o-Anisidine, urine | 97.90 | 20 | no |
| PCB105 Lipid Adj | 97.74 | 17 | no |
| PCB105 | 97.74 | 17 | no |
| Total Cotinine, urine | 97.71 | 20 | no |
| 2,2',4,4'-tetrabromodiphenyl ether lipid adj | 97.71 | 17 | no |
| 2,2',4,4'-tetrabromodiphenyl ether | 97.71 | 17 | no |
| N-Acetyl-S-(4-hydroxy-2-butenyl)-L-Cysteine | 97.62 | 73 | no |
| Hexanaldehyde | 97.55 | 16 | no |
| PCB101 Lipid Adj | 96.65 | 17 | no |
| PCB101 | 96.65 | 17 | no |
| Ethylene Oxide | 95.92 | 20 | no |
| PCB149 Lipid Adj | 95.85 | 17 | no |
| PCB149 | 95.85 | 17 | no |
| PCB206 Lipid Adj | 95.72 | 16.5 | no |
| PCB206 | 95.72 | 16.5 | no |
| PCB209 Lipid Adj | 95.18 | 16 | no |
| PCB209 | 95.18 | 16 | no |
| Benzaldehyde | 94.78 | 16 | no |
| N-acetyl-S-(2-hydroxypropyl)-L-cysteine | 94.26 | 75 | no |
| Estradiol | 93.89 | 58 | no |
| 2-Methylhippuric acid | 93.74 | 74 | no |
| 2,2',4,4',6-pentabromodiphenyl lipid adj | 93.48 | 17 | no |
| 2,2',4,4',6-pentabromodiphenyl ether | 93.48 | 17 | no |
| 2,2',4,4',5,5'-hexabromodiphenyl ether lipid adj | 93.25 | 17 | no |
| 2,2',4,4',5,5'-hexabromodiphenyl ether | 93.25 | 17 | no |
| Mono-2-hydroxy-isobutyl phthalate | 92.87 | 22 | no |

**Formatted:** Position: Horizontal: Right, Relative to: Margin, Vertical: 0", Relative to: Paragraph, Wrap Around

| | | | |
|---|---|---|---|
| Phenylglyoxylic acid | 92.58 | 75 | no |
| Bis(1,3-dichloro-2-propyl) phosphate | 92.32 | 45 | no |
| PCB183 Lipid Adj | 92.15 | 17 | no |
| PCB183 | 92.15 | 17 | no |
| Heptaldehyde | 92.06 | 16 | no |
| Diphenyl phosphate | 91.14 | 46 | no |
| PCB196 Lipid Adj | 90.87 | 17 | no |
| PCB196 & 203 Lipid Adj | 90.87 | 17 | no |
| PCB196 & 203 | 90.87 | 17 | no |
| PCB196 | 90.87 | 17 | no |
| PCB199 Lipid Adj | 90.54 | 17 | no |
| PCB199 | 90.54 | 17 | no |
| N-acetyl-S-(2-cyanoethyl)-L-cysteine | 90.53 | 75 | no |
| Urinary Bisphenol S | 90.42 | 22 | no |
| Propanaldehyde | 88.48 | 18 | no |
| PCB156 Lipid Adj | 88.32 | 17 | no |
| PCB156 | 88.32 | 17 | no |
| 4-phenanthrene | 87.90 | 71 | no |
| Methylmercury | 87.77 | 86 | no |
| Tin, urine | 87.12 | 43 | no |
| Bis(2-chloroethyl) phosphate | 86.15 | 46 | no |
| PCB177 Lipid Adj | 86.09 | 17 | no |
| PCB177 | 86.09 | 17 | no |
| o-Toluidine, urine | 85.14 | 19 | no |
| Trans-nonachlor | 84.79 | 52 | no |
| Trans-nonachlor Lipid Adj | 84.79 | 52 | no |
| PCB194 Lipid Adj | 84.25 | 16 | no |
| PCB194 | 84.25 | 16 | no |
| 2-Aminothiazole-4-carboxylic acid | 83.71 | 74 | no |
| PCB87 Lipid Adj | 83.57 | 17 | no |
| PCB87 | 83.57 | 17 | no |
| PCB178 Lipid Adj | 82.20 | 17 | no |
| PCB178 | 82.20 | 17 | no |
| 2,2',4,4',5,5'-hexbrombiphenyl lipid adj | 80.46 | 17 | no |
| 2,2',4,4',5,5'-hexabromobiphenyl | 80.46 | 17 | no |
| 2,4,4'-tribromodiphenyl ether lipid adj | 79.92 | 17 | no |
| 2,4,4'-tribromodiphenyl ether | 79.92 | 17 | no |
| 1,2,3,4,6,7,8,9-Octachlorodibenzo-p-dioxin Lipid Adj | 79.81 | 49 | no |
| 1,2,3,4,6,7,8,9-Octachlorodibenzo-p-dioxin | 79.81 | 49 | no |
| PCB151 Lipid Adj | 79.48 | 17 | no |

**Formatted:** Position: Horizontal: Right, Relative to: Margin, Vertical: 0", Relative to: Paragraph, Wrap Around

| | | | |
|---|---|---|---|
| PCB151 | 79.48 | 17 | no |
| Butyraldehyde | 79.35 | 17 | no |
| 3,3',4,4',5-Pentachlorobiphenyl (pncb) Lipid Adj | 78.94 | 51 | no |
| 3,3',4,4',5-Pentachlorobiphenyl (pncb) | 78.94 | 51 | no |
| 4-Aminobiphenyl, urine | 77.33 | 20 | no |
| 1,2,3,4,6,7,8-Heptachlorodibenzofuran (hpcdf) Lipid Adj | 75.70 | 49 | no |
| 1,2,3,4,6,7,8-Heptachlorodibenzofuran | 75.70 | 49 | no |
| 3,3',4,4',5,5'-hexachlorobiphenyl Lipid Adj | 75.53 | 33.5 | no |
| 3,3',4,4',5,5'-hexachlorobiphenyl | 75.53 | 33.5 | no |
| 2-Aminonaphthalene, urine | 74.99 | 20 | no |
| Oxychlordane | 73.18 | 49 | no |
| Oxychlordane Lipid Adj | 73.18 | 49 | no |
| 1,2,3,6,7,8-Hexachlorodibenzo-p-dioxin Lipid Adj | 73.13 | 52 | no |
| 1,2,3,6,7,8-Hexachlorodibenzo-p-dioxin | 73.13 | 52 | no |
| N-Acetyl-S-(n-propyl)-L-cysteine | 72.46 | 75 | no |
| PCB172 Lipid Adj | 72.23 | 17 | no |
| PCB172 | 72.23 | 17 | no |
| 1-Aminonaphthalene, urine | 71.59 | 19 | no |
| 3-fluoranthene | 71.37 | 20 | no |
| Mono-3-hydroxy-n-butyl phthalate | 70.35 | 22 | no |
| PCB157 Lipid Adj | 70.20 | 17 | no |
| PCB157 | 70.20 | 17 | no |
| Dieldrin | 70.07 | 38 | no |
| Dieldrin Lipid Adj | 70.07 | 38 | no |
| 2,2',4,4',5-pentabromodiphenyl ether lipid adj | 68.42 | 17 | no |
| 2,2',4,4',5-pentabromodiphenyl ether | 68.42 | 17 | no |
| Beta-hexachlorocyclohexane | 68.23 | 52 | no |
| Beta-hexachlorocyclohexane Lipid Adj | 68.23 | 52 | no |
| Urinary Bisphenol F | 65.75 | 22 | no |
| 2,6-Dimethylaniline, urine | 65.19 | 18 | no |
| PCB167 Lipid Adj | 63.62 | 16.5 | no |
| PCB167 | 63.62 | 16.5 | no |
| 1,2,3,4,7,8-Hexachlorodibenzofuran Lipid Adj | 61.90 | 52 | no |
| 1,2,3,4,7,8-Hexachlorodibenzofuran | 61.90 | 52 | no |
| PCB195 Lipid Adj | 60.85 | 16 | no |
| PCB195 | 60.85 | 16 | no |
| 2,3,4,7,8-Pentachlorodibenzofuran Lipid Adj | 60.42 | 52 | no |
| 2,3,4,7,8-Pentachlorodibenzofuran | 60.42 | 52 | no |
| Dibutyl phosphate | 58.98 | 46 | no |
| 2-Amino-1-methyl-6-phenylimidazo[4,5-b]pyridine | 58.50 | 20 | no |

**Formatted:** Position: Horizontal: Right, Relative to: Margin, Vertical: 0", Relative to: Paragraph, Wrap Around

| | | | |
|---|---|---|---|
| Heptachlor Epoxide | 58.46 | 17 | no |
| Heptachlor Epoxide Lipid Adj | 58.46 | 17 | no |
| Bis(1-chloro-2-propyl) phosphate | 56.68 | 46 | no |
| 2,2',4,4',5,6'-hexabromodiphenyl ether lipid adj | 54.72 | 17 | no |
| 2,2',4,4',5,6'-hexabromodiphenyl ether | 54.72 | 17 | no |
| 2-Amino-9H-pyrido[2,3-b]indole | 53.31 | 20 | no |
| Hydroxycotinine, Serum | 49.79 | 60 | no |
| N-acetyl-S-(2-carbamoyl-2-hydroxyethyl)-L-cysteine | 49.46 | 75 | no |
| p,p'-DDT | 48.50 | 49 | no |
| p,p'-DDT Lipid Adj | 48.50 | 49 | no |
| 1,2,3,6,7,8-Hexachlorodibenzofuran Lipid Adj | 47.64 | 52 | no |
| 1,2,3,6,7,8-Hexachlorodibenzofuran | 47.64 | 52 | no |
| Urinary Triclocarban | 41.64 | 22 | no |
| N-acetyl-S-(2-Hydroxyethyl)-L-cysteine | 41.00 | 75 | no |
| cis-3-(2,2-dichlorovinyl)-2,2-dimethylcyclopropane carboxylic acid | 38.66 | 37 | no |
| 2-thioxothiazolidine-4-carboxylic acid | 37.77 | 75 | no |
| Nonanaldehyde | 36.86 | 16 | no |
| 1,2,3,7,8-Pentachlorodibenzo-p-dioxin Lipid Adj | 35.94 | 52 | no |
| 1,2,3,7,8-Pentachlorodibenzo-p-dioxin | 35.94 | 52 | no |
| Alachlor mercapturate | 34.64 | 13 | no |
| Pentanaldehyde | 34.02 | 18 | no |
| Crotonaldehyde | 33.48 | 17 | no |
| Pentachlorophenol | 32.64 | 19 | no |
| N-Acetyl-S-(phenyl)-L-cysteine | 32.08 | 75 | no |
| 1,2,3,4,7,8-Hexachlorodibenzo-p-dioxin Lipid Adj | 31.17 | 35 | no |
| 1,2,3,4,7,8-Hexachlorodibenzo-p-dioxin | 31.17 | 35 | no |
| 2,3,7,8-Tetrachlorodienzo-p-dioxin Lipid Adj | 29.34 | 35 | no |
| 2,3,7,8-Tetrachlorodienzo-p-dioxin | 29.34 | 35 | no |
| 1,2,3,7,8,9-Hexachlorodibenzo-p-dioxin Lipid Adj | 28.41 | 52 | no |
| 1,2,3,7,8,9-Hexachlorodibenzo-p-dioxin | 28.41 | 52 | no |
| Cotinine-n-oxide, urine | 28.08 | 7 | no |
| Mirex | 27.19 | 51 | no |
| Mirex Lipid Adj | 27.19 | 51 | no |
| Nicotine-1 N-oxide, urine | 26.96 | 7 | no |
| PCB189 Lipid Adj | 26.36 | 15 | no |
| PCB189 | 26.36 | 15 | no |
| Nicotine, urine | 25.64 | 7 | no |
| PCB128 Lipid Adj | 25.36 | 17 | no |
| PCB128 | 25.36 | 17 | no |

**Formatted:** Position: Horizontal: Right, Relative to: Margin, Vertical: 0", Relative to: Paragraph, Wrap Around

| | | | |
|---|---|---|---|
| Malathion dicarboxylic acid | 24.91 | 70 | no |
| 2,2',3,4,4'-pentabromodiphenyl ether lipid adj | 24.91 | 17 | no |
| 2,2',3,4,4'-pentabromodiphenyl ether | 24.91 | 17 | no |
| Nornicotine, urine | 24.12 | 7 | no |
| Urinary Perfluorohexanoic acid | 24.08 | 22 | no |
| N-acetyl-S-(phenyl-2-hydroxyethyl)-L-cysteine | 24.04 | 75 | no |
| Anatabine, urine | 23.70 | 7 | no |
| N'-Nitrosanatabine, urine | 23.40 | 66 | no |
| Anabasine, urine | 23.40 | 7 | no |
| N'-Nitrosoanabasine (NAB), urine | 23.00 | 65 | no |
| 2,3',4,4'-tetrabromodiphenyl lipid adj | 22.82 | 17 | no |
| 2,3',4,4'-tetrabromodiphenyl ether | 22.82 | 17 | no |
| MHNCH | 19.85 | 42 | no |
| N'-Nitrosonornicotine, urine | 19.50 | 66 | no |
| 2-Amino-3-methyl-9H-pyrido[2,3-b]indole | 19.31 | 20 | no |
| Branched isomers of perfluorooctanoate | 18.25 | 20 | no |
| 2,2',3,4,4',5',6-heptabromodiphenyl ether lipid adj | 16.77 | 17 | no |
| 2,2',3,4,4',5',6-heptabromodiphenyl ether | 16.77 | 17 | no |
| Octanaldehyde | 14.79 | 15 | no |
| 3,4,4',5-Tetrachlorobiphenyl Lipid Adj | 14.16 | 51 | no |
| 3,4,4',5-Tetrachlorobiphenyl | 14.16 | 51 | no |
| N-acetyl-S-(2-hydroxy-3-butenyl)-L-cysteine | 13.63 | 75 | no |
| Di-p-cresyl phosphate | 11.29 | 22 | no |
| Urinary Perfluorobutanoic acid | 10.91 | 22 | no |
| 1,2,3,4,6,7,8,9-Octachlorodibenzofuran Lipid Adj | 10.46 | 49 | no |
| 1,2,3,4,6,7,8,9-Octachlorodibenzofuran | 10.46 | 49 | no |
| Perfluoroheptanoic acid | 9.92 | 154 | no |
| Platinum, urine | 9.46 | 71 | no |
| Urinary 4-tert-octylphenol | 9.03 | 69 | no |
| Ethylenethio urea | 8.85 | 62 | no |
| 4-fluoro-3-phenoxybenzoic acid | 7.08 | 145 | no |
| o,p'-DDT | 6.96 | 17 | no |
| o,p'-DDT Lipid Adj | 6.96 | 17 | no |
| Mono-cyclohexyl phthalate | 6.41 | 145 | no |
| DEET | 6.34 | 145 | no |
| 2,3,4,6,7,8-Hexchlorodibenzofuran Lipid Adj | 6.03 | 52 | no |
| 2,3,4,6,7,8-Hexchlorodibenzofuran | 6.03 | 52 | no |
| 2-(diethylamino)-6-methylpyrimidin-4-ol/one | 5.92 | 22 | no |
| Urinary Arsenocholine | 5.29 | 147 | no |
| N-Nitrosomorpholine (NMOR) | 5.05 | 20 | no |

**Formatted:** Position: Horizontal: Right, Relative to: Margin, Vertical: 0", Relative to: Paragraph, Wrap Around

| | | | |
|---|---|---|---|
| 2,2',4-tribromodiphenyl ether lipid adj | 4.94 | 17 | no |
| 2,2',4-tribromodiphenyl ether | 4.94 | 17 | no |
| N-Nitrosopiperidine (NPIP) | 4.83 | 20 | no |
| 2,3,4,5-tetrabromobenzoic acid | 4.50 | 46 | no |
| o-Tolualdehyde | 4.03 | 16 | no |
| 3-chloro-7-hydroxy-4-methyl-2H-chromen-2-one/ol | 3.67 | 22 | no |
| Perfluorododecanoic acid | 3.64 | 133 | no |
| Metolachlor mercapturate | 3.48 | 23 | no |
| Mono-n-octyl phthalate | 3.39 | 145 | no |
| 1,2,3,4,7,8,9-Heptachlorodibenzofuran Lipid Adj | 3.22 | 33.5 | no |
| 1,2,3,4,7,8,9-Heptachlorodibenzofuran | 3.22 | 33.5 | no |
| Carbofuranphenol | 3.20 | 57 | no |
| Urinary Arsenic acid | 3.18 | 147 | no |
| Ethylmercury | 2.82 | 86 | no |
| Acetochlor mercapturate | 2.37 | 22 | no |
| 1-Methyl-3-amino-5H-pyrido[4,3-b]indole | 2.29 | 20 | no |
| 2-amino-3-methyl-3H-imidazo[4,5-f] quinolone | 2.19 | 20 | no |
| 2,4,5-Trichlorophenoxyacetic acid | 2.16 | 98 | no |
| 2-Amino-6-methyldipyrido[1,2-a:3',2'-d]imidazole | 2.09 | 20 | no |
| Blood Heptane | 1.89 | 30 | no |
| Acephate | 1.84 | 61 | no |
| Perfluorobutane sulfonic acid | 1.82 | 137 | no |
| Blood Cyclohexane | 1.77 | 29 | no |
| Blood Methylcyclopentane | 1.76 | 30 | no |
| 3-Amino-1,4-dimethyl-5H-pyrido[4,3-b]indole | 1.73 | 20 | no |
| Blood Ethyl Acetate | 1.73 | 30 | no |
| Gamma-hexachlorocyclohexane | 1.64 | 51 | no |
| Gamma-hexachlorocyclohexane Lipid Adj | 1.64 | 51 | no |
| 2,3,7,8-Tetrachlorodibenzofuran Lipid Adj | 1.60 | 51 | no |
| 2,3,7,8-Tetrachlorodibenzofuran | 1.60 | 51 | no |
| Blood Trichloroethene | 1.43 | 211 | no |
| Blood Hexane | 1.42 | 85 | no |
| Blood Octane | 1.31 | 30 | no |
| Blood 1,2-Dichloroethane | 1.26 | 186 | no |
| Atrazine mercapturate | 1.21 | 59 | no |
| Blood Carbon Tetrachloride | 1.19 | 209 | no |
| Blood 1,1,1-Trichloroethane | 1.11 | 185 | no |
| Urinary Perfluoroheptanoic acid | 1.08 | 22 | no |
| 1,2,3,7,8-Pentachlorodibenzofuran Lipid Adj | 1.02 | 52 | no |
| 1,2,3,7,8-Pentachlorodibenzofuran | 1.02 | 52 | no |

**Formatted:** Position: Horizontal: Right, Relative to: Margin, Vertical: 0", Relative to: Paragraph, Wrap Around

| | | | |
|---|---|---|---|
| Beryllium, urine | 1.00 | 142 | no |
| Urinary GenX (HFPO-DA propanoateammonium salt of heptafluoropropoxy)-propanoate | 0.93 | 22 | no |
| cis-3-(2,2-dibromovinyl)-2,2-dimethylcyclopropane carboxylic acid | 0.90 | 96 | no |
| N-Nitrosoethylmethylamine (NMEA) | 0.89 | 20 | no |
| N-acetyl-s-(1-hydroxyMethyl)-2-propenyl)-L-cysteine | 0.87 | 75 | no |
| Blood Tetrahydrofuran | 0.86 | 30 | no |
| Decanaldehyde | 0.85 | 14 | no |
| Urinary Trimethylarsine Oxide | 0.76 | 128 | no |
| 2-isopropoxyphenol | 0.56 | 56 | no |
| Blood isopropylbenzene | 0.46 | 118 | no |
| N-Nitrosopyrrolidine | 0.46 | 20 | no |
| N-acetyl-S-(2,2-Dichlorovinyl)-L-cysteine | 0.45 | 75 | no |
| N-Nitrosodiethylamine (NDEA) | 0.42 | 20 | no |
| Blood Chloroethane | 0.42 | 30 | no |
| Diaminochloroatrazine | 0.40 | 24 | no |
| Methamidophos | 0.37 | 62 | no |
| N-acetyl-S-(dimethylphenyl)-L-cysteine | 0.33 | 73 | no |
| 2-Aminodipyrido[1,2-a:3',2'-d]imidazole | 0.31 | 20 | no |
| Blood 1,3-Dichlorobenzene | 0.30 | 182 | no |
| Blood Methylene Chloride | 0.30 | 168 | no |
| Dimethoate | 0.26 | 62 | no |
| Blood Diethyl Ether | 0.22 | 29 | no |
| 1,2,3,7,8,9-Hexachlorodibenzofuran Lipid Adj | 0.18 | 51 | no |
| 1,2,3,7,8,9-Hexachlorodibenzofuran | 0.18 | 51 | no |
| Propylenethio urea | 0.17 | 62 | no |
| Aldrin | 0.17 | 40 | no |
| Aldrin Lipid Adj | 0.17 | 40 | no |
| Desethyl atrazine | 0.16 | 23 | no |
| Blood Dibromomethane | 0.15 | 155 | no |
| Endrin | 0.15 | 36 | no |
| Endrin Lipid Adj | 0.15 | 36 | no |
| Ethametsulfuron methyl | 0.14 | 61 | no |
| Blood Chlorobenzene | 0.13 | 185 | no |
| Nicosulfuron | 0.13 | 56 | no |
| Desisopropyl atrazine mercapturate | 0.10 | 22 | no |
| Di-o-cresyl phosphate | 0.10 | 22 | no |
| Urinary Branched isomers of perfluorooctanoate | 0.10 | 22 | no |
| Blood trans-1,2-Dichloroethene | 0.08 | 156 | no |
| Dibenzyl phosphate | 0.08 | 46 | no |

**Formatted:** Position: Horizontal: Right, Relative to: Margin, Vertical: 0", Relative to: Paragraph, Wrap Around

| | | | |
|---|---|---|---|
| N-acetyl-S-(1,2-dichlorovinyl)-L-cysteine | 0.06 | 75 | no |
| N-acetyl-s-(trichlorovinyl)-L-cysteine | 0.06 | 75 | no |
| Desisopropyl atrazine | 0.05 | 22 | no |
| Urinary Perfluoroheptane sulfonic acid | 0.05 | 22 | no |
| Urinary Perfluorohexane sulfonic acid | 0.05 | 22 | no |
| Urinary Perfluorononanoic acid | 0.05 | 22 | no |
| Blood 1,2-Dichlorobenzene | 0.05 | 116 | no |
| Blood 1,1,2,2-Tetrachloroethane | 0.04 | 156 | no |
| Metsulfuron methyl | 0.04 | 61 | no |
| Sulfosulfuron | 0.04 | 59 | no |
| O-methoate | 0.03 | 62 | no |
| Blood 1,2-Dichloropropane | 0.02 | 157 | no |
| Chlorsulfuron | 0.02 | 59 | no |
| Prosulfuron | 0.02 | 60 | no |
| Oxasulfuron | 0.02 | 62 | no |
| Blood 1,2,3-trichloropropane | 0.01 | 117 | no |
| Blood Hexachloroethane | 0.01 | 141 | no |
| Blood Nitrobenzene | 0.01 | 175 | no |
| Blood 1,1-Dichloroethane | 0 | 156 | no |
| Blood cis-1,2-Dichloroethene | 0 | 156 | no |
| Blood 1,1,2-Trichloroethane | 0 | 155 | no |
| Blood 1,1-Dichloroethene | 0 | 153 | no |
| Blood 1,1,1,2-tetrachloroethane | 0 | 119 | no |
| Blood 1,2-dibromoethane | 0 | 117 | no |
| Blood 1,2-Dibromo-3-chloropropane | 0 | 115 | no |
| Blood 1,4-Dioxane | 0 | 75 | no |
| Rimsulfuron | 0 | 62 | no |
| Sulfometuron methyl | 0 | 61 | no |
| Thifensulfuron methyl | 0 | 61 | no |
| Bensulfuron methyl | 0 | 60 | no |
| Mesosulfuron methyl | 0 | 60 | no |
| Triasulfuron | 0 | 60 | no |
| Triflusulfuron methyl | 0 | 59 | no |
| Halosulfuron | 0 | 58 | no |
| Primisulfuron methyl | 0 | 58 | no |
| Foramsulfuron | 0 | 56 | no |
| Blood aaa-Trifluorotoluene | 0 | 30 | no |
| Blood Vinyl Bromide | 0 | 30 | no |
| Atrazine | 0 | 24 | no |

**Formatted:** Position: Horizontal: Right, Relative to: Margin, Vertical: 0", Relative to: Paragraph, Wrap Around

| | | | |
|---|---|---|---|
| Urinary 9-chlorohexadecafluoro-3-oxanonane-1-sulfonate | 0 | 22 | no |
| Urinary Adona (ammonium salt of 4,8-dioxa-3H-perfluorononanoate) | 0 | 22 | no |
| Urinary Perfluoropentanoic acid | 0 | 22 | no |
| Urinary Branched iso of PFOS | 0 | 22 | no |
| Urinary Perfluorobutane sulfonic acid | 0 | 22 | no |
| Urinary Perfluorodecanoic acid | 0 | 22 | no |
| Urinary Perfluoroundecanoic acid | 0 | 22 | no |

**Formatted:** Position: Horizontal: Right, Relative to: Margin, Vertical: 0", Relative to: Paragraph, Wrap Around

**Table S5. Chemical name and class for included 129 substances.**

| Chemical Name | Chemical Class |
|---|---|
| Acrylamide | Acrylamide |
| Glycideamide | Acrylamide |
| Methylmalonic Acid | Dietary Components |
| 25-hydroxyvitamin D2 | Dietary Components |
| 25-hydroxyvitamin D3 | Dietary Components |
| epi-25-hydroxyvitamin D3 | Dietary Components |
| Vitamin C | Dietary Components |
| Vitamin D | Dietary Components |
| 4-pyridoxic acid | Dietary Components |
| a-Carotene | Dietary Components |
| Vitamin B12, serum | Dietary Components |
| trans-b-carotene | Dietary Components |
| cis-b-carotene | Dietary Components |
| b-cryptoxanthin | Dietary Components |
| g-tocopherol | Dietary Components |
| total Lycopene | Dietary Components |
| Combined Lutein/zeaxanthin | Dietary Components |
| trans-lycopene | Dietary Components |
| Vitamin B6 | Dietary Components |
| Retinyl palmitate | Dietary Components |
| Retinyl stearate | Dietary Components |
| Total Calcium | Dietary Components |
| Chloride | Dietary Components |
| Potassium | Dietary Components |
| Sodium | Dietary Components |
| Vitamin A | Dietary Components |
| d-Tocopherol | Dietary Components |
| Blood cadmium | Metals |
| Blood lead | Metals |
| Mercury, Inorganic | Metals |
| Blood mercury, total | Metals |
| Urinary Arsenous acid | Metals |
| Urinary Arsenobetaine | Metals |
| Urinary total Arsenic | Metals |
| Barium, urine | Metals |
| Cadmium, urine | Metals |
| Cobalt, urine | Metals |

**Formatted:** Position: Horizontal: Right, Relative to: Margin, Vertical: 0", Relative to: Paragraph, Wrap Around

| | |
|---|---|
| Cesium, urine | Metals |
| Urinary Dimethylarsonic acid | Metals |
| Mercury, urine | Metals |
| Urinary Monomethylarsonic acid | Metals |
| Molybdenum, urine | Metals |
| Lead, urine | Metals |
| Antimony, urine | Metals |
| Thallium, urine | Metals |
| Tungsten, urine | Metals |
| Uranium, urine | Metals |
| Sum of arsenic metabolites | Metals |
| Urinary nitrate | Other |
| Urinary thiocyanate | Other |
| Iodine, urine | Other |
| Perchlorate, urine | Other |
| 2-(N-ethyl-PFOSA) acetate | Per- and Polyfluoroalkyl Substances (PFAS) |
| 2-(N-methyl-PFOSA) acetate | Per- and Polyfluoroalkyl Substances (PFAS) |
| Perfluorodecanoic acid | Per- and Polyfluoroalkyl Substances (PFAS) |
| Perfluorohexane sulfonic acid | Per- and Polyfluoroalkyl Substances (PFAS) |
| Perfluorononanoic acid | Per- and Polyfluoroalkyl Substances (PFAS) |
| Perfluorooctanoic acid | Per- and Polyfluoroalkyl Substances (PFAS) |
| Perfluorooctane sulfonic acid | Per- and Polyfluoroalkyl Substances (PFAS) |
| Perfluorooctane sulfonamide | Per- and Polyfluoroalkyl Substances (PFAS) |
| Perfluoroundecanoic acid | Per- and Polyfluoroalkyl Substances (PFAS) |
| Urinary Benzophenone-3 | Personal Care & Consumer Product Compounds |
| Urinary Bisphenol A | Personal Care & Consumer Product Compounds |
| Butyl paraben | Personal Care & Consumer Product Compounds |
| Ethyl paraben | Personal Care & Consumer Product Compounds |
| Methyl paraben | Personal Care & Consumer Product Compounds |

**Formatted:** Position: Horizontal: Right, Relative to: Margin, Vertical: 0", Relative to: Paragraph, Wrap Around

| | |
|---|---|
| Propyl paraben | Personal Care & Consumer Product Compounds |
| Urinary Triclosan | Personal Care & Consumer Product Compounds |
| 2,5-dichlorophenol | Pesticides |
| 2,4,5-trichlorophenol | Pesticides |
| 2,4-D | Pesticides |
| 2,4,6-trichlorophenol | Pesticides |
| 3,5,6-trichloropyridinol | Pesticides |
| 2,4-dichlorophenol | Pesticides |
| DEET acid | Pesticides |
| Desethyl hydroxy DEET | Pesticides |
| Dimethyl phosphate | Pesticides |
| Diethylphosphate | Pesticides |
| Dimethyl thiophosphate | Pesticides |
| Diethylthiophosphate | Pesticides |
| Dimethyl dithiophosphate | Pesticides |
| Diethyl dithiophosphate | Pesticides |
| 3-phenoxybenzoic acid | Pesticides |
| O-Phenylphenol | Pesticides |
| Oxypyrimidine | Pesticides |
| Paranitrophenol | Pesticides |
| trans-3-(2,2-dichlorovinyl)-2,2-dimethylcyclopropane carboxylic acid | Pesticides |
| Mono(carboxynonyl) phthalate | Phthalates & Plasticizers |
| Mono(carboxyoctyl) phthalate | Phthalates & Plasticizers |
| Mono-2-ethyl-5-carboxypentyl phthalate | Phthalates & Plasticizers |
| Mono-n-butyl phthalate | Phthalates & Plasticizers |
| Mono-(3-carboxypropyl) phthalate | Phthalates & Plasticizers |
| Mono-ethyl phthalate | Phthalates & Plasticizers |
| Mono-(2-ethyl-5-hydroxyhexyl) phthalate | Phthalates & Plasticizers |
| Mono-(2-ethylhexyl) phthalate | Phthalates & Plasticizers |
| Mono-isobutyl phthalate | Phthalates & Plasticizers |
| Mono-n-methyl phthalate | Phthalates & Plasticizers |
| Mono-isononyl phthalate | Phthalates & Plasticizers |
| Mono-(2-ethyl-5-oxohexyl) phthalate | Phthalates & Plasticizers |
| Mono-benzyl phthalate | Phthalates & Plasticizers |
| Sum of metabolites of DEHP | Phthalates & Plasticizers |
| Daidzein | Phytoestrogens |
| o-Desmethylangolensin (O-DMA) | Phytoestrogens |
| Enterolactone | Phytoestrogens |

**Formatted:** Position: Horizontal: Right, Relative to: Margin, Vertical: 0", Relative to: Paragraph, Wrap Around

| | |
|---|---|
| Genistein | Phytoestrogens |
| 1-naphthol | Polyaromatic Hydrocarbons (PAH) |
| 2-naphthol | Polyaromatic Hydrocarbons (PAH) |
| 3-fluorene | Polyaromatic Hydrocarbons (PAH) |
| 2-fluorene | Polyaromatic Hydrocarbons (PAH) |
| 3-phenanthrene | Polyaromatic Hydrocarbons (PAH) |
| 1-phenanthrene | Polyaromatic Hydrocarbons (PAH) |
| 2-phenanthrene | Polyaromatic Hydrocarbons (PAH) |
| 1-pyrene | Polyaromatic Hydrocarbons (PAH) |
| 9-fluorene | Polyaromatic Hydrocarbons (PAH) |
| Cotinine | Smoking Related Compounds |
| NNAL , urine | Smoking Related Compounds |
| Blood 2,5-Dimethylfuran | Volatile Organic Compounds (VOC) |
| Blood Tetrachloroethene | Volatile Organic Compounds (VOC) |
| Blood Bromoform | Volatile Organic Compounds (VOC) |
| Blood Bromodichloromethane | Volatile Organic Compounds (VOC) |
| Blood Benzene | Volatile Organic Compounds (VOC) |
| Blood Chloroform | Volatile Organic Compounds (VOC) |
| Blood Dibromochloromethane | Volatile Organic Compounds (VOC) |
| Blood 1,4-Dichlorobenzene | Volatile Organic Compounds (VOC) |
| Blood Ethylbenzene | Volatile Organic Compounds (VOC) |
| Blood furan | Volatile Organic Compounds (VOC) |
| Blood MTBE | Volatile Organic Compounds (VOC) |
| Blood o-Xylene | Volatile Organic Compounds (VOC) |
| Blood Styrene | Volatile Organic Compounds (VOC) |
| Blood Toluene | Volatile Organic Compounds (VOC) |
| Blood m-/p-Xylene | Volatile Organic Compounds (VOC) |

Table S6. Number of participants by included chemicals. Participants have data available for age, sex, race, poverty income ratio, smoking, cycle, occupational group, and given chemical. The total number of participants is 51,008 who have data available for age, sex, race, poverty income ratio, smoking, cycle, and occupational group.

| Chemical Name | Number of Participants |
|---|---|
| Cotinine | 41710 |
| Sodium | 41500 |
| Chloride | 41498 |
| Potassium | 41498 |
| Total Calcium | 41472 |
| Blood cadmium | 38933 |
| Blood lead | 38933 |
| Vitamin D | 36596 |
| Blood mercury, total | 32433 |
| Mercury, Inorganic | 32320 |
| Vitamin B12, serum | 24322 |
| 25-hydroxyvitamin D3 | 20866 |
| 25-hydroxyvitamin D2 | 20865 |
| epi-25-hydroxyvitamin D3 | 20565 |
| NNAL , urine | 20173 |
| Perchlorate, urine | 18893 |
| Urinary nitrate | 17219 |
| Urinary thiocyanate | 17218 |
| Vitamin B6 | 16052 |
| 4-pyridoxic acid | 16048 |
| Iodine, urine | 15620 |
| Blood o-Xylene | 14766 |
| Blood m-/p-Xylene | 14731 |
| Vitamin A | 14683 |
| d-Tocopherol | 14650 |
| Blood Benzene | 14594 |
| Blood Ethylbenzene | 14580 |
| Blood Dibromochloromethane | 14542 |
| Blood Tetrachloroethene | 14490 |
| Blood MTBE | 14181 |
| g-tocopherol | 14098 |
| Blood 2,5-Dimethylfuran | 13795 |
| Blood Bromoform | 13767 |
| Mercury, urine | 13678 |
| Cobalt, urine | 13623 |

**Formatted:** Position: Horizontal: Right, Relative to: Margin, Vertical: 0", Relative to: Paragraph, Wrap Around

| | |
|---|---|
| Lead, urine | 13623 |
| Cesium, urine | 13622 |
| Retinyl palmitate | 13613 |
| Thallium, urine | 13575 |
| Mono-n-butyl phthalate | 13531 |
| Mono-(2-ethylhexyl) phthalate | 13531 |
| Mono-isononyl phthalate | 13531 |
| Mono-benzyl phthalate | 13531 |
| Tungsten, urine | 13530 |
| Mono-ethyl phthalate | 13527 |
| Antimony, urine | 13512 |
| Molybdenum, urine | 13501 |
| Cadmium, urine | 13490 |
| Barium, urine | 13475 |
| Retinyl stearate | 12590 |
| 1-phenanthrene | 12362 |
| 2-fluorene | 12351 |
| 3-fluorene | 12310 |
| Blood Bromodichloromethane | 12305 |
| Uranium, urine | 12253 |
| Mono-(3-carboxypropyl) phthalate | 12051 |
| Mono-(2-ethyl-5-hydroxyhexyl) phthalate | 12051 |
| Mono-isobutyl phthalate | 12051 |
| Mono-(2-ethyl-5-oxohexyl) phthalate | 12051 |
| Blood 1,4-Dichlorobenzene | 12049 |
| Blood Toluene | 11669 |
| Acrylamide | 11588 |
| Blood Chloroform | 11565 |
| Glycideamide | 11559 |
| Perfluorooctanoic acid | 11463 |
| Perfluorooctane sulfonic acid | 11463 |
| Perfluorodecanoic acid | 11460 |
| Perfluorohexane sulfonic acid | 11460 |
| Perfluorononanoic acid | 11460 |
| Perfluoroundecanoic acid | 11460 |
| 2,4-D | 11125 |
| 2-naphthol | 10614 |
| 1-pyrene | 10576 |
| 1-naphthol | 10567 |
| 2-phenanthrene | 10529 |

**Formatted:** Position: Horizontal: Right, Relative to: Margin, Vertical: 0", Relative to: Paragraph, Wrap Around

| | |
|---|---|
| 3-phenanthrene | 10522 |
| Urinary Arsenobetaine | 10517 |
| Urinary Arsenous acid | 10517 |
| Urinary Dimethylarsonic acid | 10517 |
| Urinary Monomethylarsonic acid | 10516 |
| Sum of arsenic metabolites | 10516 |
| Urinary total Arsenic | 10505 |
| Vitamin C | 10348 |
| trans-b-carotene | 10339 |
| Combined Lutein/zeaxanthin | 10339 |
| trans-lycopene | 10339 |
| a-Carotene | 10332 |
| b-cryptoxanthin | 10293 |
| 2,5-dichlorophenol | 10291 |
| 2,4-dichlorophenol | 10291 |
| Urinary Benzophenone-3 | 10287 |
| Urinary Bisphenol A | 10287 |
| Urinary Triclosan | 10287 |
| Mono-2-ethyl-5-carboxypentyl phthalate | 10284 |
| Sum of metabolites of DEHP | 10284 |
| total Lycopene | 10260 |
| Mono-n-methyl phthalate | 10260 |
| o-Desmethylangolensin (O-DMA) | 9927 |
| cis-b-carotene | 9882 |
| Blood furan | 9735 |
| 2-(N-ethyl-PFOSA) acetate | 9671 |
| 2-(N-methyl-PFOSA) acetate | 9671 |
| Perfluorooctane sulfonamide | 9671 |
| Methylmalonic Acid | 9476 |
| 3-phenoxybenzoic acid | 9457 |
| trans-3-(2,2-dichlorovinyl)-2,2-dimethylcyclopropane carboxylic acid | 9433 |
| Dimethyl thiophosphate | 9344 |
| Dimethyl phosphate | 9334 |
| Diethylphosphate | 9328 |
| Dimethyl dithiophosphate | 9327 |
| Diethylthiophosphate | 9294 |
| Diethyl dithiophosphate | 9211 |
| 9-fluorene | 8823 |
| Oxypyrimidine | 8654 |
| Butyl paraben | 8615 |

**Formatted:** Position: Horizontal: Right, Relative to: Margin, Vertical: 0", Relative to: Paragraph, Wrap Around

| | |
|---|---|
| Ethyl paraben | 8615 |
| Methyl paraben | 8615 |
| Propyl paraben | 8615 |
| Mono(carboxynonyl) phthalate | 8614 |
| Mono(carboxyoctyl) phthalate | 8614 |
| Paranitrophenol | 8484 |
| Genistein | 8457 |
| Daidzein | 8318 |
| Enterolactone | 8317 |
| Blood Styrene | 7991 |
| 3,5,6-trichloropyridinol | 7985 |
| 2,4,5-trichlorophenol | 6895 |
| 2,4,6-trichlorophenol | 6895 |
| O-Phenylphenol | 6895 |
| Desethyl hydroxy DEET | 6876 |
| DEET acid | 6868 |

**Formatted:** Position: Horizontal: Right, Relative to: Margin, Vertical: 0", Relative to: Paragraph, Wrap Around

**Table S7. Distribution statistics of chemical biomarkers.**

| Chemical Name | Minimum | 25th Percentile | Median | Mean | 75th Percentile | Maximum |
|---|---|---|---|---|---|---|
| 1-naphthol | 31.60 | 838.95 | 2071.00 | 29555.39 | 7572.55 | 35920000.00 |
| 1-phenanthrene | 2.80 | 73.00 | 137.00 | 225.82 | 257.85 | 22334.00 |
| 1-pyrene | 3.50 | 52.00 | 112.90 | 224.27 | 238.05 | 12517.90 |
| 2-(N-ethyl-PFOSA) acetate | 0.07 | 0.07 | 0.14 | 0.24 | 0.30 | 24.60 |
| 2-(N-methyl-PFOSA) acetate | 0.06 | 0.13 | 0.30 | 0.49 | 0.60 | 54.30 |
| 2-fluorene | 2.80 | 126.00 | 267.00 | 705.02 | 686.50 | 75607.00 |
| 2-naphthol | 9.30 | 1945.65 | 4476.55 | 9000.49 | 10627.08 | 590676.00 |
| 2-phenanthrene | 2.10 | 35.10 | 68.00 | 117.17 | 130.10 | 12420.00 |
| 2,4-D | 0.01 | 0.14 | 0.28 | 0.66 | 0.47 | 1232.00 |
| 2,4-dichlorophenol | 0.07 | 0.40 | 0.80 | 5.66 | 2.10 | 1230.00 |
| 2,4,5-trichlorophenol | 0.07 | 0.07 | 0.07 | 0.14 | 0.10 | 17.50 |
| 2,4,6-trichlorophenol | 0.35 | 0.35 | 0.35 | 0.60 | 0.50 | 95.00 |
| 2,5-dichlorophenol | 0.07 | 1.90 | 7.10 | 190.98 | 31.30 | 47200.00 |
| 25-hydroxyvitamin D2 | 1.45 | 1.45 | 1.45 | 3.84 | 1.45 | 315.00 |
| 25-hydroxyvitamin D3 | 3.02 | 40.08 | 57.10 | 59.40 | 74.90 | 373.67 |
| 3-fluorene | 1.40 | 45.00 | 98.30 | 356.89 | 316.00 | 23854.60 |
| 3-phenanthrene | 2.80 | 46.30 | 93.00 | 212.30 | 190.00 | 126202.00 |
| 3-phenoxybenzoic acid | 0.07 | 0.14 | 0.44 | 1.77 | 1.09 | 999.61 |
| 3,5,6-trichloropyridinol | 0.07 | 0.56 | 1.32 | 2.40 | 2.71 | 180.00 |
| 4-pyridoxic acid | 0.50 | 15.00 | 23.30 | 84.52 | 45.13 | 18800.00 |
| 9-fluorene | 7.10 | 152.85 | 315.00 | 632.44 | 662.50 | 79645.00 |
| a-Carotene | 0.21 | 1.28 | 2.40 | 4.02 | 4.72 | 98.71 |
| Acrylamide | 2.12 | 39.50 | 52.20 | 74.17 | 77.50 | 910.00 |
| Antimony, urine | 0.02 | 0.04 | 0.07 | 0.11 | 0.13 | 47.00 |

Format
Vertical

| | | | | | | |
|---|---|---|---|---|---|---|
| b-cryptoxanthin | 0.14 | 5.05 | 7.88 | 10.12 | 12.52 | 106.00 |
| Barium, urine | 0.04 | 0.65 | 1.32 | 2.24 | 2.53 | 419.00 |
| Blood 1,4-Dichlorobenzene | 0.03 | 0.03 | 0.08 | 1.24 | 0.22 | 238.00 |
| Blood 2,5-Dimethylfuran | 0.01 | 0.01 | 0.01 | 0.03 | 0.01 | 0.78 |
| Blood Benzene | 0.02 | 0.02 | 0.02 | 0.07 | 0.05 | 2.50 |
| Blood Bromodichloromethane | 0.15 | 0.44 | 1.53 | 3.08 | 3.59 | 93.00 |
| Blood Bromoform | 0.39 | 0.71 | 0.71 | 2.93 | 3.74 | 540.00 |
| Blood cadmium | 0.07 | 0.20 | 0.34 | 0.52 | 0.60 | 10.80 |
| Blood Chloroform | 1.48 | 4.30 | 8.95 | 19.84 | 18.00 | 22000.00 |
| Blood Dibromochloromethane | 0.14 | 0.44 | 1.10 | 2.46 | 3.50 | 180.00 |
| Blood Ethylbenzene | 0.01 | 0.02 | 0.02 | 0.06 | 0.05 | 8.37 |
| Blood furan | 0.02 | 0.02 | 0.02 | 0.03 | 0.02 | 0.47 |
| Blood lead | 0.07 | 0.81 | 1.30 | 1.77 | 2.13 | 61.29 |
| Blood m-/p-Xylene | 0.02 | 0.05 | 0.09 | 0.19 | 0.16 | 23.80 |
| Blood mercury, total | 0.07 | 0.42 | 0.81 | 1.48 | 1.62 | 85.70 |
| Blood MTBE | 0.13 | 0.99 | 2.20 | 14.39 | 7.10 | 3800.00 |
| Blood o-Xylene | 0.02 | 0.02 | 0.03 | 0.06 | 0.04 | 7.91 |
| Blood Styrene | 0.01 | 0.02 | 0.02 | 0.09 | 0.05 | 146.00 |
| Blood Tetrachloroethene | 0.01 | 0.03 | 0.03 | 0.07 | 0.03 | 57.50 |
| Blood Toluene | 0.02 | 0.05 | 0.09 | 0.22 | 0.21 | 56.30 |
| Butyl paraben | 0.07 | 0.14 | 0.14 | 3.80 | 0.60 | 1150.00 |
| Cadmium, urine | 0.00 | 0.12 | 0.23 | 0.38 | 0.47 | 36.78 |
| Cesium, urine | 0.07 | 2.85 | 4.70 | 5.46 | 6.93 | 552.12 |
| Chloride | 70.00 | 102.00 | 104.00 | 103.60 | 105.10 | 119.00 |
| cis-b-carotene | 0.21 | 0.50 | 0.80 | 1.18 | 1.35 | 23.28 |
| Cobalt, urine | 0.02 | 0.22 | 0.37 | 0.54 | 0.58 | 127.87 |
| Combined Lutein/zeaxanthin | 0.14 | 10.49 | 14.30 | 16.05 | 19.49 | 113.05 |
| Cotinine | 0.01 | 0.02 | 0.07 | 53.41 | 8.85 | 1820.00 |
| d-Tocopherol | 2.00 | 8.00 | 798.00 | 769.45 | 1177.60 | 7230.30 |

**Formatted:** Position: Horizontal: Right, Relative to: Margin, Vertical: 0", Relative to: Paragraph, Wrap Around

| | | | | | | |
|---|---|---|---|---|---|---|
| Daidzein | 0.20 | 19.30 | 57.60 | 367.13 | 201.21 | 46000.00 |
| DEET acid | 0.34 | 0.70 | 2.06 | 96.85 | 7.16 | 382000.00 |
| Desethyl hydroxy DEET | 0.06 | 0.06 | 0.06 | 2.19 | 0.06 | 11400.00 |
| Diethyl dithiophosphate | 0.04 | 0.07 | 0.07 | 0.23 | 0.28 | 44.78 |
| Diethylphosphate | 0.07 | 0.26 | 0.54 | 3.63 | 3.60 | 1344.61 |
| Diethylthiophosphate | 0.06 | 0.18 | 0.40 | 1.09 | 0.97 | 205.78 |
| Dimethyl dithiophosphate | 0.06 | 0.07 | 0.36 | 1.60 | 0.58 | 297.12 |
| Dimethyl phosphate | 0.07 | 0.35 | 0.72 | 5.27 | 4.57 | 516.70 |
| Dimethyl thiophosphate | 0.07 | 0.39 | 1.68 | 8.25 | 5.60 | 1800.00 |
| Enterolactone | 0.10 | 102.00 | 344.00 | 783.45 | 844.00 | 122000.00 |
| epi-25-hydroxyvitamin D3 | 1.16 | 1.74 | 2.86 | 3.54 | 4.42 | 59.80 |
| Ethyl paraben | 0.71 | 0.71 | 0.71 | 19.05 | 6.00 | 3130.00 |
| g-tocopherol | 7.00 | 142.00 | 205.50 | 227.12 | 286.00 | 3122.00 |
| Genistein | 0.10 | 9.50 | 26.70 | 176.02 | 89.40 | 25700.00 |
| Glycideamide | 2.83 | 34.80 | 49.00 | 61.70 | 72.20 | 599.00 |
| Iodine, urine | 2.90 | 84.00 | 149.40 | 431.29 | 255.03 | 1406700.00 |
| Lead, urine | 0.02 | 0.30 | 0.57 | 0.86 | 1.00 | 71.70 |
| Mercury, Inorganic | 0.19 | 0.25 | 0.25 | 0.32 | 0.30 | 42.00 |
| Mercury, urine | 0.04 | 0.19 | 0.42 | 0.90 | 0.94 | 83.03 |
| Methyl paraben | 0.71 | 16.95 | 68.70 | 258.56 | 247.00 | 17300.00 |
| Methylmalonic Acid | 26.60 | 107.00 | 139.00 | 171.52 | 186.00 | 5540.00 |
| Molybdenum, urine | 0.57 | 23.90 | 45.20 | 58.23 | 76.20 | 1215.90 |
| Mono-(2-ethyl-5-hydroxyhexyl) phthalate | 0.14 | 6.20 | 13.80 | 45.43 | 31.70 | 9326.10 |
| Mono-(2-ethyl-5-oxohexyl) phthalate | 0.14 | 4.00 | 8.90 | 27.53 | 19.92 | 6079.90 |
| Mono-(2-ethylhexyl) phthalate | 0.35 | 0.80 | 2.00 | 7.35 | 5.20 | 1966.10 |
| Mono-(3-carboxypropyl) phthalate | 0.10 | 1.10 | 2.48 | 6.47 | 5.00 | 1588.70 |
| Mono-2-ethyl-5-carboxypentyl phthalate | 0.14 | 9.70 | 20.40 | 59.65 | 44.00 | 15828.00 |
| Mono-benzyl phthalate | 0.06 | 2.95 | 7.42 | 17.16 | 17.14 | 16510.32 |
| Mono-ethyl phthalate | 0.37 | 31.57 | 91.34 | 390.89 | 279.58 | 31660.00 |

**Formatted:** Position: Horizontal: Right, Relative to: Margin, Vertical: 0", Relative to: Paragraph, Wrap Around

| | | | | | | |
|---|---|---|---|---|---|---|
| Mono-isobutyl phthalate | 0.14 | 2.70 | 6.10 | 12.18 | 12.70 | 14537.00 |
| Mono-isononyl phthalate | 0.35 | 0.87 | 0.87 | 2.45 | 1.09 | 454.61 |
| Mono-n-butyl phthalate | 0.28 | 8.00 | 17.40 | 34.45 | 34.70 | 9873.60 |
| Mono-n-methyl phthalate | 0.10 | 0.70 | 0.90 | 5.46 | 3.09 | 10530.00 |
| Mono(carboxynonyl) phthalate | 0.14 | 1.22 | 2.50 | 5.60 | 4.90 | 876.40 |
| Mono(carboxyoctyl) phthalate | 0.14 | 3.90 | 9.39 | 34.72 | 26.60 | 3287.40 |
| NNAL , urine | 0.00 | 0.00 | 0.00 | 0.17 | 0.03 | 890.00 |
| o-Desmethylangolensin (O-DMA) | 0.10 | 0.60 | 3.20 | 79.00 | 20.10 | 21600.00 |
| O-Phenylphenol | 0.07 | 0.07 | 0.07 | 0.21 | 0.14 | 35.40 |
| Oxypyrimidine | 0.07 | 0.07 | 0.07 | 0.31 | 0.49 | 43.13 |
| Paranitrophenol | 0.07 | 0.23 | 0.62 | 1.17 | 1.31 | 59.89 |
| Perchlorate, urine | 0.04 | 1.90 | 3.46 | 5.29 | 6.01 | 583.00 |
| Perfluorodecanoic acid | 0.07 | 0.14 | 0.20 | 0.33 | 0.40 | 25.20 |
| Perfluorohexane sulfonic acid | 0.07 | 0.54 | 1.40 | 2.15 | 2.60 | 82.00 |
| Perfluorononanoic acid | 0.06 | 0.40 | 0.82 | 1.09 | 1.39 | 80.77 |
| Perfluorooctane sulfonamide | 0.07 | 0.07 | 0.07 | 0.14 | 0.10 | 9.60 |
| Perfluorooctane sulfonic acid | 0.14 | 6.40 | 12.30 | 17.37 | 22.40 | 1403.00 |
| Perfluorooctanoic acid | 0.07 | 1.97 | 3.20 | 3.84 | 4.90 | 123.00 |
| Perfluoroundecanoic acid | 0.07 | 0.07 | 0.14 | 0.24 | 0.20 | 28.50 |
| Potassium | 2.30 | 3.80 | 4.00 | 4.00 | 4.20 | 7.30 |
| Propyl paraben | 0.07 | 1.30 | 9.00 | 64.91 | 51.00 | 7210.00 |
| Retinyl palmitate | 0.09 | 0.70 | 1.40 | 1.85 | 2.40 | 58.52 |
| Retinyl stearate | 0.08 | 0.31 | 0.49 | 0.50 | 0.50 | 13.01 |
| Sodium | 99.00 | 138.00 | 139.00 | 139.17 | 141.00 | 160.00 |
| Sum of arsenic metabolites | 1.49 | 2.74 | 4.50 | 6.50 | 7.36 | 384.00 |
| Sum of metabolites of DEHP | 0.86 | 21.04 | 44.88 | 136.78 | 97.73 | 32041.50 |
| Thallium, urine | 0.01 | 0.10 | 0.17 | 0.19 | 0.26 | 2.82 |
| Total Calcium | 6.50 | 9.20 | 9.50 | 9.46 | 9.70 | 14.80 |
| total Lycopene | 0.21 | 27.28 | 39.27 | 41.59 | 53.10 | 175.62 |

**Formatted:** Position: Horizontal: Right, Relative to: Margin, Vertical: 0", Relative to: Paragraph, Wrap Around

| | | | | | | |
|---|---|---|---|---|---|---|
| trans-3-(2,2-dichlorovinyl)-2,2-dimethylcyclopropane carboxylic acid | 0.28 | 0.42 | 0.42 | 1.44 | 0.42 | 614.75 |
| trans-b-carotene | 0.43 | 6.48 | 11.05 | 16.78 | 19.90 | 388.19 |
| trans-lycopene | 0.21 | 14.30 | 20.91 | 22.22 | 28.79 | 110.00 |
| Tungsten, urine | 0.01 | 0.03 | 0.07 | 0.14 | 0.15 | 32.91 |
| Uranium, urine | 0.00 | 0.00 | 0.01 | 0.01 | 0.01 | 1.82 |
| Urinary Arsenobetaine | 0.28 | 0.51 | 1.13 | 9.95 | 5.82 | 1280.00 |
| Urinary Arsenous acid | 0.08 | 0.71 | 0.85 | 0.79 | 0.85 | 156.00 |
| Urinary Benzophenone-3 | 0.20 | 4.20 | 13.60 | 270.28 | 63.55 | 92900.00 |
| Urinary Bisphenol A | 0.14 | 0.90 | 1.90 | 3.95 | 3.80 | 965.00 |
| Urinary Dimethylarsonic acid | 1.20 | 2.11 | 3.71 | 5.57 | 6.30 | 270.00 |
| Urinary Monomethylarsonic acid | 0.14 | 0.63 | 0.64 | 0.93 | 1.04 | 177.00 |
| Urinary nitrate | 494.97 | 26900.00 | 46100.00 | 56638.61 | 72200.00 | 1540000.00 |
| Urinary thiocyanate | 14.00 | 548.00 | 1160.00 | 2419.91 | 2570.00 | 204000.00 |
| Urinary total Arsenic | 0.35 | 4.12 | 8.00 | 19.46 | 16.60 | 1470.00 |
| Urinary Triclosan | 1.20 | 2.40 | 9.40 | 103.22 | 52.65 | 9572.00 |
| Vitamin A | 0.21 | 44.00 | 54.00 | 56.19 | 65.80 | 223.20 |
| Vitamin B12, serum | 18.00 | 364.00 | 486.00 | 619.47 | 655.00 | 240599.00 |
| Vitamin B6 | 2.00 | 26.50 | 43.35 | 67.18 | 74.50 | 2330.00 |
| Vitamin C | 0.01 | 0.64 | 0.97 | 0.96 | 1.24 | 4.83 |
| Vitamin D | 5.49 | 42.20 | 58.50 | 60.65 | 75.30 | 375.00 |

**Formatted:** Position: Horizontal: Right, Relative to: Margin, Vertical: 0", Relative to: Paragraph, Wrap Around

Table S8. Number of participants with a given number of measured chemicals. For example, 716 participants have measurements available for 89 chemical biomarkers.

| Number of measured chemicals | Number of participants |
|---|---|
| 0 | 2969 |
| 1 | 77 |
| 2 | 13 |
| 3 | 6 |
| 4 | 23 |
| 5 | 28 |
| 6 | 11 |
| 7 | 5 |
| 8 | 45 |
| 9 | 361 |
| 10 | 261 |
| 11 | 260 |
| 12 | 177 |
| 13 | 385 |
| 14 | 257 |
| 15 | 216 |
| 16 | 1163 |
| 17 | 194 |
| 18 | 327 |
| 19 | 1591 |
| 20 | 1562 |
| 21 | 605 |
| 22 | 1306 |
| 23 | 437 |
| 24 | 501 |
| 25 | 1486 |
| 26 | 1487 |
| 27 | 366 |
| 28 | 766 |
| 29 | 1678 |
| 30 | 362 |
| 31 | 797 |
| 32 | 260 |

Formatted: Position: Horizontal: Right, Relative to: Margin, Vertical: 0", Relative to: Paragraph, Wrap Around

| | |
|---|---|
| 33 | 1634 |
| 34 | 468 |
| 35 | 339 |
| 36 | 601 |
| 37 | 345 |
| 38 | 364 |
| 39 | 1632 |
| 40 | 316 |
| 41 | 388 |
| 42 | 1090 |
| 43 | 1126 |
| 44 | 108 |
| 45 | 182 |
| 46 | 358 |
| 47 | 404 |
| 48 | 622 |
| 49 | 1809 |
| 50 | 3512 |
| 51 | 1412 |
| 52 | 1185 |
| 53 | 1030 |
| 54 | 1278 |
| 55 | 1987 |
| 56 | 2207 |
| 57 | 327 |
| 58 | 1066 |
| 59 | 68 |
| 60 | 53 |
| 61 | 120 |
| 62 | 222 |
| 63 | 307 |
| 64 | 137 |
| 65 | 163 |
| 66 | 79 |
| 67 | 366 |
| 68 | 915 |
| 69 | 182 |

**Formatted:** Position: Horizontal: Right, Relative to: Margin, Vertical: 0", Relative to: Paragraph, Wrap Around

| | |
|---|---|
| 70 | 229 |
| 71 | 65 |
| 72 | 122 |
| 73 | 102 |
| 74 | 7 |
| 75 | 5 |
| 76 | 46 |
| 77 | 4 |
| 78 | 11 |
| 79 | 20 |
| 80 | 52 |
| 81 | 166 |
| 82 | 185 |
| 83 | 22 |
| 84 | 33 |
| 85 | 23 |
| 86 | 98 |
| 87 | 234 |
| 88 | 484 |
| 89 | 716 |

**Formatted:** Position: Horizontal: Right, Relative to: Margin, Vertical: 0", Relative to: Paragraph, Wrap Around

**Table S9. Description of each hierarchical clustering method.**

| Linkage Method | Description |
|---|---|
| Single (nearest neighbor) | Distance between two clusters is determined by a pair of elements that are closest to each other. At each step, if a pair of elements, with the shortest distance, do not belong in the same cluster, then the two clusters will be combined. |
| Complete (farthest neighbor) | Distance between two clusters is determined by a pair of elements that are farthest from each other. At each step, if a pair of elements, with the shortest distance, do not belong in the same cluster, then the two clusters will be combined. |
| Average (unweighted pair group method with arithmetic mean or UPGMA) | Distance between two clusters is determined by the average of all distances between each pairs of distance in either clusters. |
| McQuitty (Weighted Pair Group Method with Arithmetic Mean or WPGMA) | Distance of two nearest clusters (*i* and *j*) to another cluster (*k*) is the arithmetic mean of average distances between elements of *k* and *i* along with *k* and *j*. |

Formatted: Position: Horizontal: Right, Relative to: Margin, Vertical: 0", Relative to: Paragraph, Wrap Around

**Table S10. Biomonitoring equivalents for noncancer effects.**

| Chemical Name | CAS No. | Biological Medium | Biomonitoring Equivalents | Exposure Guidelines | Website source |
|---|---|---|---|---|---|
| Blood Ethylbenzene (ng/ml) | 100-41-4 | blood | 1 | MRL | https://ehp.niehs.nih.gov/doi/10.1289/ehp.1205740 |
| Blood Styrene (ng/ml) | 100-42-5 | blood | 3 | RfC | https://ehp.niehs.nih.gov/doi/10.1289/ehp.1205740 |
| Blood 1,4-Dichlorobenzene (ng/ml) | 106-46-7 | blood | 3 | RfC | https://ehp.niehs.nih.gov/doi/10.1289/ehp.1205740 |
| Blood m-/p-Xylene (ng/ml) | 108-38-3/106-42-3 | blood | 0.3 | RfC | https://ehp.niehs.nih.gov/doi/10.1289/ehp.1205740 |
| Blood Toluene (ng/mL) | 108-88-3 | blood | 20 | RfC | https://ehp.niehs.nih.gov/doi/10.1289/ehp.1205740 |
| Blood Chlorobenzene (ng/mL) | 108-90-7 | blood | 0.2 | RfD | https://ehp.niehs.nih.gov/doi/10.1289/ehp.1205740 |
| Hexachlorobenzene (ng/g) | 118-74-1 | blood | 47 | MRL | https://ehp.niehs.nih.gov/doi/10.1289/ehp.1205740 |
| Hexachlorobenzene Lipid Adj (ng/g) | 118-74-1 | blood | 47 | MRL | https://ehp.niehs.nih.gov/doi/10.1289/ehp.1205740 |
| Blood Dibromochloromethane (pg/ml) | 124-48-1 | blood | 20 | RfD | https://ehp.niehs.nih.gov/doi/10.1289/ehp.1205740 |
| Blood Tetrachloroethene (ng/mL) | 127-18-4 | blood | 1 | RfD | https://ehp.niehs.nih.gov/doi/10.1289/ehp.1205740 |
| Mono-n-butyl phthalate (ng/mL) | 131-70-4 | urine | 2700 | RfD | https://ehp.niehs.nih.gov/doi/10.1289/ehp.1205740 |
| Blood cis-1,2-Dichloroethene (ng/mL) | 156-59-2 | blood | 0.034 | RfD | https://ehp.niehs.nih.gov/doi/10.1289/ehp.1205740 |
| Blood trans-1,2-Dichloroethene (ng/mL) | 156-60-5 | blood | 0.07 | RfD | https://ehp.niehs.nih.gov/doi/10.1289/ehp.1205740 |
| Blood MTBE (pg/ml) | 1634-04-04 | blood | 20 | RfC | https://ehp.niehs.nih.gov/doi/10.1289/ehp.1205740 |
| Mono-ethyl phthalate (ng/mL) | 2306-33-4 | urine | 18000 | RfD | https://ehp.niehs.nih.gov/doi/10.1289/ehp.1205740 |
| Mono-benzyl phthalate (ng/mL) | 2528-16-7 | urine | 3800 | RfD | https://ehp.niehs.nih.gov/doi/10.1289/ehp.1205740 |
| Mono(carboxynonyl) phthalate (ng/mL) | 26761-40-0 | urine | 390 | ADI | https://ehp.niehs.nih.gov/doi/10.1289/ehp.1205740 |

**Formatted:** Position: Horizontal: Right, Relative to: Margin, Vertical: 0", Relative to: Paragraph, Wrap Around

| | | | | | |
|---|---|---|---|---|---|
| Uriry Triclosan (ng/mL) | 3380-34-5 | urine | 6400 | RfD | https://ehp.niehs.nih.gov/doi/10.1289/ehp.1205740 |
| Uriry Triclosan (ng/mL) | 3380-34-5 | urine | 1783 | Acceptable daily intake | https://www.sciencedirect.com/science/article/pii/S1438463919304626 |
| 3-phenoxybenzoic acid (ug/L) | 3739-38-6 | urine | 1.7 | RfD | https://www.sciencedirect.com/science/article/pii/S1438463919304626 |
| cis-3-(2,2-dichlorovinyl)-2,2-dimethylcyclopropane carboxylic acid (ug/L) | 55701-05-08 | urine | 50 | RfD | https://ehp.niehs.nih.gov/doi/10.1289/ehp.1205740 |
| Blood Carbon Tetrachloride (ng/ml) | 56-23-5 | blood | 0.19 | RfC | https://ehp.niehs.nih.gov/doi/10.1289/ehp.1205740 |
| Glycideamide (pmol/g Hb) | 5694-00-8 | blood | 176 | RfD | https://www.sciencedirect.com/science/article/pii/S1438463919304626 |
| cis-3-(2,2-dibromovinyl)-2,2-dimethylcyclopropane carboxylic acid (ug/L) | 63597-73-9 | urine | 20 | Acceptable daily intake | https://www.sciencedirect.com/science/article/pii/S1438463919304626 |
| 3,5,6-trichloropyridinol (ug/L) | 6515-38-4 | urine | 2100 | biomonitoring guidance value | https://www.sciencedirect.com/science/article/pii/S1438463919304626 |
| Blood Chloroform (pg/ml) | 67-66-3 | blood | 230 | RfD | https://ehp.niehs.nih.gov/doi/10.1289/ehp.1205740 |
| Blood Hexachloroethane (ng/mL) | 67-72-1 | blood | 0.2 | RfC | https://ehp.niehs.nih.gov/doi/10.1289/ehp.1205740 |
| Blood Benzene (ng/ml) | 71-43-2 | blood | 0.15 | RfC | https://ehp.niehs.nih.gov/doi/10.1289/ehp.1205740 |
| Blood 1,1,1-Trichloroethane (ng/mL) | 71-55-6 | blood | 20 | RfC | https://ehp.niehs.nih.gov/doi/10.1289/ehp.1205740 |
| Blood lead (ug/dL) | 7439-92-1 | blood | 10 | | https://www.cdc.gov/biomonitoring/Lead_BiomonitoringSummary.html |
| Blood mercury, total (ug/L) | 7439-97-6 | blood | 5.8 | NRC benchmark concentration assessment | https://ehp.niehs.nih.gov/doi/10.1289/ehp.1205740 |
| Molybdenum, urine (ng/mL) | 7439-98-7 | urine | 7516 | NOAEL + Uncertainty Factor | https://www.sciencedirect.com/science/article/pii/S1438463919304626 |
| Thallium, urine (ng/mL) | 7440-28-0 | urine | 5 | occupatiol biomonitoring data | https://ehp.niehs.nih.gov/doi/10.1289/ehp.1205740 |
| Blood cadmium (ug/L) | 7440-43-9 | blood | 1.7 | RfD | https://www.sciencedirect.com/science/article/pii/S1438463919304626 |

**Formatted:** Position: Horizontal: Right, Relative to: Margin, Vertical: 0", Relative to: Paragraph, Wrap Around

| | | | | | |
|---|---|---|---|---|---|
| Cadmium, urine (ng/mL) | 7440-43-9 | urine | 1.5 | RfD | https://ehp.niehs.nih.gov/doi/10.1289/ehp.1205740 |
| Blood Bromoform (pg/ml) | 75-25-2 | blood | 130 | RfD | https://ehp.niehs.nih.gov/doi/10.1289/ehp.1205740 |
| Blood Bromodichloromethane (pg/ml) | 75-27-4 | blood | 80 | RfD | https://ehp.niehs.nih.gov/doi/10.1289/ehp.1205740 |
| Blood 1,1-Dichloroethene (ng/mL) | 75-35-4 | blood | 0.3 | RfC | https://ehp.niehs.nih.gov/doi/10.1289/ehp.1205740 |
| 4-fluoro-3-phenoxybenzoic acid (ug/L) | 77279-89-1 | urine | 240 | RfD | https://ehp.niehs.nih.gov/doi/10.1289/ehp.1205740 |
| 4-fluoro-3-phenoxybenzoic acid (ug/L) | 77279-89-1 | urine | 46 | Acceptable daily intake | https://www.sciencedirect.com/science/article/pii/S1438463919304626 |
| Selenium (ug/L) | 7782-49-2 | blood | 480 | Uncertain factor | https://www.sciencedirect.com/science/article/pii/S1438463919304626 |
| Blood 1,2-Dichloropropane (ng/mL) | 78-87-5 | blood | 0.01 | RfC | https://ehp.niehs.nih.gov/doi/10.1289/ehp.1205740 |
| Blood 1,1,2-Trichloroethane (ng/mL) | 79-00-5 | blood | 0.05 | RfD | https://ehp.niehs.nih.gov/doi/10.1289/ehp.1205740 |
| Blood Trichloroethene (ng/ml) | 79-01-6 | blood | 0.0062 | RfC | https://ehp.niehs.nih.gov/doi/10.1289/ehp.1205740 |
| Acrylamide (pmoL/g Hb) | 79-06-1 | blood | 190 | RfD | https://www.sciencedirect.com/science/article/pii/S1438463919304626 |
| Blood 1,1,2,2-Tetrachloroethane (ng/mL) | 79-34-5 | blood | 0.2 | RfD | https://ehp.niehs.nih.gov/doi/10.1289/ehp.1205740 |
| Uriry Bisphenol A (ng/mL) | 80-05-7 | urine | 2000 | RfD | https://ehp.niehs.nih.gov/doi/10.1289/ehp.1205740 |
| Pentachlorophenol (ug/L) | 87-86-5 | urine | 25 | occupatiol biomonitoring data | https://ehp.niehs.nih.gov/doi/10.1289/ehp.1205740 |
| 2,4-D (ug/L) | 94-75-7 | urine | 2000 | RfD | https://ehp.niehs.nih.gov/doi/10.1289/ehp.1205740 |
| Blood o-Xylene (ng/mL) | 95-47-6 | blood | 0.3 | RfC | https://ehp.niehs.nih.gov/doi/10.1289/ehp.1205740 |
| Blood 1,2-Dichlorobenzene (ng/mL) | 95-50-1 | blood | 0.7 | RfD | https://ehp.niehs.nih.gov/doi/10.1289/ehp.1205740 |
| Blood 1,2-Dibromo-3-chloropropane (ng/mL) | 96-12-8 | blood | 0.001 | RfC | https://ehp.niehs.nih.gov/doi/10.1289/ehp.1205740 |
| Blood Nitrobenzene (ng/mL) | 98-95-3 | blood | 0.03 | RfC | https://ehp.niehs.nih.gov/doi/10.1289/ehp.1205740 |

**Formatted:** Position: Horizontal: Right, Relative to: Margin, Vertical: 0", Relative to: Paragraph, Wrap Around

| | | | | |
|---|---|---|---|---|
| Sum of metabolites of DEHP | urine | 400 | RfD | https://ehp.niehs.nih.gov/doi/10.1289/ehp.1205740 |
| Sum of arsenic metabolites | urine | 5.8 | RfD | https://ehp.niehs.nih.gov/doi/10.1289/ehp.1205740 |

**Formatted:** Position: Horizontal: Right, Relative to: Margin, Vertical: 0", Relative to: Paragraph, Wrap Around

Table S11. Cancer and intake slope factors used to calculate biomonitoring equivalents for cancer effects via inhalation or ingestion. Biomonitoring equivalents are calculated with the following formula: $\frac{extra\ cancer\ risk}{cancer\ slope\ factor} \times intake\ slope\ factor$. Extra cancer risk can be 10E-4, 10E-5, 10E-6.

| Chemical Name | CAS No. | Biological medium | Intake type | Cancer slope factor | Intake slope factor | Source for cancer slope factor | Source for intake slope factor |
|---|---|---|---|---|---|---|---|
| Blood 1,1-Dichloroethane (ng/mL) | 75-34-3 | blood | ingestion | 0 | 2.6 | USEtox | https://pubmed.ncbi.nlm.nih.gov/20685286/ |
| Blood 1,1-Dichloroethene (ng/mL) | 75-35-4 | blood | ingestion | 0 | 1.1 | USEtox | https://pubmed.ncbi.nlm.nih.gov/20685286/ |
| Blood 1,1,1-Trichloroethane (ng/mL) | 71-55-6 | blood | ingestion | 0 | 48 | USEtox | https://pubmed.ncbi.nlm.nih.gov/20685286/ |
| Blood 1,1,1,2-tetrachloroethane (ng/mL) | 630-20-6 | blood | ingestion | 0.020068681 | 18 | USEtox | https://pubmed.ncbi.nlm.nih.gov/20685286/ |
| Blood 1,1,2-Trichloroethane (ng/mL) | 79-00-5 | blood | ingestion | 0.066409091 | 13 | USEtox | https://pubmed.ncbi.nlm.nih.gov/20685286/ |
| Blood 1,1,2,2-Tetrachloroethane (ng/mL) | 79-34-5 | blood | ingestion | 0.095365535 | 9 | USEtox | https://pubmed.ncbi.nlm.nih.gov/20685286/ |
| Blood 1,2-dibromoethane (ng/ml) | 106-93-4 | blood | ingestion | 1.434548328 | 12 | USEtox | https://pubmed.ncbi.nlm.nih.gov/20685286/ |
| Blood 1,2,3-trichloropropane (ng/ml) | 96-18-4 | blood | ingestion | 4.174285714 | 14 | USEtox | https://pubmed.ncbi.nlm.nih.gov/20685286/ |
| Blood 1,4-Dioxane (ng/mL) | 123-91-1 | blood | ingestion | 0.017904412 | 70.2 | USEtox | https://pubmed.ncbi.nlm.nih.gov/20685286/ |
| Blood Benzene (ng/ml) | 71-43-2 | blood | ingestion | 0.026 | 34 | Usetox | https://pubmed.ncbi.nlm.nih.gov/20685286/ |
| Blood Bromodichloromethane (pg/ml) | 75-27-4 | blood | ingestion | 0.077 | 5000 | USEtox | https://pubmed.ncbi.nlm.nih.gov/20685286/ |
| Blood Bromoform (pg/ml) | 75-25-2 | blood | ingestion | 0.003165747 | 5800 | USEtox | https://pubmed.ncbi.nlm.nih.gov/20685286/ |
| Blood Carbon Tetrachloride (ng/ml) | 56-23-5 | blood | ingestion | 0.02435 | 16 | USEtox | https://pubmed.ncbi.nlm.nih.gov/20685286/ |
| Blood Chloroform (pg/ml) | 67-66-3 | blood | ingestion | 0.040448505 | 4100 | USEtox | https://pubmed.ncbi.nlm.nih.gov/20685286/ |

**Formatted:** Position: Horizontal: Right, Relative to: Margin, Vertical: 0", Relative to: Paragraph, Wrap Around

| | | | | | | | |
|---|---|---|---|---|---|---|---|
| Blood Dibromochloromethane (pg/ml) | 124-48-1 | blood | ingestion | 0.026276978 | 7400 | USEtox | https://pubmed.ncbi.nlm.nih.gov/20685286/ |
| Blood Ethylbenzene (ng/ml) | 100-41-4 | blood | ingestion | 0.042209961 | 21 | USEtox | https://pubmed.ncbi.nlm.nih.gov/20685286/ |
| Blood furan (ng/mL) | 110-00-9 | blood | ingestion | 5.180313408 | 39 | USEtox | https://pubmed.ncbi.nlm.nih.gov/20685286/ |
| Blood Hexachloroethane (ng/mL) | 67-72-1 | blood | ingestion | 0.037028955 | 52 | USEtox | https://pubmed.ncbi.nlm.nih.gov/20685286/ |
| Blood Hexane (ng/mL) | 110-54-3 | blood | ingestion | 1.21E-04 | 8 | USEtox | https://pubmed.ncbi.nlm.nih.gov/20685286/ |
| Blood Methylene Chloride (ng/mL) | 75-09-2 | blood | ingestion | 0.004690711 | 12 | USEtox | https://pubmed.ncbi.nlm.nih.gov/20685286/ |
| Blood Styrene (ng/ml) | 100-42-5 | blood | ingestion | 0 | 5.4 | USEtox | https://pubmed.ncbi.nlm.nih.gov/20685286/ |
| Blood Tetrachloroethene (ng/mL) | 127-18-4 | blood | ingestion | 0.039022436 | 120 | USEtox | https://pubmed.ncbi.nlm.nih.gov/20685286/ |
| Blood Tetrahydrofuran (ng/mL) | 109-99-9 | blood | ingestion | 0.005040305 | 0.5 | USEtox | https://pubmed.ncbi.nlm.nih.gov/20685286/ |
| Blood Toluene (ng/mL) | 108-88-3 | blood | ingestion | 6.70E-04 | 11 | USEtox | https://pubmed.ncbi.nlm.nih.gov/20685286/ |
| Blood Trichloroethene (ng/ml) | 79-01-6 | blood | ingestion | 0.005285818 | 15 | USEtox | https://pubmed.ncbi.nlm.nih.gov/20685286/ |
| N-acetyl-S-(dimethylphenyl)-L-cysteine (ng/mL) | 1330-20-7 | urine | ingestion | 6.60E-04 | 3.3 | USEtox | https://pubmed.ncbi.nlm.nih.gov/20685286/ |
| Blood 1,1-Dichloroethene (ng/mL) | 75-35-4 | blood | inhalation | 0.105563584 | 1.3 | USEtox | https://pubmed.ncbi.nlm.nih.gov/20685286/ |
| Blood 1,1,1,2-tetrachloroethane (ng/mL) | 630-20-6 | blood | inhalation | 0.020068681 | 4.1 | USEtox | https://pubmed.ncbi.nlm.nih.gov/20685286/ |
| Blood 1,1,2-Trichloroethane (ng/mL) | 79-00-5 | blood | inhalation | 0.066409091 | 3.7 | USEtox | https://pubmed.ncbi.nlm.nih.gov/20685286/ |
| Blood 1,1,2,2-Tetrachloroethane (ng/mL) | 79-34-5 | blood | inhalation | 0.095365535 | 3.5 | USEtox | https://pubmed.ncbi.nlm.nih.gov/20685286/ |
| Blood 1,2-dibromoethane (ng/ml) | 106-93-4 | blood | inhalation | 1.298357031 | 3.8 | USEtox | https://pubmed.ncbi.nlm.nih.gov/20685286/ |
| Blood 1,2,3-trichloropropane (ng/ml) | 96-18-4 | blood | inhalation | 4.174285714 | 1.6 | USEtox | https://pubmed.ncbi.nlm.nih.gov/20685286/ |

**Formatted:** Position: Horizontal: Right, Relative to: Margin, Vertical: 0", Relative to: Paragraph, Wrap Around

| | | | | | | | |
|---|---|---|---|---|---|---|---|
| Blood 1,4-Dioxane (ng/mL) | 123-91-1 | blood | inhalation | 0.017904412 | 9.9 | USEtox | https://pubmed.ncbi.nlm.nih.gov/20685286/ |
| Blood Benzene (ng/ml) | 71-43-2 | blood | inhalation | 0.026 | 4.2 | Usetox | https://pubmed.ncbi.nlm.nih.gov/20685286/ |
| Blood Bromoform (pg/ml) | 75-25-2 | blood | inhalation | 0.003165747 | 3200 | USEtox | https://pubmed.ncbi.nlm.nih.gov/20685286/ |
| Blood Carbon Tetrachloride (ng/ml) | 56-23-5 | blood | inhalation | 0.193253968 | 1.9 | USEtox | https://pubmed.ncbi.nlm.nih.gov/20685286/ |
| Blood Chloroform (pg/ml) | 67-66-3 | blood | inhalation | 0.010089779 | 2400 | USEtox | https://pubmed.ncbi.nlm.nih.gov/20685286/ |
| Blood Dibromochloromethane (pg/ml) | 124-48-1 | blood | inhalation | 0.026276978 | 3200 | USEtox | https://pubmed.ncbi.nlm.nih.gov/20685286/ |
| Blood Ethylbenzene (ng/ml) | 100-41-4 | blood | inhalation | 0.042209961 | 4.3 | USEtox | https://pubmed.ncbi.nlm.nih.gov/20685286/ |
| Blood furan (ng/mL) | 110-00-9 | blood | inhalation | 5.180313408 | 4.4 | USEtox | https://pubmed.ncbi.nlm.nih.gov/20685286/ |
| Blood Hexachloroethane (ng/mL) | 67-72-1 | blood | inhalation | 0.037028955 | 7.5 | USEtox | https://pubmed.ncbi.nlm.nih.gov/20685286/ |
| Blood Hexane (ng/mL) | 110-54-3 | blood | inhalation | 1.21E-04 | 0.7 | USEtox | https://pubmed.ncbi.nlm.nih.gov/20685286/ |
| Blood Methylene Chloride (ng/mL) | 75-09-2 | blood | inhalation | 0.003320455 | 2.8 | USEtox | https://pubmed.ncbi.nlm.nih.gov/20685286/ |
| Blood Styrene (ng/ml) | 100-42-5 | blood | inhalation | 0.088043095 | 3 | USEtox | https://pubmed.ncbi.nlm.nih.gov/20685286/ |
| Blood Tetrachloroethene (ng/mL) | 127-18-4 | blood | inhalation | 0.01521875 | 11 | USEtox | https://pubmed.ncbi.nlm.nih.gov/20685286/ |
| Blood Tetrahydrofuran (ng/mL) | 109-99-9 | blood | inhalation | 0.005040305 | 2.7 | USEtox | https://pubmed.ncbi.nlm.nih.gov/20685286/ |
| Blood Toluene (ng/mL) | 108-88-3 | blood | inhalation | 0 | 3.1 | USEtox | https://pubmed.ncbi.nlm.nih.gov/20685286/ |
| Blood Trichloroethene (ng/ml) | 79-01-6 | blood | inhalation | 0.003070964 | 3.1 | USEtox | https://pubmed.ncbi.nlm.nih.gov/20685286/ |
| N-acetyl-S-(dimethylphenyl)-L-cysteine (ng/mL) | 1330-20-7 | urine | inhalation | 6.60E-04 | 2.8 | USEtox | https://pubmed.ncbi.nlm.nih.gov/20685286/ |

Formatted: Position: Horizontal: Right, Relative to: Margin, Vertical: 0", Relative to: Paragraph, Wrap Around

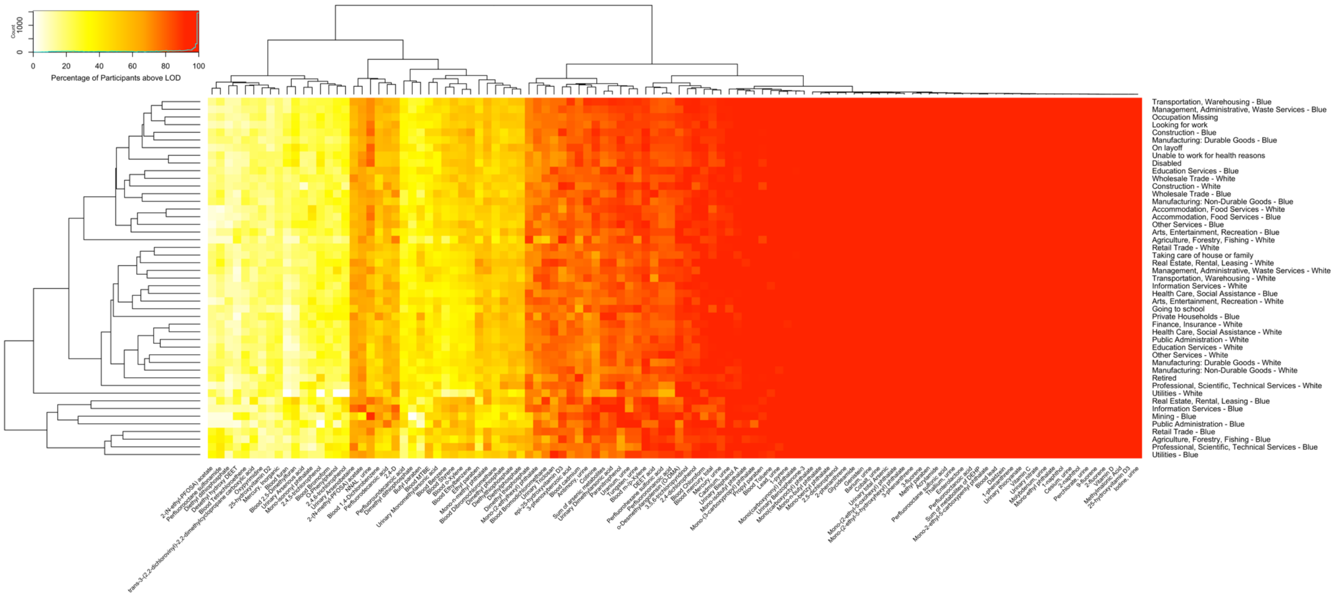

Figure S1. Heatmap of detection frequency by chemical biomarker and occupational group. Both chemical biomarkers and occupational groups are clustered by hierarchical clustering using Euclidean distance.

**Formatted:** Position: Horizontal: Right, Relative to: Margin, Vertical: 0", Relative to: Paragraph, Wrap Around

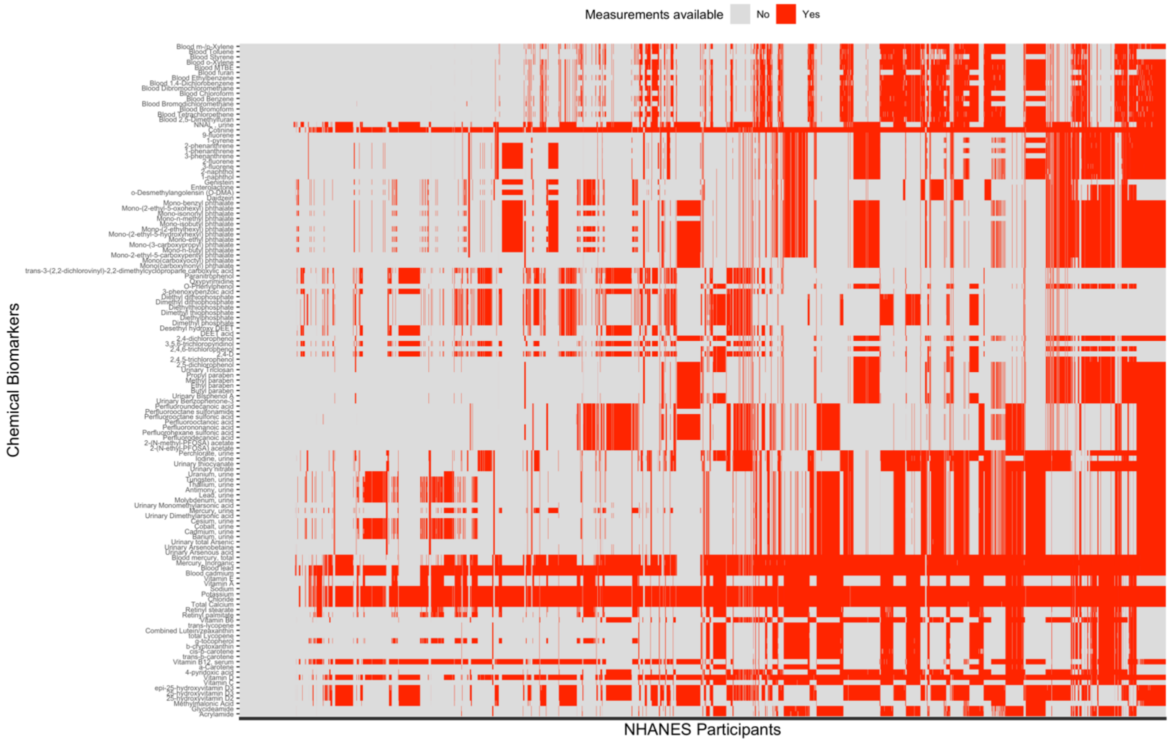

Figure S2. Heatmap of dichotomized biomarker measurements by participants and included chemicals to show sparsity of the chemical biomarker dataset. NHANES participants have data available for age, sex, race/ethnicity, poverty income ratio, smoking, study period, and occupational data. Chemical biomarkers are grouped by chemical class. Participants are order by number of measured chemicals, which ranges from 0 to 89 chemicals, e.g. participants to the far-right have measurements available for 89 chemical biomarkers.

**Formatted:** Position: Horizontal: Right, Relative to: Margin, Vertical: 0", Relative to: Paragraph, Wrap Around

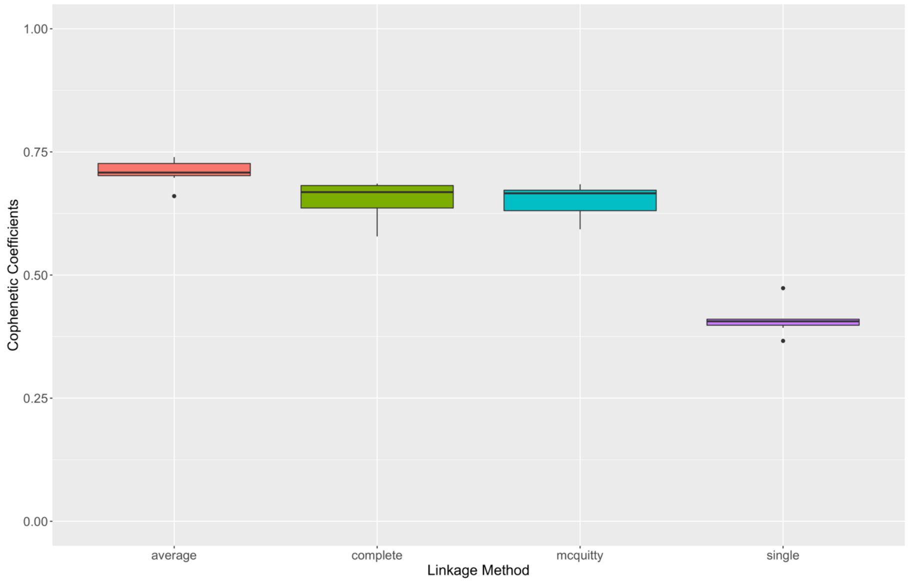

Figure S3. Boxplot of cophenetic correlation coefficients by linkage methods for clustering the occupational groups based on chemical exposure profiles. A cophenetic coefficient is calculated for each stepwise regression models with log10-transformed chemical biomarker levels as the outcome variable and the main predictor as the occupational groups. Covariates include age (continuous), sex (categorical), race (categorical), poverty income ratio (continuous), NHANES cycle (continuous), and smoking with serum cotinine levels (continuous).

**Formatted:** Position: Horizontal: Right, Relative to: Margin, Vertical: 0", Relative to: Paragraph, Wrap Around

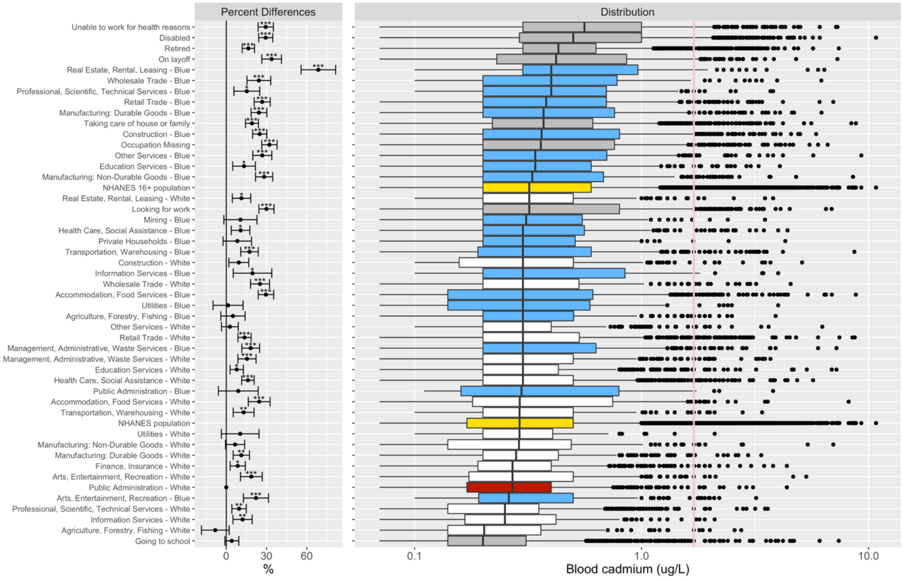

Figure S4. Box plot of distribution of blood cadmium. The "NHANES Population" consists of participants in 1999-2014. The "NHANES 16+ Population" consists of participants in 1999-2014 and are 16 years old or older. Percent differences are derived from fully adjusted models, which were adjusted for age, sex, race/ethnicity, poverty income ratio, study period, and serum cotinine (biomarker of smoking). Reference group for the occupational groups is comprised of white collars from Public Administration. Number of asterisks indicate statistical significance of the percent differences: * (p-value ∈ (0.01, 0.05]), ** (p-value ∈ (0.001, 0.01]), and *** (p-value ≤ 0.001). The p-values corrected for multiple comparison with the Benjamini and Hochberg FDR procedure of 5%.

**Formatted:** Position: Horizontal: Right, Relative to: Margin, Vertical: 0", Relative to: Paragraph, Wrap Around

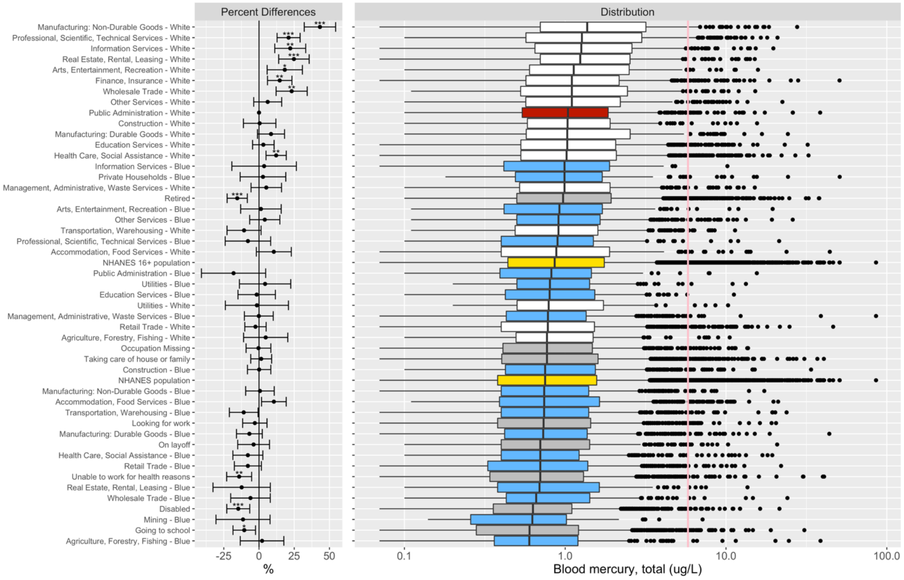

Figure S5. Box plot of distribution of total mercury in blood. The "NHANES Population" consists of participants in 1999-2014. The "NHANES 16+ Population" consists of participants in 1999-2014 and are 16 years old or older. Percent differences are derived from fully adjusted models, which were adjusted for age, sex, race/ethnicity, poverty income ratio, study period, and serum cotinine (biomarker of smoking). Reference group for the occupational groups is comprised of white collars from Public Administration. Number of asterisks indicate statistical significance of the percent differences: * (p-value ∈ (0.01, 0.05]), ** (p-value ∈ (0.001, 0.01]), and *** (p-value ≤ 0.001). The p-values corrected for multiple comparison with the Benjamini and Hochberg FDR procedure of 5%.

**Formatted:** Position: Horizontal: Right, Relative to: Margin, Vertical: 0", Relative to: Paragraph, Wrap Around

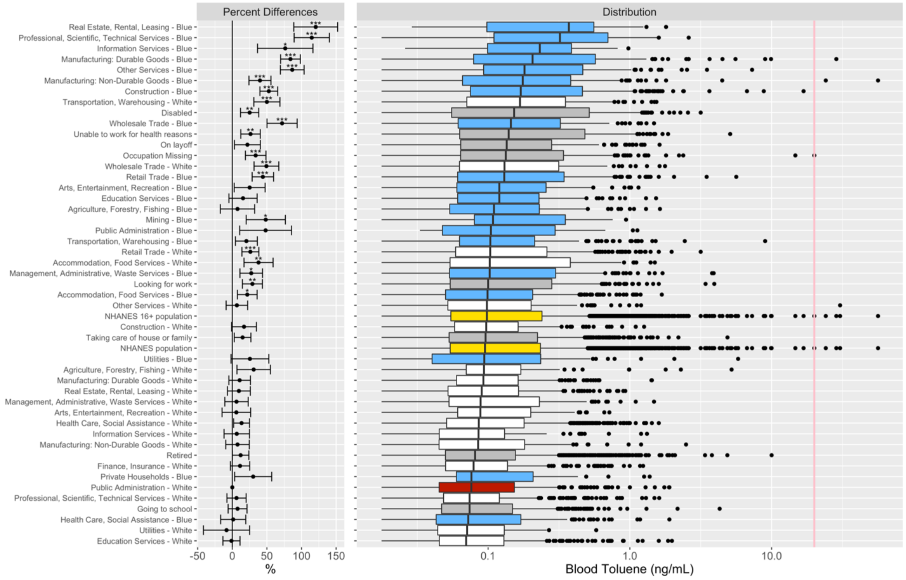

Figure S6. Box plot of distribution of toluene in blood. The “NHANES Population” consists of participants in 1999-2014. The “NHANES 16+ Population” consists of participants in 1999-2014 and are 16 years old or older. Percent differences are derived from fully adjusted models, which were adjusted for age, sex, race/ethnicity, poverty income ratio, study period, and serum cotinine (biomarker of smoking). Reference group for the occupational groups is comprised of white collars from Public Administration. Number of asterisks indicate statistical significance of the percent differences: * (p-value ∈ (0.01, 0.05]), ** (p-value ∈ (0.001, 0.01]), and *** (p-value ≤ 0.001). The p-values corrected for multiple comparison with the Benjamini and Hochberg FDR procedure of 5%.

**Formatted:** Position: Horizontal: Right, Relative to: Margin, Vertical: 0", Relative to: Paragraph, Wrap Around

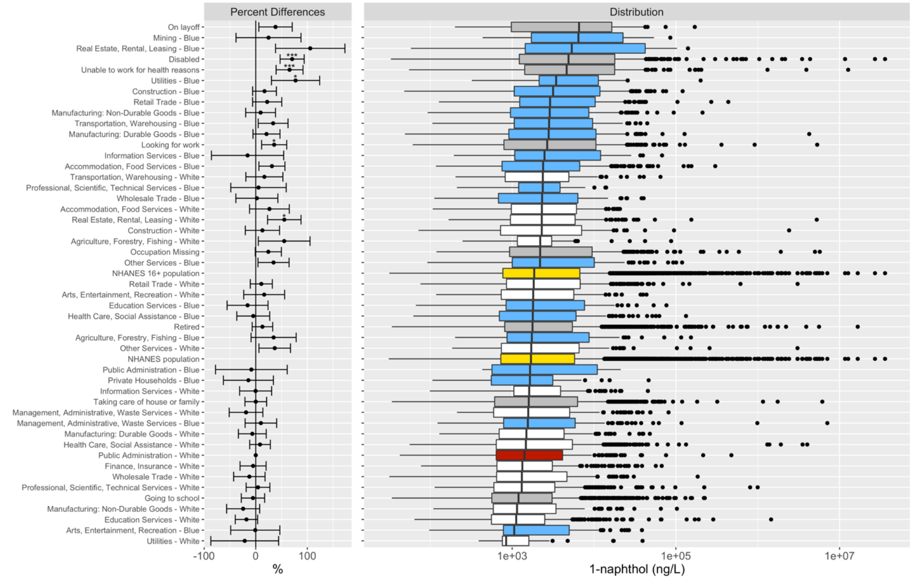

Figure S7. Box plot of distribution of 1-napthol in blood. The “NHANES Population” consists of participants in 1999-2014. The “NHANES 16+ Population” consists of participants in 1999-2014 and are 16 years old or older. Percent differences are derived from fully adjusted models, which were adjusted for age, sex, race/ethnicity, poverty income ratio, study period, and serum cotinine (biomarker of smoking). Reference group for the occupational groups is comprised of white collars from Public Administration. Number of asterisks indicate statistical significance of the percent differences: * (p-value ∈ (0.01, 0.05]), ** (p-value ∈ (0.001, 0.01]), and *** (p-value ≤ 0.001). The p-values corrected for multiple comparison with the Benjamini and Hochberg FDR procedure of 5%.

**Formatted:** Position: Horizontal: Right, Relative to: Margin, Vertical: 0", Relative to: Paragraph, Wrap Around

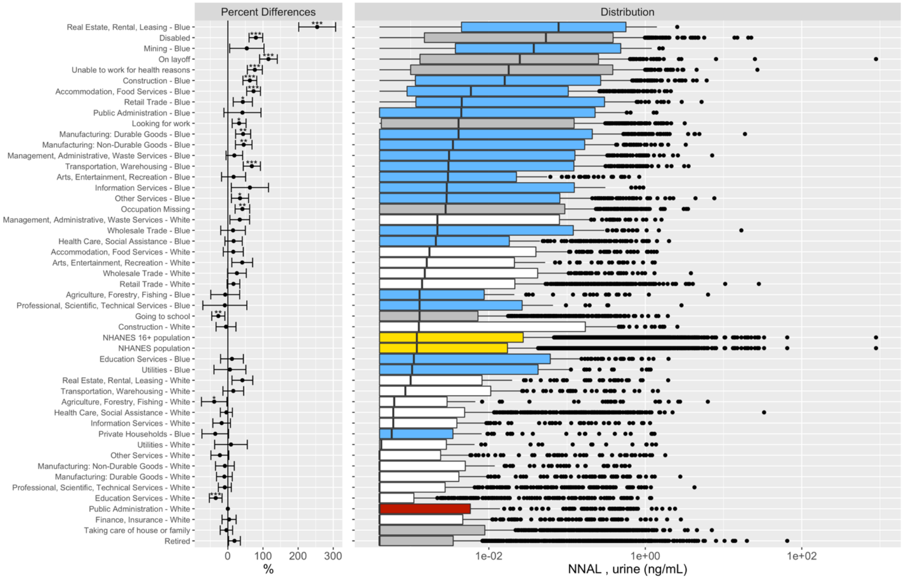

Figure S8. Box plot of distribution of NNAL in urine. The "NHANES Population" consists of participants in 1999-2014. The "NHANES 16+ Population" consists of participants in 1999-2014 and are 16 years old or older. Percent differences are derived from fully adjusted models, which were adjusted for age, sex, race/ethnicity, poverty income ratio, and study period. Reference group for the occupational groups is comprised of white collars from Public Administration. Number of asterisks indicate statistical significance of the percent differences: * (p-value $\in$ (0.01, 0.05]), ** (p-value $\in$ (0.001, 0.01]), and *** (p-value $\leq$ 0.001). The p-values corrected for multiple comparison with the Benjamini and Hochberg FDR procedure of 5%.

**Formatted:** Position: Horizontal: Right, Relative to: Margin, Vertical: 0", Relative to: Paragraph, Wrap Around

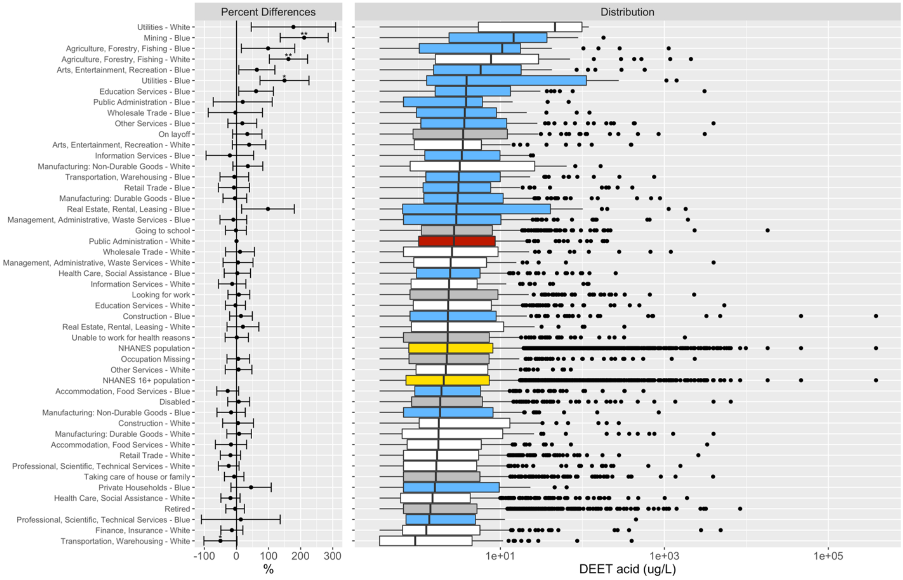

Figure S9. Box plot of distribution of DEET acid in urine. The “NHANES Population” consists of participants in 1999-2014. The “NHANES 16+ Population” consists of participants in 1999-2014 and are 16 years old or older. Percent differences are derived from fully adjusted models, which were adjusted for age, sex, race/ethnicity, poverty income ratio, study period, and serum cotinine (biomarker of smoking). Reference group for the occupational groups is comprised of white collars from Public Administration. Number of asterisks indicate statistical significance of the percent differences: * (p-value ∈ (0.01, 0.05]), ** (p-value ∈ (0.001, 0.01]), and *** (p-value ≤ 0.001). The p-values corrected for multiple comparison with the Benjamini and Hochberg FDR procedure of 5%.

**Formatted:** Position: Horizontal: Right, Relative to: Margin, Vertical: 0", Relative to: Paragraph, Wrap Around

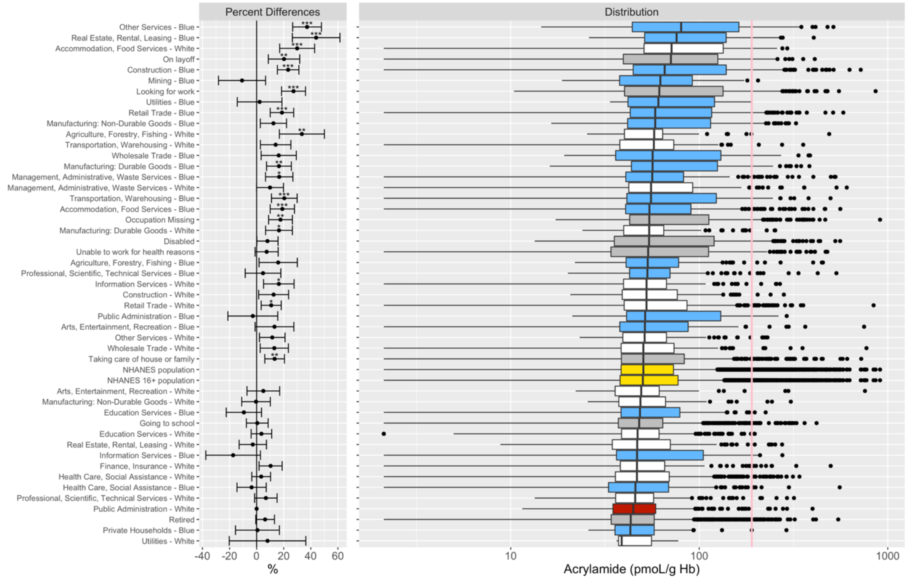

Figure S10. Box plot of distribution of acrylamide in urine. The “NHANES Population” consists of participants in 1999-2014. The “NHANES 16+ Population” consists of participants in 1999-2014 and are 16 years old or older. Percent differences are derived from fully adjusted models, which were adjusted for age, sex, race/ethnicity, poverty income ratio, study period, and serum cotinine (biomarker of smoking). Reference group for the occupational groups is comprised of white collars from Public Administration. Number of asterisks indicate statistical significance of the percent differences: * (p-value ∈ (0.01, 0.05]), ** (p-value ∈ (0.001, 0.01]), and *** (p-value ≤ 0.001). The p-values corrected for multiple comparison with the Benjamini and Hochberg FDR procedure of 5%.

**Formatted:** Position: Horizontal: Right, Relative to: Margin, Vertical: 0", Relative to: Paragraph, Wrap Around

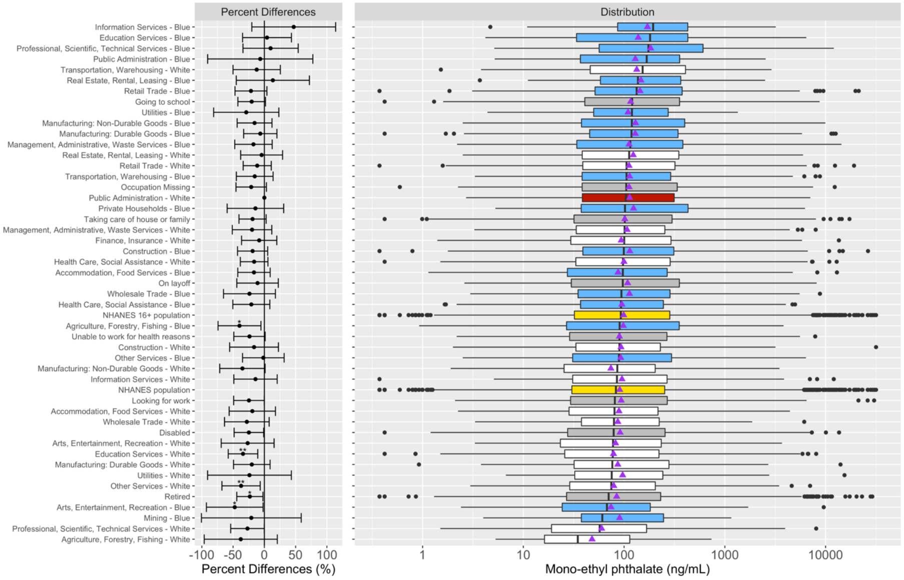

Figure S11. Box plot of distribution of mono-ethyl phthalate in urine. The "NHANES Population" consists of participants in 1999-2014. The "NHANES 16+ Population" consists of participants in 1999-2014 and are 16 years old or older. Percent differences are derived from fully adjusted models, which were adjusted for age, sex, race/ethnicity, poverty income ratio, study period, and serum cotinine (biomarker of smoking). Reference group for the occupational groups is comprised of white collars from Public Administration. Number of asterisks indicate statistical significance of the percent differences: * (p-value ∈ (0.01, 0.05]), ** (p-value ∈ (0.001, 0.01]), and *** (p-value ≤ 0.001). The p-values corrected for multiple comparison with the Benjamini and Hochberg FDR procedure of 5%.

**Formatted:** Position: Horizontal: Right, Relative to: Margin, Vertical: 0", Relative to: Paragraph, Wrap Around

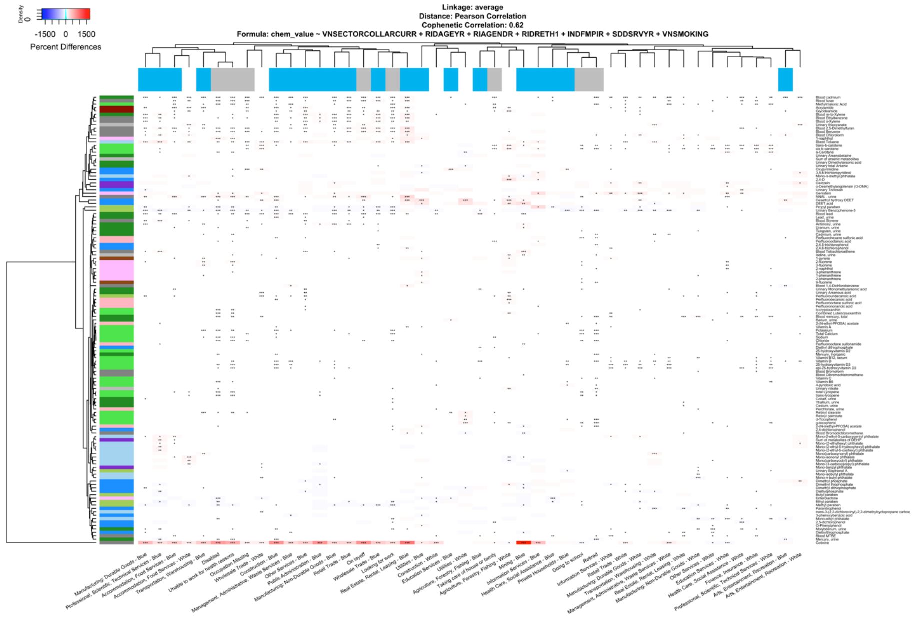

Figure S12. Heatmap of percent differences in chemical biomarker concentrations by occupational group, relative to white collars from Public Administration. Chemical biomarkers in white color indicates that the concentrations are the same between the given sector-collar combination and the reference group. The color bar for the columns represents the collar categorization and unemployment. Blue presents the blue-collar workers. White represents the white-collar workers. Gray presents the unemployed participants. Results are adjusted for age, sex, race/ethnicity, poverty income ratio, study period, and serum cotinine (biomarker of smoking). Results for cotinine is not adjusted for smoking. Number of asterisks indicate statistical significance of the percent differences: * (p-value $\in$ (0.01, 0.05]), ** (p-value $\in$ (0.001, 0.01]), and *** (p-value $\leq$ 0.001).

**Formatted:** Position: Horizontal: Right, Relative to: Margin, Vertical: 0", Relative to: Paragraph, Wrap Around

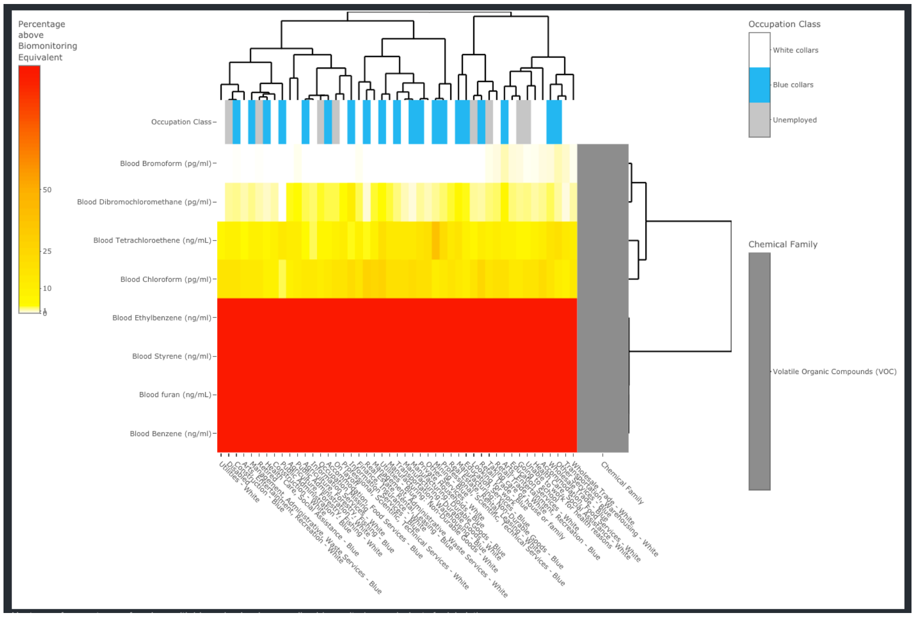

Figure S13. Heatmap of percentages of workers with biomarker levels exceeding biomonitoring equivalents for cancer effects via inhalation. The calculations for the biomonitoring equivalents are based on a 1/10,000 extra risk for cancer.

**Formatted:** Position: Horizontal: Right, Relative to: Margin, Vertical: 0", Relative to: Paragraph, Wrap Around

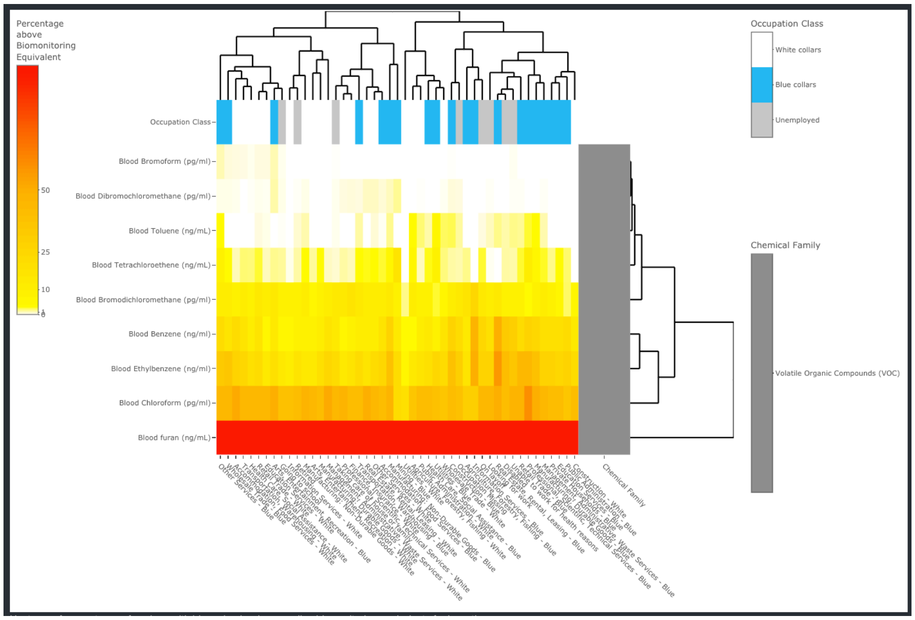

Figure S14. Heatmap of percentages of workers with biomarker levels exceeding biomonitoring equivalents for cancer effects via ingestion. The calculations for the biomonitoring equivalents are based on a 1/10,000 extra risk for cancer.

**Formatted:** Position: Horizontal: Right, Relative to: Margin, Vertical: 0", Relative to: Paragraph, Wrap Around

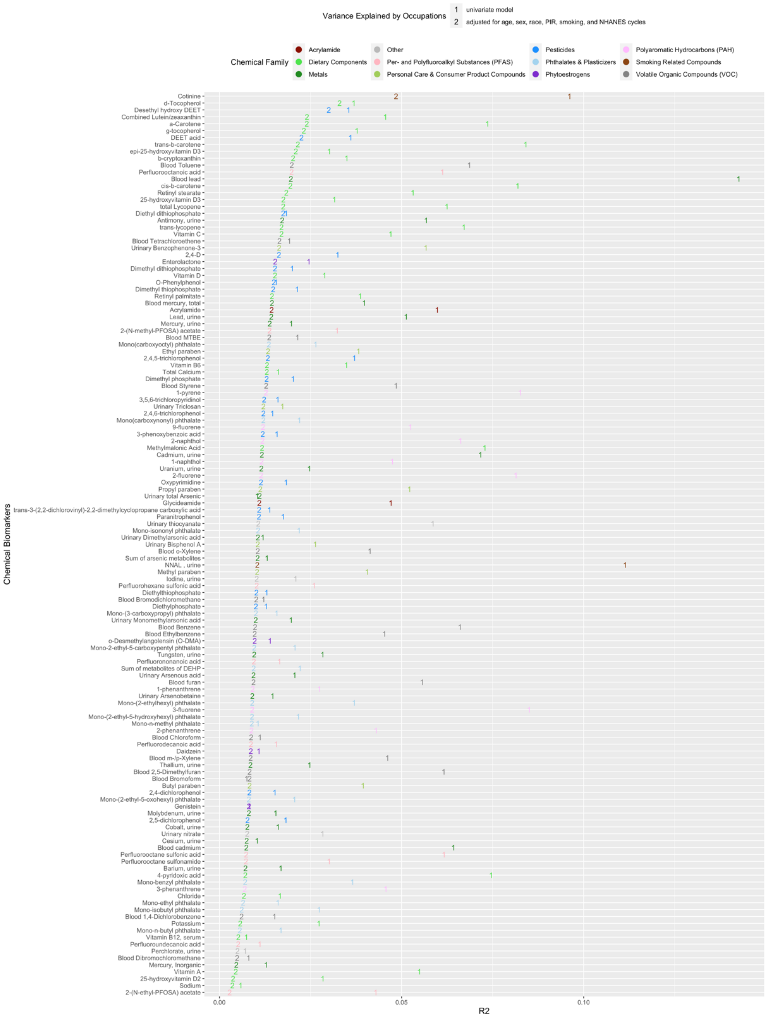

Figure S15. Alphabet soup plot of variance explained by occupation alone and after adjusting for all other covariates. “1” represent the R2 from the model with the biomarker levels of a given chemical as the outcome variable and the occupational groups as the main predictor. “2” represents the difference in R2 between the fully adjusted model and those adjusted for all other covariates except for occupation. Chemicals are ordered by the contribution of occupation explaining the variance of the chemical biomarker levels after adjusting for all other covariates (i.e. chemicals are ordered by the magnitude of the “2”s).

**Formatted:** Position: Horizontal: Right, Relative to: Margin, Vertical: 0", Relative to: Paragraph, Wrap Around

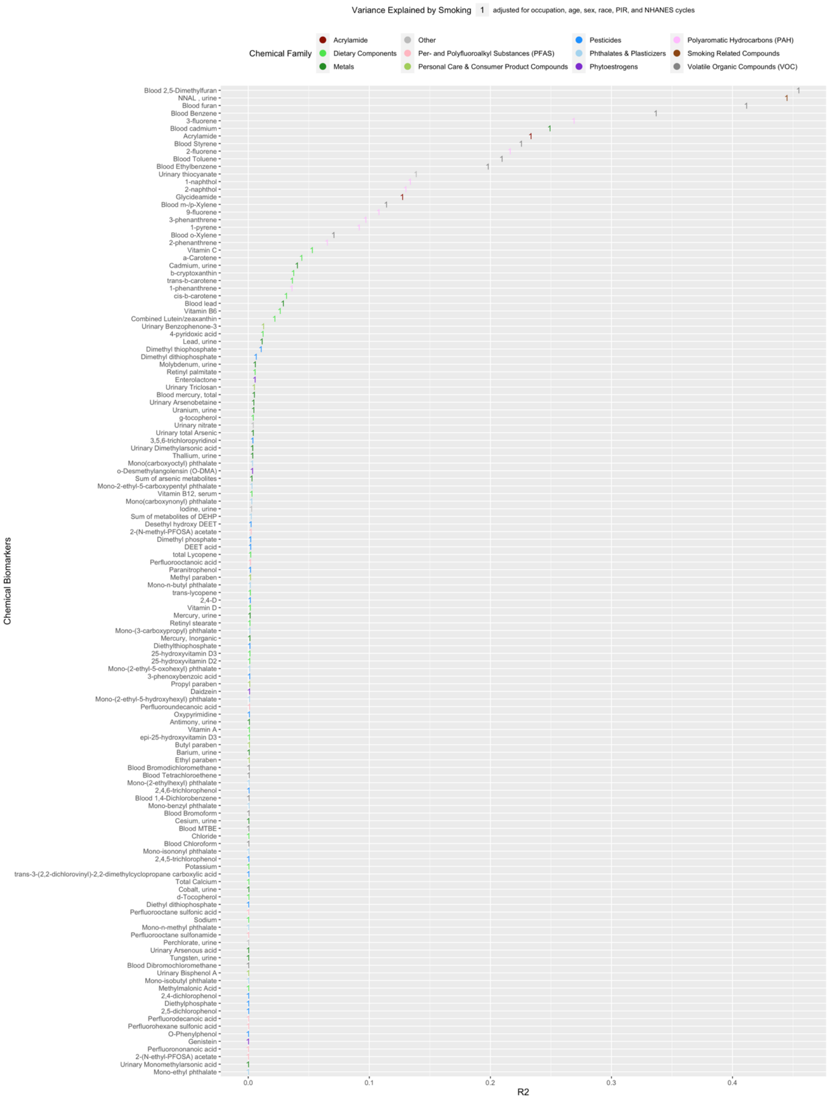

Figure S16. Alphabet soup plot of variance explained by smoking after adjusting for all other covariates. "1" represents the difference in R2 between the fully adjusted model and those adjusted for all other covariates except for smoking.

**Formatted:** Position: Horizontal: Right, Relative to: Margin, Vertical: 0", Relative to: Paragraph, Wrap Around

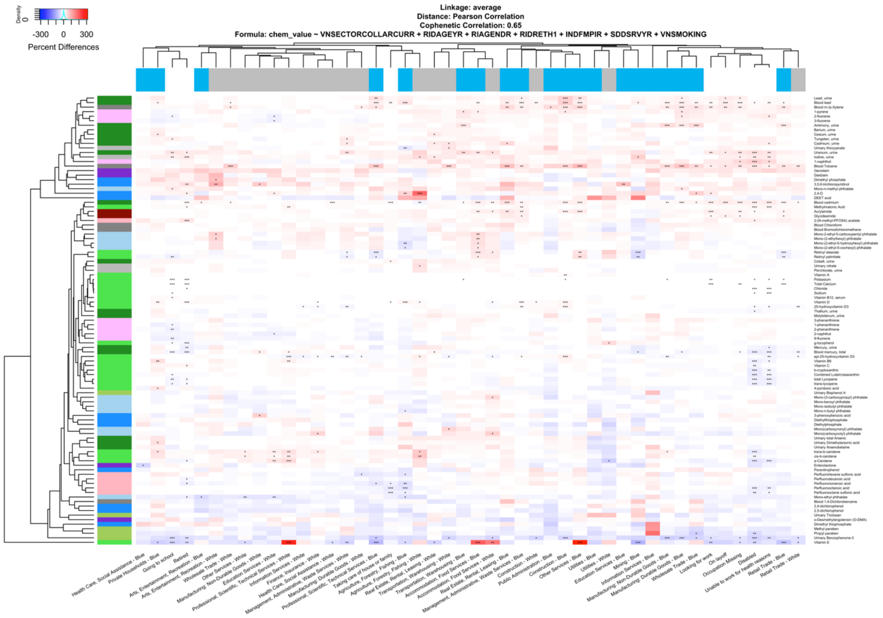

Figure S17. Heatmap of percent differences in chemical biomarker concentrations by occupational group, relative to white collars from Public Administration. Cotinine and NNAL were excluded from the clustering analysis used to form the dendrogram. Chemical biomarkers in white color indicates that the concentrations are the same between the given sector-collar combination and the reference group. The color bar for the columns represents the collar categorization and unemployment. Blue presents the blue-collar workers. White represents the white-collar workers. Gray presents the unemployed participants. Results are adjusted for age, sex, race/ethnicity, poverty income ratio, study period, and serum cotinine (biomarker of smoking). Number of asterisks indicate statistical significance of the percent differences: * (p-value ∈ (0.01, 0.05]), ** (p-value ∈ (0.001, 0.01]), and *** (p-value ≤ 0.001).

**Formatted:** Position: Horizontal: Right, Relative to: Margin, Vertical: 0", Relative to: Paragraph, Wrap Around

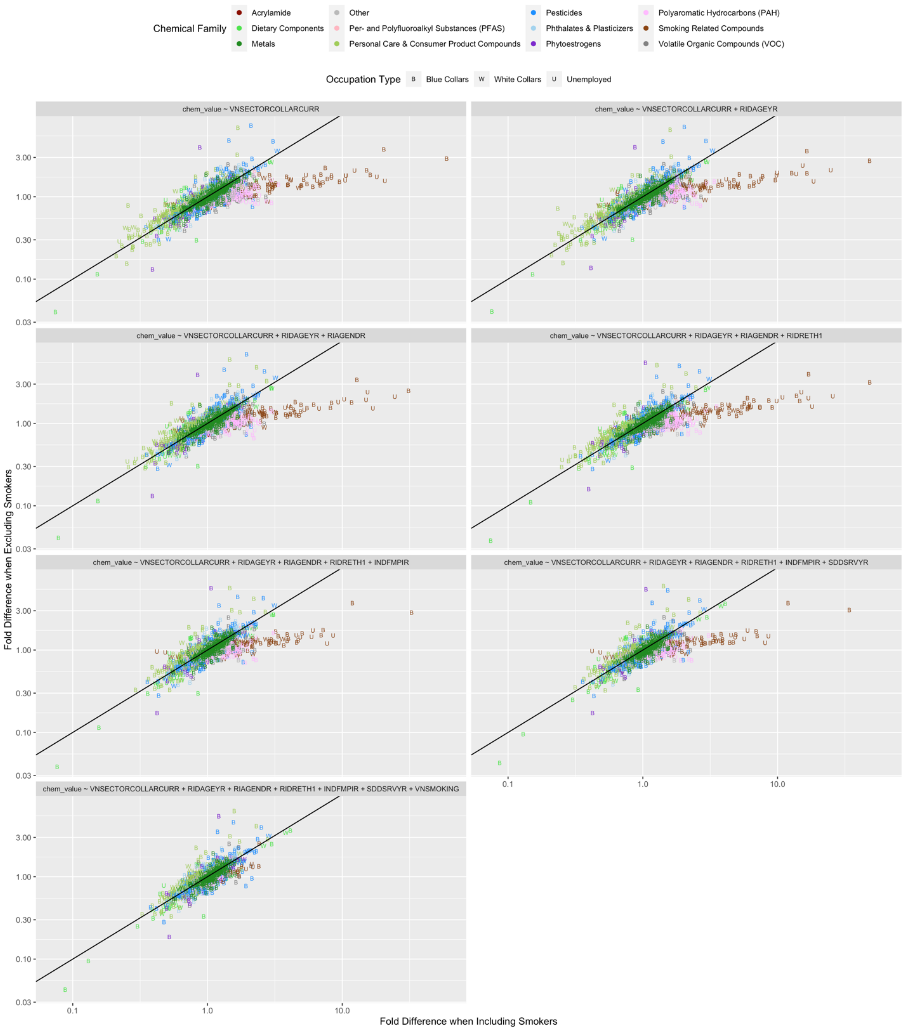

**Figure S18. Panel of correlation plots comparing percent difference in chemical biomarker levels when including active smokers versus excluding active smokers across the different stepwise regression models.**

**Formatted:** Position: Horizontal: Right, Relative to: Margin, Vertical: 0", Relative to: Paragraph, Wrap Around
